## Supplemental Table 3 for "The spread of SARS-CoV-2 variant Omicron with the doubling time of 2.0–3.3 days can be explained by immune evasion"

We gratefully acknowledge the following Authors from the Originating laboratories responsible for obtaining the specimens, as well as the Submitting laboratories where the genome data were generated and shared via GISAID, on which this research is based.

All Submitters of data may be contacted directly via www.gisaid.org

Authors are sorted alphabetically.

| Accession ID | Originating Laboratory | Submitting Laboratory | Authors |
|---|---|---|---|
| EPI_ISL_2444137, EPI_ISL_2444138, EPI_ISL_2444139, EPI_ISL_2444140, EPI_ISL_2444141, EPI_ISL_2444142, EPI_ISL_2444143, EPI_ISL_2444144, EPI_ISL_2444145, EPI_ISL_2444146, EPI_ISL_2444147, EPI_ISL_2444148, EPI_ISL_2444149, EPI_ISL_2444150, EPI_ISL_2444151, EPI_ISL_2444152 | | | |
| see above | AAMPATH LABORATORIES | National Institute for Communicable Diseases of the National Health Laboratory Service | Amoako DG; Bhiman JN; Ismail A; Mahlangu B; Mohale T; Ntuli N; Scheepers C |
| EPI_ISL_7886687, EPI_ISL_7886688, EPI_ISL_7886689, EPI_ISL_7886690 | AHRI - Alex Sigal lab | KRISP, KZn Research Innovation and Sequencing Platform | Arisha Maharaj; Bernstein Mallory; Cele Sandile; Giandhari J; Karim Farina; Khan Khadija; Lessells R; Moir M; Naidoo Y; Pillay S; Ramphal U; Ramphal Y; San JE; Sigal Alex; Tegally H; Tshiabuila D; Wilkinson E; de Oliveira T |
| EPI_ISL_3838578, EPI_ISL_3838579, EPI_ISL_3838580, EPI_ISL_3838581, EPI_ISL_3838583, EPI_ISL_3838584, EPI_ISL_3838586, EPI_ISL_3838594, EPI_ISL_3838595, EPI_ISL_3838596, EPI_ISL_3838597, EPI_ISL_3838598, EPI_ISL_4651621, EPI_ISL_4651622, EPI_ISL_4651629, EPI_ISL_4651637, EPI_ISL_4651641, EPI_ISL_4651646, EPI_ISL_4651652, EPI_ISL_4651660, EPI_ISL_4651664, EPI_ISL_4651665, EPI_ISL_4651673, EPI_ISL_4651678, EPI_ISL_4651692, EPI_ISL_4651700, EPI_ISL_4651701, EPI_ISL_4651703, EPI_ISL_4651714, EPI_ISL_4651719, EPI_ISL_4651720, EPI_ISL_4651727, EPI_ISL_4651741, EPI_ISL_4651742, EPI_ISL_4651748, EPI_ISL_4651751, EPI_ISL_4651771, EPI_ISL_4651782, EPI_ISL_4651786, EPI_ISL_4651795, EPI_ISL_4651796, EPI_ISL_4651805, EPI_ISL_4651809, EPI_ISL_4651816, EPI_ISL_4651823, EPI_ISL_4651831, EPI_ISL_4651832, EPI_ISL_4651843, EPI_ISL_4651849, EPI_ISL_4651854, EPI_ISL_4651875, EPI_ISL_4651888, EPI_ISL_4651914, EPI_ISL_4651930, EPI_ISL_4652093, EPI_ISL_4652103, EPI_ISL_4652108, EPI_ISL_4652117, EPI_ISL_4652127, EPI_ISL_4652135, EPI_ISL_4652141, EPI_ISL_4652145, EPI_ISL_4652146, EPI_ISL_4652153, EPI_ISL_4652158, EPI_ISL_4652159, EPI_ISL_4652169, EPI_ISL_4652170, EPI_ISL_4652175, EPI_ISL_4652180, EPI_ISL_4652181, EPI_ISL_4652185, EPI_ISL_4652186, EPI_ISL_4652191, EPI_ISL_4652194, EPI_ISL_4652195, EPI_ISL_4652201, EPI_ISL_5416273, EPI_ISL_5416289, EPI_ISL_5416298, EPI_ISL_5416299, EPI_ISL_5416344, EPI_ISL_5416371, EPI_ISL_5416508, EPI_ISL_5416509, EPI_ISL_5416510, EPI_ISL_5416511, EPI_ISL_5416512, EPI_ISL_5416513, EPI_ISL_5416514, EPI_ISL_5416515, EPI_ISL_5416516, EPI_ISL_5416517, EPI_ISL_5416518, EPI_ISL_5416519, EPI_ISL_5416520, EPI_ISL_5416522, EPI_ISL_5476869, EPI_ISL_5510394, EPI_ISL_5603071, EPI_ISL_5603072, EPI_ISL_5603073, EPI_ISL_5603075, EPI_ISL_5603076, EPI_ISL_5603077, EPI_ISL_5603078, EPI_ISL_5603079, EPI_ISL_5603080, EPI_ISL_5603081, EPI_ISL_5603082, EPI_ISL_5603083, EPI_ISL_5603084, EPI_ISL_5603120, EPI_ISL_5603121, EPI_ISL_5603130, EPI_ISL_5603139, EPI_ISL_5603156, EPI_ISL_5603159, EPI_ISL_5603161, EPI_ISL_5603166, EPI_ISL_5603172, EPI_ISL_5603183, EPI_ISL_6203619, EPI_ISL_6327724, EPI_ISL_6327725, EPI_ISL_6327726, EPI_ISL_6327727, EPI_ISL_6327728, EPI_ISL_6327729, EPI_ISL_6327730, EPI_ISL_6327731, EPI_ISL_6327836, EPI_ISL_6327850, EPI_ISL_6327862, EPI_ISL_6327865, EPI_ISL_6327880, EPI_ISL_6327881, EPI_ISL_6327882, EPI_ISL_6327896, EPI_ISL_6327897, EPI_ISL_6327902, EPI_ISL_6327913, EPI_ISL_6327917, EPI_ISL_6327920, EPI_ISL_6327931, EPI_ISL_6327938, EPI_ISL_6327962, EPI_ISL_6327970, EPI_ISL_6327977, EPI_ISL_6327987, EPI_ISL_6327996, EPI_ISL_6900030, EPI_ISL_6900031, EPI_ISL_6900033, EPI_ISL_6900037, EPI_ISL_6900042, EPI_ISL_6900044, EPI_ISL_6900050, EPI_ISL_6900051, EPI_ISL_6900052, EPI_ISL_6900055, EPI_ISL_6900058, EPI_ISL_6900059, EPI_ISL_6900061, EPI_ISL_6900062, EPI_ISL_6900063, EPI_ISL_6900064, EPI_ISL_6900065, EPI_ISL_6900072, EPI_ISL_6900076, EPI_ISL_6900078, EPI_ISL_6900079, EPI_ISL_6900080, EPI_ISL_6900081, EPI_ISL_6900082, EPI_ISL_6900083, EPI_ISL_6900084, EPI_ISL_6900087 | | | |
| see above | AMPATH | National Institute for Communicable Diseases of the National Health Laboratory Service | Amoako DG; Bhiman JN; Everatt J; Ismail A; Mahlangu B; Mnguni A; Mohale T; Ntuli N; Scheepers C |
| EPI_ISL_2086984, EPI_ISL_2086985, EPI_ISL_2086986, EPI_ISL_2086996, EPI_ISL_2086997, EPI_ISL_2086998, EPI_ISL_2086999, EPI_ISL_2087000, EPI_ISL_2087002, EPI_ISL_2087005, EPI_ISL_2087006, EPI_ISL_2087008, EPI_ISL_2087009, EPI_ISL_2087010, EPI_ISL_2087012, EPI_ISL_2087013, EPI_ISL_2087014, EPI_ISL_2087018, EPI_ISL_2087019, EPI_ISL_2087022, EPI_ISL_2087024 | | | |
| see above | AMPATH LABORATORIES | National Institute for Communicable Diseases of the National Health Laboratory Service | Amoako DG; Bhiman JN; Ismail A; Mahlangu B; Mohale T; Ntuli N; Scheepers C |
| EPI_ISL_4029909, EPI_ISL_4029924, EPI_ISL_4029937, EPI_ISL_4029951, EPI_ISL_4029966, EPI_ISL_4029981, EPI_ISL_4029982, EPI_ISL_4029988, EPI_ISL_4029992, EPI_ISL_4029993, EPI_ISL_4029998, EPI_ISL_4030002, EPI_ISL_4030004, EPI_ISL_4030005, EPI_ISL_4030006, EPI_ISL_4030007, EPI_ISL_4030008, EPI_ISL_4030009, EPI_ISL_4030010, EPI_ISL_4030015, EPI_ISL_4030062, EPI_ISL_4030063, EPI_ISL_4030064, EPI_ISL_4030066, EPI_ISL_4030067 | | | |
| see above | AMPATH LABORATORY | National Institute for Communicable Diseases of the National Health Laboratory Service | Amoako DG; Bhiman JN; Everatt J; Ismail A; Mahlangu B; Mnguni A; Mohale T; Ntuli N; Scheepers C |
| EPI_ISL_5917889, EPI_ISL_5917982, EPI_ISL_5917984, EPI_ISL_5918032, EPI_ISL_5918041, EPI_ISL_5918047, EPI_ISL_5918050, EPI_ISL_5918074, EPI_ISL_5918090, EPI_ISL_5918092, EPI_ISL_5918101, EPI_ISL_5918107, EPI_ISL_5919986, EPI_ISL_5919994, EPI_ISL_5920002, EPI_ISL_5920016, EPI_ISL_5920102, EPI_ISL_5920133, EPI_ISL_5920137, EPI_ISL_5920155, EPI_ISL_5920159, EPI_ISL_5920164, EPI_ISL_5920170, EPI_ISL_5920196, EPI_ISL_5920204, EPI_ISL_5920214 | | | |
| see above | Ampath | National Institute for Communicable Diseases of the National Health Laboratory Service | Amoako DG; Bhiman JN; Everatt J; Ismail A; Mahlangu B; Mnguni A; Mohale T; Ntuli N; Scheepers C |
| EPI_ISL_2178779, EPI_ISL_2178791, EPI_ISL_2178792, EPI_ISL_2193354, EPI_ISL_2193355, EPI_ISL_2193356, EPI_ISL_2193357, EPI_ISL_2193358, EPI_ISL_2193359, EPI_ISL_2193360, EPI_ISL_2193361, EPI_ISL_6914011, EPI_ISL_6914012, EPI_ISL_6914013, EPI_ISL_6914015, EPI_ISL_6914016, EPI_ISL_6914017, EPI_ISL_6914018, EPI_ISL_6914020, EPI_ISL_6914021, EPI_ISL_6914022, EPI_ISL_6914023, EPI_ISL_6914024, EPI_ISL_6914025, EPI_ISL_6914026, EPI_ISL_6914027, EPI_ISL_6914029, EPI_ISL_6914030, EPI_ISL_6914031, EPI_ISL_6914032, EPI_ISL_6914033, EPI_ISL_6914034, EPI_ISL_6914036 | | | |
| see above | Ampath Laboratories | National Institute for Communicable Diseases of the National Health Laboratory Service | Amoako DG; Bhiman JN; Everatt J; Glass A; Gottberg A; Ismail A; Mahlangu B; Marshall T; Mnguni A; Mohale T; Ntuli N; Scheepers C; Viana R; Wolter N |
| EPI_ISL_2893300, EPI_ISL_2893301, EPI_ISL_2893302, EPI_ISL_2893303, EPI_ISL_2893304, EPI_ISL_2893305, EPI_ISL_2893306, EPI_ISL_2893307, EPI_ISL_2893308, EPI_ISL_2893309, EPI_ISL_2893310, EPI_ISL_2893311, EPI_ISL_2893312, EPI_ISL_2893313, EPI_ISL_2893314 | | | |
| see above | BARC - Sisonke | KRISP, KZN Research Innovation and Sequencing Platform | Giandhari Jennifer; Naidoo Yeshnee; Pillay Sureshnee; San James; Sisonke; Tegally Houriiyah; Tshabuila Derek; Wilkinson Eduan; Yajna Ramphal; de Oliveira Tulio |
| EPI_ISL_2710306, EPI_ISL_2710307, EPI_ISL_2710308, EPI_ISL_2710309, EPI_ISL_2710310, EPI_ISL_2710312, EPI_ISL_2710313, EPI_ISL_2710314, EPI_ISL_2710315, EPI_ISL_2710316, EPI_ISL_2710317, EPI_ISL_2710318, EPI_ISL_2710319, EPI_ISL_2710320, EPI_ISL_2710321 | | | |
| see above | BARC / Lancet | KRISP, KZN Research Innovation and Sequencing Platform | Giandhari Jennifer; Naidoo Yeshnee; Pillay Sureshnee; San James; Sisonke; Tegally Houriiyah; Tshabuila Derek; Wilkinson Eduan; Yajna Ramphal; de Oliveira Tulio |
| EPI_ISL_5510285, EPI_ISL_5510294, EPI_ISL_5510298, EPI_ISL_5510351, EPI_ISL_5510367, EPI_ISL_5510370, EPI_ISL_5510378, EPI_ISL_5510383, EPI_ISL_5510396, EPI_ISL_5510397, EPI_ISL_5510401, EPI_ISL_5510407, EPI_ISL_5510410, EPI_ISL_5510411, EPI_ISL_5510445, EPI_ISL_5510447, EPI_ISL_5510448, EPI_ISL_5510452, EPI_ISL_5510453, EPI_ISL_5510462, EPI_ISL_5510471, EPI_ISL_5510490, EPI_ISL_5510491 | | | |
| see above | Baragwanath | National Institute for Communicable Diseases of the National Health Laboratory Service | Amoako DG; Bhiman JN; Everatt J; Ismail A; Mahlangu B; Mnguni A; Mohale T; Ntuli N; Scheepers C |
| EPI_ISL_2695506 | Bertha gxowa hospital | National Institute for Communicable Diseases of the National Health Laboratory Service | Amoako DG; Bhiman JN; Everatt J; Ismail A; Mahlangu B; Mnguni A; Mohale T; Ntuli N; Scheepers C |
| EPI_ISL_5510365, EPI_ISL_5510443 | Braamfontein Central | National Institute for Communicable Diseases of the National Health Laboratory Service | Amoako DG; Bhiman JN; Everatt J; Ismail A; Mahlangu B; Mnguni A; Mohale T; Ntuli N; Scheepers C |
| EPI_ISL_2086880, EPI_ISL_2086881, EPI_ISL_2086882, EPI_ISL_2086883, EPI_ISL_2086884, EPI_ISL_2086885, EPI_ISL_2086886, EPI_ISL_2086887, EPI_ISL_2086888, EPI_ISL_2086889, EPI_ISL_2086890, EPI_ISL_2086891, EPI_ISL_2086892, EPI_ISL_2086893, EPI_ISL_2086894, EPI_ISL_2086895, EPI_ISL_2086896, EPI_ISL_2086897, EPI_ISL_2086898, EPI_ISL_2086899, EPI_ISL_2086900, EPI_ISL_2086901, EPI_ISL_2086902, EPI_ISL_2086903, EPI_ISL_2285233, EPI_ISL_2285234, EPI_ISL_2285235, EPI_ISL_2285236, EPI_ISL_2285237, EPI_ISL_2285238, EPI_ISL_2285239, EPI_ISL_2285337, EPI_ISL_2285338 | | | |
| see above | CHARLOTTE MAXEKE JOHANNESBURG ACADEMIC HOSPITAL | National Institute for Communicable Diseases of the National Health Laboratory Service | Amoako DG; Bhiman JN; Ismail A; Mahlangu B; Mohale T; Ntuli N; Scheepers C |
| EPI_ISL_2694627, EPI_ISL_2694628, EPI_ISL_2694629, EPI_ISL_2694630, EPI_ISL_2694631, EPI_ISL_2694633, EPI_ISL_2694634, EPI_ISL_2694635, EPI_ISL_2694636, EPI_ISL_2694637, EPI_ISL_2694638, EPI_ISL_2694639, EPI_ISL_2694640, EPI_ISL_2694641, EPI_ISL_2694642, EPI_ISL_2694643, EPI_ISL_2694644, EPI_ISL_2694645, EPI_ISL_2694646, EPI_ISL_2694647, EPI_ISL_2694648, EPI_ISL_2694649, EPI_ISL_2694650, EPI_ISL_2694651, EPI_ISL_2694652, EPI_ISL_2694653, EPI_ISL_2694654, EPI_ISL_2694655, EPI_ISL_2694656, EPI_ISL_2694657, EPI_ISL_2694658, EPI_ISL_2694659, EPI_ISL_2694660, EPI_ISL_2694661, EPI_ISL_2694662, EPI_ISL_2694663, EPI_ISL_2694664, EPI_ISL_2694665, EPI_ISL_2694666, EPI_ISL_2694667, EPI_ISL_2694668, EPI_ISL_2694669, EPI_ISL_2694670, EPI_ISL_2694671, EPI_ISL_2694672, EPI_ISL_2694673, EPI_ISL_2694674, EPI_ISL_2694675, EPI_ISL_2694676, EPI_ISL_2694677, EPI_ISL_2709879, EPI_ISL_2709880, EPI_ISL_2709881, EPI_ISL_2709882, EPI_ISL_2709883, EPI_ISL_2709884, EPI_ISL_2709887, EPI_ISL_2709888, EPI_ISL_2709889, EPI_ISL_2709890, EPI_ISL_2709891, EPI_ISL_2709892, EPI_ISL_2709893, EPI_ISL_2709894, EPI_ISL_2709895, EPI_ISL_2709896, EPI_ISL_2709897, EPI_ISL_2709898, EPI_ISL_2709899, EPI_ISL_2709900, EPI_ISL_2709901, EPI_ISL_2709902, EPI_ISL_2709903, EPI_ISL_2709904, EPI_ISL_2709905, EPI_ISL_2709906, EPI_ISL_2709907, EPI_ISL_2709908, EPI_ISL_2709909, EPI_ISL_2709910, EPI_ISL_2709911, EPI_ISL_2709912, EPI_ISL_2709913, EPI_ISL_2709914, EPI_ISL_2709915, EPI_ISL_2709917, EPI_ISL_2709918, EPI_ISL_2709919, EPI_ISL_2709920, EPI_ISL_2709921, EPI_ISL_2709922, EPI_ISL_2709923, EPI_ISL_2709924, EPI_ISL_2709925, EPI_ISL_2709926, EPI_ISL_2709927, EPI_ISL_2709928, EPI_ISL_2709929, EPI_ISL_2709930, EPI_ISL_2709931, EPI_ISL_2709932, EPI_ISL_2709933, EPI_ISL_2709934, EPI_ISL_2709935, EPI_ISL_2709936, EPI_ISL_2709937, EPI_ISL_2709938, EPI_ISL_2709939, EPI_ISL_2709940, EPI_ISL_2709941, EPI_ISL_2709942, EPI_ISL_5510470 | | | |
| see above | CHRIS HANI BARAGWANATH | National Institute for Communicable Diseases of the National Health Laboratory Service | Amoako DG; Bhiman JN; Everatt J; Ismail A; Mahlangu B; Mnguni A; Mohale T; Ntuli N; Scheepers C |
| EPI_ISL_5510372 | CHRIS HANI BARAGWANATH HOSPITAL | National Institute for Communicable Diseases of the National Health Laboratory Service | Amoako DG; Bhiman JN; Everatt J; Ismail A; Mahlangu B; Mnguni A; Mohale T; Ntuli N; Scheepers C |
| EPI_ISL_2086904, EPI_ISL_2086905, EPI_ISL_2086906, EPI_ISL_2086907, EPI_ISL_2086908, EPI_ISL_2086910, EPI_ISL_2086911, EPI_ISL_2086912, EPI_ISL_2086913, EPI_ISL_2086915, EPI_ISL_2086921, EPI_ISL_2086922, EPI_ISL_2086923, EPI_ISL_2086924, EPI_ISL_2087030, EPI_ISL_2087031, EPI_ISL_2087032, EPI_ISL_2087033, EPI_ISL_2087034, EPI_ISL_2087035, EPI_ISL_2087036, EPI_ISL_2087037, EPI_ISL_2087038, EPI_ISL_2087039, EPI_ISL_2087040, EPI_ISL_2087041, EPI_ISL_2087042, EPI_ISL_2087043, EPI_ISL_2087044, EPI_ISL_2087045, EPI_ISL_2087046, EPI_ISL_2087047, EPI_ISL_2153516, EPI_ISL_2153517, EPI_ISL_2153518, EPI_ISL_2153519, EPI_ISL_2153520, EPI_ISL_2153521, EPI_ISL_2153522, EPI_ISL_2153523, EPI_ISL_2153525, EPI_ISL_2153526, EPI_ISL_2153527, EPI_ISL_2153528, EPI_ISL_2153529, EPI_ISL_2153530, EPI_ISL_2153531, EPI_ISL_2153532, EPI_ISL_2153533, EPI_ISL_2153534, EPI_ISL_2153535, EPI_ISL_2153536, EPI_ISL_2153537, EPI_ISL_2273005, EPI_ISL_2273006, EPI_ISL_2273007, EPI_ISL_2273008, EPI_ISL_2273009, EPI_ISL_2273010, EPI_ISL_2273011, EPI_ISL_2273012, EPI_ISL_2273013, EPI_ISL_2273014, EPI_ISL_2273015, EPI_ISL_2273016, EPI_ISL_2273017, EPI_ISL_2273018, EPI_ISL_2273019, EPI_ISL_2273020, EPI_ISL_2273021, EPI_ISL_2273022, EPI_ISL_2273023, EPI_ISL_2273024, EPI_ISL_2273025, EPI_ISL_2273026, EPI_ISL_2273027, EPI_ISL_2273028, EPI_ISL_2273029, EPI_ISL_2273030, EPI_ISL_2273031, EPI_ISL_2273032, EPI_ISL_2273033, EPI_ISL_2273034, EPI_ISL_2273035, EPI_ISL_2273036, EPI_ISL_2273037, EPI_ISL_2273038, EPI_ISL_2273039, EPI_ISL_2273040, EPI_ISL_2273041, EPI_ISL_2273042, EPI_ISL_2273043, EPI_ISL_2273044, EPI_ISL_2273045, EPI_ISL_2273046, EPI_ISL_2273047, EPI_ISL_2273048, EPI_ISL_2273049, EPI_ISL_2273050, EPI_ISL_2273051, EPI_ISL_2273052, EPI_ISL_2273053, EPI_ISL_2273054, EPI_ISL_2273055, EPI_ISL_2273056, EPI_ISL_2285249, EPI_ISL_2285250, EPI_ISL_2382225, EPI_ISL_2382226, EPI_ISL_2382227, EPI_ISL_2382228, EPI_ISL_2382229, EPI_ISL_2382230, EPI_ISL_2382231, EPI_ISL_2382232, EPI_ISL_2382233, EPI_ISL_2382234, EPI_ISL_2382235, EPI_ISL_2382236, EPI_ISL_2382237, EPI_ISL_2382238, EPI_ISL_2382239, EPI_ISL_2382240, EPI_ISL_2382241, EPI_ISL_2382242, EPI_ISL_2382243, EPI_ISL_2382244, EPI_ISL_2382245, EPI_ISL_2382246, EPI_ISL_2382247, EPI_ISL_2382248, EPI_ISL_2382249, EPI_ISL_2382250, EPI_ISL_2382251, EPI_ISL_2382252, EPI_ISL_2382253, EPI_ISL_2382254, EPI_ISL_2382255, EPI_ISL_2382256, EPI_ISL_2382257, EPI_ISL_2382258, EPI_ISL_2382259, EPI_ISL_2382676, EPI_ISL_2382677, EPI_ISL_2382678, EPI_ISL_2382679, EPI_ISL_2382680, EPI_ISL_2382681, EPI_ISL_2382682, EPI_ISL_2382683, EPI_ISL_2382684, EPI_ISL_2382685, EPI_ISL_2382686, EPI_ISL_2382687, EPI_ISL_2382688, EPI_ISL_2382689, EPI_ISL_2382690, EPI_ISL_2382691, EPI_ISL_2382692, EPI_ISL_2382693, EPI_ISL_2382694, EPI_ISL_2382695, EPI_ISL_2382696, EPI_ISL_2382697, EPI_ISL_2382698, EPI_ISL_2442366, EPI_ISL_2442367, EPI_ISL_2442368, EPI_ISL_2442369, EPI_ISL_2454623, EPI_ISL_2454624, EPI_ISL_2454625, EPI_ISL_2454626, EPI_ISL_2454627, EPI_ISL_2454628, EPI_ISL_2454629, EPI_ISL_2454630, EPI_ISL_2454631, EPI_ISL_2454632, EPI_ISL_2454633, EPI_ISL_2454634, EPI_ISL_2454635, EPI_ISL_2454636, EPI_ISL_2454637, EPI_ISL_2454638, EPI_ISL_2454639, EPI_ISL_2454640, EPI_ISL_2454641, EPI_ISL_2454642, EPI_ISL_2454643, EPI_ISL_2454644, EPI_ISL_2454645, EPI_ISL_2454646, EPI_ISL_2454647, EPI_ISL_2454648, EPI_ISL_2454649, EPI_ISL_2454650, EPI_ISL_2454651, EPI_ISL_2454652, EPI_ISL_2454653, EPI_ISL_2454654, EPI_ISL_2454655, EPI_ISL_2454656, EPI_ISL_2454657, EPI_ISL_2454658, EPI_ISL_2454659, EPI_ISL_2582812, EPI_ISL_2582815, EPI_ISL_2582818, EPI_ISL_2582822, EPI_ISL_2582825, EPI_ISL_2662468, EPI_ISL_2662469, EPI_ISL_2662470, EPI_ISL_2662471, EPI_ISL_2662472, EPI_ISL_2662473, EPI_ISL_2662474, EPI_ISL_2662492, EPI_ISL_2662493, EPI_ISL_2662494, EPI_ISL_2662495, EPI_ISL_2662496, EPI_ISL_2662497, EPI_ISL_2662498, EPI_ISL_2662499, EPI_ISL_2662500, EPI_ISL_2662501, EPI_ISL_2662502, EPI_ISL_2662503, EPI_ISL_2662504, EPI_ISL_2662505, EPI_ISL_2662506, EPI_ISL_2662507, EPI_ISL_2662508, EPI_ISL_2662509, EPI_ISL_2662510, EPI_ISL_2662554, EPI_ISL_2662555, EPI_ISL_2662618, EPI_ISL_2662619, EPI_ISL_2662620, EPI_ISL_2662621, EPI_ISL_2662622, EPI_ISL_2662623, EPI_ISL_2662624, EPI_ISL_2662625, EPI_ISL_2662626, EPI_ISL_2662627, EPI_ISL_2662628, EPI_ISL_2662629, EPI_ISL_2662630, EPI_ISL_2662631, EPI_ISL_2662632, EPI_ISL_2662633, EPI_ISL_2662634, EPI_ISL_2662635, EPI_ISL_2662636, EPI_ISL_2662637, EPI_ISL_2662638, EPI_ISL_2662639, EPI_ISL_2662640, EPI_ISL_2662641, EPI_ISL_2662642, EPI_ISL_2662643, EPI_ISL_2662644, EPI_ISL_2662645, EPI_ISL_2662646, EPI_ISL_2662647, EPI_ISL_2662648, EPI_ISL_2662649, EPI_ISL_2662650, EPI_ISL_2662651, EPI_ISL_2662652, EPI_ISL_2662653, EPI_ISL_2662654, EPI_ISL_2662655, EPI_ISL_2662656, EPI_ISL_2662657, EPI_ISL_2662658, EPI_ISL_2662659, EPI_ISL_2662660, EPI_ISL_2662661, EPI_ISL_2827112, EPI_ISL_2827156, EPI_ISL_2827157, EPI_ISL_3024646, EPI_ISL_3024647, EPI_ISL_3024648, EPI_ISL_3024649, EPI_ISL_3024650, EPI_ISL_3024651, EPI_ISL_3024652, EPI_ISL_3024653, EPI_ISL_3024654, EPI_ISL_3024655, EPI_ISL_3024656, EPI_ISL_3024657, EPI_ISL_3024658, EPI_ISL_3024659, EPI_ISL_3024660, EPI_ISL_3024661, EPI_ISL_3024662, EPI_ISL_3024663, EPI_ISL_3024664, EPI_ISL_3024665, EPI_ISL_3024666, EPI_ISL_3024667, EPI_ISL_3024668, EPI_ISL_3024669, EPI_ISL_3024670, EPI_ISL_3024671, EPI_ISL_3024672, EPI_ISL_3024673, EPI_ISL_3024674, EPI_ISL_3024675, EPI_ISL_3024676, EPI_ISL_3024677, EPI_ISL_3024678, EPI_ISL_3024679, EPI_ISL_3024680, EPI_ISL_3024681, EPI_ISL_3030439, EPI_ISL_3030440, EPI_ISL_3451304, EPI_ISL_3451305, EPI_ISL_3451306, EPI_ISL_3451307, EPI_ISL_3451308, EPI_ISL_3451309, EPI_ISL_3451310, EPI_ISL_3451311, EPI_ISL_3451312, EPI_ISL_3451313, EPI_ISL_3451314, EPI_ISL_3451315, EPI_ISL_3451316, EPI_ISL_3451317, EPI_ISL_3451318, EPI_ISL_3451319, EPI_ISL_3451320, EPI_ISL_3451321, EPI_ISL_3451322, EPI_ISL_3451323, EPI_ISL_3451324, EPI_ISL_3451325, EPI_ISL_3451326, EPI_ISL_3451327, EPI_ISL_3451328, EPI_ISL_3451329, EPI_ISL_3451439, EPI_ISL_3451440, EPI_ISL_3451441, EPI_ISL_3451442, EPI_ISL_3838343, EPI_ISL_3838344, EPI_ISL_3838345, EPI_ISL_3838346, EPI_ISL_3838347, EPI_ISL_3838348, EPI_ISL_3838349, EPI_ISL_3838350, EPI_ISL_3838351, EPI_ISL_3838352, EPI_ISL_3838353, EPI_ISL_3838354, EPI_ISL_3838355, EPI_ISL_3838356, EPI_ISL_3838503, EPI_ISL_3838504, EPI_ISL_3838505, EPI_ISL_3838506, EPI_ISL_3838507, EPI_ISL_3838508, EPI_ISL_3838509, EPI_ISL_3838559, EPI_ISL_3838560, EPI_ISL_3838561, EPI_ISL_3838562, EPI_ISL_3838563, EPI_ISL_3838564, EPI_ISL_3838565, EPI_ISL_5416339, EPI_ISL_5416353, EPI_ISL_5416354, EPI_ISL_5416362, EPI_ISL_5416368, EPI_ISL_5416475, EPI_ISL_6201546, EPI_ISL_6201554, EPI_ISL_6201569, EPI_ISL_6201919, EPI_ISL_6202099, EPI_ISL_6202172, EPI_ISL_6202591, EPI_ISL_6327743, EPI_ISL_6327744, EPI_ISL_6327838, EPI_ISL_6327839, EPI_ISL_6327847, EPI_ISL_6327869, EPI_ISL_6327879, EPI_ISL_6327925, EPI_ISL_6327947, EPI_ISL_6327973, EPI_ISL_6693527, EPI_ISL_6693528, EPI_ISL_6693529, EPI_ISL_6693530, EPI_ISL_6693531, EPI_ISL_6693532, EPI_ISL_6693533, EPI_ISL_6693534, EPI_ISL_6693535, EPI_ISL_6693536, EPI_ISL_6693537, EPI_ISL_6693538, EPI_ISL_6693539, EPI_ISL_6693540, EPI_ISL_6693541, EPI_ISL_6693542, EPI_ISL_6693546, EPI_ISL_6693563, EPI_ISL_6693565, EPI_ISL_6693571, EPI_ISL_6693575, EPI_ISL_6693581, EPI_ISL_6693588, EPI_ISL_6693597, EPI_ISL_6693608, EPI_ISL_6693614, EPI_ISL_6693617, EPI_ISL_6693620, EPI_ISL_6693628, EPI_ISL_6693630, EPI_ISL_6900016, EPI_ISL_6900017, EPI_ISL_6900019, EPI_ISL_6900020, EPI_ISL_6906406, EPI_ISL_6906448, EPI_ISL_6906474, EPI_ISL_6906491, EPI_ISL_7971610, EPI_ISL_7971611, EPI_ISL_7971613, EPI_ISL_7971614, EPI_ISL_7971615, EPI_ISL_7971646, EPI_ISL_7971647, EPI_ISL_7971648, EPI_ISL_7971649, EPI_ISL_7971650, EPI_ISL_7971651, EPI_ISL_7971652, EPI_ISL_7971653, EPI_ISL_7971654, EPI_ISL_7971674, EPI_ISL_7971675, EPI_ISL_7971676, EPI_ISL_7971677, EPI_ISL_7971678, EPI_ISL_7971679, EPI_ISL_7971680, EPI_ISL_7971681, EPI_ISL_7971682, EPI_ISL_7971683, EPI_ISL_7971684, EPI_ISL_7971685, EPI_ISL_7971686, EPI_ISL_7971687, EPI_ISL_7971688, EPI_ISL_7971689, EPI_ISL_7971690, EPI_ISL_7971691, EPI_ISL_7971692, EPI_ISL_8182816, EPI_ISL_8182817, EPI_ISL_8182818, EPI_ISL_8182819, EPI_ISL_8182820, EPI_ISL_8182821, EPI_ISL_8182822, EPI_ISL_8182823, EPI_ISL_8182824, EPI_ISL_8182825, EPI_ISL_8182826, EPI_ISL_8182827, EPI_ISL_8182828, EPI_ISL_8182829, EPI_ISL_8182857, EPI_ISL_8182858, EPI_ISL_8182859, EPI_ISL_8182860, EPI_ISL_8182861, EPI_ISL_8182862, EPI_ISL_8182863, EPI_ISL_8182885, EPI_ISL_8182886, EPI_ISL_8182887, EPI_ISL_8182888, EPI_ISL_8182889, EPI_ISL_8182890 | | | |
| see above | CHRIS HANI BARAGWANATH LABORATORY | National Institute for Communicable Diseases of the National Health Laboratory Service | Amoako DG; Bhiman JN; Everatt J; Ismail A; Kekana D; Mahlangu B; Mnguni A; Mohale T; Ntuli N; Scheepers C; Wolter N |

| | | | |
|---|---|---|---|
| EPI_ISL_2463311 | Charlotte Maxeke | National Institute for Communicable Diseases of the National Health Laboratory Service | Amoako DG; Bhiman JN; Everatt J; Glass A; Sisonke Team; Viana R |
| EPI_ISL_7337463, EPI_ISL_7337464, EPI_ISL_7337465, EPI_ISL_7337466, EPI_ISL_7337467, EPI_ISL_7337468, EPI_ISL_7337469, EPI_ISL_7337470, EPI_ISL_7337471, EPI_ISL_7337473, EPI_ISL_7337474, EPI_ISL_7337475, EPI_ISL_7337476, EPI_ISL_7337477, EPI_ISL_7337478, EPI_ISL_7337479, EPI_ISL_7337480, EPI_ISL_7337481, EPI_ISL_7337482, EPI_ISL_7337483, EPI_ISL_7337484, EPI_ISL_7337485, EPI_ISL_7337486, EPI_ISL_7337487, EPI_ISL_7337488, EPI_ISL_7337489, EPI_ISL_7337490, EPI_ISL_7337495, EPI_ISL_7337496, EPI_ISL_7337497, EPI_ISL_7337498, EPI_ISL_7337499, EPI_ISL_7337500, EPI_ISL_7337501, EPI_ISL_7337502, EPI_ISL_7337503, EPI_ISL_7337504, EPI_ISL_7337505, EPI_ISL_7337506, EPI_ISL_7337507, EPI_ISL_7337508, EPI_ISL_7337509, EPI_ISL_7337510, EPI_ISL_7337511 | | | |
| see above | Charlotte Maxeke Johannesburg Academic Hospital | National Institute for Communicable Diseases of the National Health Laboratory Service | Amoako DG; Bhiman JN; Everatt J; Ismail A; Mahlangu B; Mnguni A; Mohale T; Ntuli N; Scheepers C |
| EPI_ISL_940075, EPI_ISL_940076, EPI_ISL_940077, EPI_ISL_940078, EPI_ISL_940079, EPI_ISL_940080, EPI_ISL_940081, EPI_ISL_940082, EPI_ISL_940083, EPI_ISL_940084, EPI_ISL_940085, EPI_ISL_940086, EPI_ISL_940087, EPI_ISL_940088, EPI_ISL_940089, EPI_ISL_940090, EPI_ISL_940091, EPI_ISL_940092, EPI_ISL_940093, EPI_ISL_940094, EPI_ISL_940095, EPI_ISL_940096, EPI_ISL_940097, EPI_ISL_940098, EPI_ISL_940099, EPI_ISL_940100, EPI_ISL_940101, EPI_ISL_940102, EPI_ISL_940103, EPI_ISL_940104, EPI_ISL_940106, EPI_ISL_940107, EPI_ISL_940109, EPI_ISL_940110, EPI_ISL_940111, EPI_ISL_940112, EPI_ISL_940113, EPI_ISL_940114, EPI_ISL_940115, EPI_ISL_940116, EPI_ISL_940117, EPI_ISL_940118, EPI_ISL_940119, EPI_ISL_940120, EPI_ISL_940121, EPI_ISL_940122, EPI_ISL_940123, EPI_ISL_940124, EPI_ISL_940126, EPI_ISL_940127, EPI_ISL_940128, EPI_ISL_940129, EPI_ISL_940130, EPI_ISL_940131, EPI_ISL_940132, EPI_ISL_940133, EPI_ISL_940134, EPI_ISL_940135, EPI_ISL_940136, EPI_ISL_940137, EPI_ISL_1239267, EPI_ISL_1239268, EPI_ISL_1239269, EPI_ISL_1239270, EPI_ISL_1239271, EPI_ISL_1239272, EPI_ISL_1239273, EPI_ISL_1239274, EPI_ISL_1239275, EPI_ISL_1239276, EPI_ISL_1239277, EPI_ISL_1239278, EPI_ISL_1239279, EPI_ISL_1239280, EPI_ISL_1239281, EPI_ISL_1239282, EPI_ISL_1239283, EPI_ISL_1239284, EPI_ISL_1239285, EPI_ISL_1239287, EPI_ISL_1239288, EPI_ISL_1239289, EPI_ISL_1239290, EPI_ISL_1239291, EPI_ISL_1239292, EPI_ISL_1239293, EPI_ISL_1239294, EPI_ISL_1239295, EPI_ISL_1239296, EPI_ISL_1239297, EPI_ISL_1239298, EPI_ISL_1239299, EPI_ISL_1239300, EPI_ISL_1239301, EPI_ISL_1239302, EPI_ISL_1239303, EPI_ISL_1239304, EPI_ISL_1239305, EPI_ISL_1239306, EPI_ISL_1239307, EPI_ISL_1239308, EPI_ISL_1239309, EPI_ISL_1239310, EPI_ISL_1239311, EPI_ISL_1239312, EPI_ISL_1239313, EPI_ISL_1239314, EPI_ISL_1239315, EPI_ISL_1239316, EPI_ISL_1239317, EPI_ISL_1239318, EPI_ISL_1239319, EPI_ISL_1239320, EPI_ISL_1239321, EPI_ISL_1239322, EPI_ISL_1239323, EPI_ISL_1239324, EPI_ISL_1239325, EPI_ISL_1239326, EPI_ISL_1239328, EPI_ISL_1239329 | | | |
| see above | Charlotte Maxeke Johannesburg Academic Hospital, National Health Laboratory Services, Gauteng, South Africa | National Institute for Communicable Diseases of the National Health Laboratory Service | Allam M; Amoako DG; Bhiman JN; Ismail A; Mahlangu B; Mohale T; Ntuli N; Scheepers C |
| EPI_ISL_4474433, EPI_ISL_4474434, EPI_ISL_4474435, EPI_ISL_4474436, EPI_ISL_4474437, EPI_ISL_4474438, EPI_ISL_4474439, EPI_ISL_4474440, EPI_ISL_4474441, EPI_ISL_4474442, EPI_ISL_4474443 | | | |
| see above | Charlotte Maxeke Johannesburg hospital | CERI, Centre for Epidemic Response and Innovation | Emmanuel S; Florette Treurnicht; Giandhari J; Kathleen Subramoney; Naidoo Yeshnee; Pillay S; Tegally H; Tshabuila Derek; Wilkinson E; Yajna Ramphal; de Oliveira T |
| EPI_ISL_3118689, EPI_ISL_3118690, EPI_ISL_3118691, EPI_ISL_3118692, EPI_ISL_3118693, EPI_ISL_3118694, EPI_ISL_3118695, EPI_ISL_3118696 | | | |
| see above | Chris Hani Baragwanath Academic Hospital POC laboratory | KRISP, KZn Research Innovation and Sequencing Platform | Brown MJ; Emmanuel SJ; Giandhari J; Lessells R; Omar S; Pillay S; Tegally H; Wadula J; Wilkinson E; Yajna R; de Oliveira T |
| EPI_ISL_1239330, EPI_ISL_1239331, EPI_ISL_1239332, EPI_ISL_1239333, EPI_ISL_1239334, EPI_ISL_1239335, EPI_ISL_1239336, EPI_ISL_1239337, EPI_ISL_1239338, EPI_ISL_1239339, EPI_ISL_1239340, EPI_ISL_1239341, EPI_ISL_1239342, EPI_ISL_1239343, EPI_ISL_1239344, EPI_ISL_1239345, EPI_ISL_1239346, EPI_ISL_1239347, EPI_ISL_1239348, EPI_ISL_1239349, EPI_ISL_1239350, EPI_ISL_1239351, EPI_ISL_1239352, EPI_ISL_1239353, EPI_ISL_1239354, EPI_ISL_1239355, EPI_ISL_1239356, EPI_ISL_1239357, EPI_ISL_1239358, EPI_ISL_1239360 | | | |
| see above | Chris Hani Baragwanath Hospital, National Health Laboratory Services, Gauteng, South Africa | National Institute for Communicable Diseases of the National Health Laboratory Service | Amoako DG; Bhiman JN; Ismail A; Mahlangu B; Mohale T; Ntuli N; Scheepers C |
| EPI_ISL_5917986, EPI_ISL_5917992, EPI_ISL_5918009, EPI_ISL_7337512, EPI_ISL_7337513, EPI_ISL_7337514, EPI_ISL_7337515, EPI_ISL_7337516, EPI_ISL_7337517, EPI_ISL_7337518, EPI_ISL_7337519, EPI_ISL_7337520, EPI_ISL_7337521, EPI_ISL_7337522, EPI_ISL_7337523, EPI_ISL_7337524, EPI_ISL_7337525, EPI_ISL_7337527, EPI_ISL_7337528, EPI_ISL_7337529, EPI_ISL_7337530, EPI_ISL_7337531, EPI_ISL_7337532, EPI_ISL_7337533, EPI_ISL_7337534, EPI_ISL_7337535, EPI_ISL_7337536, EPI_ISL_7337537, EPI_ISL_7337538, EPI_ISL_7337539, EPI_ISL_7337540, EPI_ISL_7337541, EPI_ISL_7337542, EPI_ISL_7337543, EPI_ISL_7337544, EPI_ISL_7337545, EPI_ISL_7337546, EPI_ISL_7337547, EPI_ISL_7337548, EPI_ISL_7337549, EPI_ISL_7337550, EPI_ISL_7337551, EPI_ISL_7337553, EPI_ISL_7337558, EPI_ISL_7337559, EPI_ISL_7337560, EPI_ISL_7337561, EPI_ISL_7337562, EPI_ISL_7337563, EPI_ISL_7337564, EPI_ISL_7456412 | | | |
| see above | Chris Hani Baragwanath Laboratory | National Institute for Communicable Diseases of the National Health Laboratory Service | Amoako DG; Bhiman JN; Everatt J; Ismail A; Mahlangu B; Mnguni A; Mohale T; Ntuli N; Scheepers C; Wolter N |
| EPI_ISL_3342336, EPI_ISL_3342338, EPI_ISL_3342339, EPI_ISL_3342340, EPI_ISL_3342342, EPI_ISL_3342344, EPI_ISL_3342346, EPI_ISL_3342347, EPI_ISL_3342348, EPI_ISL_3411472 | | | |
| see above | DGM Laboratory | National Institute for Communicable Diseases of the National Health Laboratory Service | Amoako DG; Bhiman JN; Everatt J; Ismail A; Mahlangu B; Mnguni A; Mohale T; Ntuli N; Scheepers C |
| EPI_ISL_2153483, EPI_ISL_2153485, EPI_ISL_2153486, EPI_ISL_2153487, EPI_ISL_2153488, EPI_ISL_2153489, EPI_ISL_2153490, EPI_ISL_2153491, EPI_ISL_2153493, EPI_ISL_2153494, EPI_ISL_2153495, EPI_ISL_2153496, EPI_ISL_2153497, EPI_ISL_2153498, EPI_ISL_2153499, EPI_ISL_2153500, EPI_ISL_2153501, EPI_ISL_2153502, EPI_ISL_2153503, EPI_ISL_2153504, EPI_ISL_2153505, EPI_ISL_2153506, EPI_ISL_2153507, EPI_ISL_2153508, EPI_ISL_2153509, EPI_ISL_2153510, EPI_ISL_2153511, EPI_ISL_2153512, EPI_ISL_2153513, EPI_ISL_2153514, EPI_ISL_2153515, EPI_ISL_2285251, EPI_ISL_2285252, EPI_ISL_2582809, EPI_ISL_2662511, EPI_ISL_2662512, EPI_ISL_2662513, EPI_ISL_2662514, EPI_ISL_2662515, EPI_ISL_2662516, EPI_ISL_2662517, EPI_ISL_2662518, EPI_ISL_2662519, EPI_ISL_2662520, EPI_ISL_2662521, EPI_ISL_2662522, EPI_ISL_2662523, EPI_ISL_2662524, EPI_ISL_2662525, EPI_ISL_2662526, EPI_ISL_2662527, EPI_ISL_2662528, EPI_ISL_2662538, EPI_ISL_2662539, EPI_ISL_2662540, EPI_ISL_2662541, EPI_ISL_2662542, EPI_ISL_2662543, EPI_ISL_2662544, EPI_ISL_2662545, EPI_ISL_2662546, EPI_ISL_2662547, EPI_ISL_2662548, EPI_ISL_2662549, EPI_ISL_2662551, EPI_ISL_2662552 | | | |
| see above | DR GEORGE MUKHARI LABORATORY | National Institute for Communicable Diseases of the National Health Laboratory Service | Amoako DG; Bhiman JN; Everatt J; Ismail A; Mahlangu B; Mnguni A; Mohale T; Ntuli N; Scheepers C |
| EPI_ISL_2382663 | Dr George Mukhari Academic Hospital | National Institute for Communicable Diseases of the National Health Laboratory Service | Amoako DG; Bhiman JN; Ismail A; Mahlangu B; Mohale T; Ntuli N; Scheepers C |
| EPI_ISL_2695588, EPI_ISL_2695589, EPI_ISL_2695590, EPI_ISL_2695591, EPI_ISL_2695592, EPI_ISL_2695593, EPI_ISL_2695594, EPI_ISL_2695595, EPI_ISL_2695596, EPI_ISL_2695597, EPI_ISL_2695598, EPI_ISL_2695599, EPI_ISL_2695600, EPI_ISL_2695601, EPI_ISL_2695602, EPI_ISL_2695603, EPI_ISL_2695604, EPI_ISL_2695605, EPI_ISL_2695606, EPI_ISL_2695727, EPI_ISL_2695728, EPI_ISL_2695729, EPI_ISL_2695730, EPI_ISL_2695731, EPI_ISL_2695732, EPI_ISL_2695733, EPI_ISL_2695734, EPI_ISL_2695735, EPI_ISL_2695736, EPI_ISL_2695737, EPI_ISL_2695738, EPI_ISL_2695739, EPI_ISL_2695740, EPI_ISL_2695741, EPI_ISL_2695742, EPI_ISL_2695743, EPI_ISL_2695744, EPI_ISL_2695745, EPI_ISL_2695746, EPI_ISL_2695747, EPI_ISL_2695748, EPI_ISL_2695749, EPI_ISL_2695750, EPI_ISL_2695751, EPI_ISL_2709863, EPI_ISL_2709864, EPI_ISL_2709872, EPI_ISL_2709873, EPI_ISL_2709874, EPI_ISL_2709875, EPI_ISL_2709876, EPI_ISL_2709877, EPI_ISL_2709878, EPI_ISL_2709885, EPI_ISL_2709886 | | | |
| see above | HELEN JOSEPH | National Institute for Communicable Diseases of the National Health Laboratory Service | Amoako DG; Bhiman JN; Everatt J; Ismail A; Mahlangu B; Mnguni A; Mohale T; Ntuli N; Scheepers C |
| EPI_ISL_2827072, EPI_ISL_2827073, EPI_ISL_2827074, EPI_ISL_2827075, EPI_ISL_2827076, EPI_ISL_2827077, EPI_ISL_2827078, EPI_ISL_2827079, EPI_ISL_2827080, EPI_ISL_2827081, EPI_ISL_2827082, EPI_ISL_2827083, EPI_ISL_2827084, EPI_ISL_2827085, EPI_ISL_2827086, EPI_ISL_2827087, EPI_ISL_2827088, EPI_ISL_2827089, EPI_ISL_2827090, EPI_ISL_2827091, EPI_ISL_2827092, EPI_ISL_2827093, EPI_ISL_2827094, EPI_ISL_2827095, EPI_ISL_2827096, EPI_ISL_2827097, EPI_ISL_2827098, EPI_ISL_2827099, EPI_ISL_2827100, EPI_ISL_2827101, EPI_ISL_2827102, EPI_ISL_2827103, EPI_ISL_2827104, EPI_ISL_2827105, EPI_ISL_2827106, EPI_ISL_2827107, EPI_ISL_2827108, EPI_ISL_2827109, EPI_ISL_2827110, EPI_ISL_2827111, EPI_ISL_2827113, EPI_ISL_2827114, EPI_ISL_2827115, EPI_ISL_2827116, EPI_ISL_2827117, EPI_ISL_2827118, EPI_ISL_2827119, EPI_ISL_2827120, EPI_ISL_2827121, EPI_ISL_2827122, EPI_ISL_2827123, EPI_ISL_2827124, EPI_ISL_2827125, EPI_ISL_2827126, EPI_ISL_2827127, EPI_ISL_2827128, EPI_ISL_2827129, EPI_ISL_2827130, EPI_ISL_2827131, EPI_ISL_2827132, EPI_ISL_2895772, EPI_ISL_2895773, EPI_ISL_2895774, EPI_ISL_2895775, EPI_ISL_2895776, EPI_ISL_2895777, EPI_ISL_2895778, EPI_ISL_2895779, EPI_ISL_2895780, EPI_ISL_2895781, EPI_ISL_2895782, EPI_ISL_2895783, EPI_ISL_2895784, EPI_ISL_2895785, EPI_ISL_2895786, EPI_ISL_2895787, EPI_ISL_2895788, EPI_ISL_2895789, EPI_ISL_2895790, EPI_ISL_2895791, EPI_ISL_2895792, EPI_ISL_2895793, EPI_ISL_2895794, EPI_ISL_2895795, EPI_ISL_2895796, EPI_ISL_2895797, EPI_ISL_2942284, EPI_ISL_2942285, EPI_ISL_2942286, EPI_ISL_2942287, EPI_ISL_2964647, EPI_ISL_2984812, EPI_ISL_2984813, EPI_ISL_2984814, EPI_ISL_2984815, EPI_ISL_2984816, EPI_ISL_2984817, EPI_ISL_2984818, EPI_ISL_2984819, EPI_ISL_2984820, EPI_ISL_2984821, EPI_ISL_2984822, EPI_ISL_2984823, EPI_ISL_2984824, EPI_ISL_2984825, EPI_ISL_2984826, EPI_ISL_2984827, EPI_ISL_2984828, EPI_ISL_2984829, EPI_ISL_2984830, EPI_ISL_2984831, EPI_ISL_2984832, EPI_ISL_2984833, EPI_ISL_2984834, EPI_ISL_2984835, EPI_ISL_2984893, EPI_ISL_2984894, EPI_ISL_2984895, EPI_ISL_2984896, EPI_ISL_2984897, EPI_ISL_2984898, EPI_ISL_2984899, EPI_ISL_2984900, EPI_ISL_2984901, EPI_ISL_2984902, EPI_ISL_2984903, EPI_ISL_2984904, EPI_ISL_2984905, EPI_ISL_2984906, EPI_ISL_2984907, EPI_ISL_2984908, EPI_ISL_2984909, EPI_ISL_2984910, EPI_ISL_2987234, EPI_ISL_2988404, EPI_ISL_2988405, EPI_ISL_2988408, EPI_ISL_2988409, EPI_ISL_2988878, EPI_ISL_3024575, EPI_ISL_3024576, EPI_ISL_3024577, EPI_ISL_3024578, EPI_ISL_3024579, EPI_ISL_3024580, EPI_ISL_3024581, EPI_ISL_3024582, EPI_ISL_3024583, EPI_ISL_3024584, EPI_ISL_3024585, EPI_ISL_3024586, EPI_ISL_3024587, EPI_ISL_3024588, EPI_ISL_3024589, EPI_ISL_3024590, EPI_ISL_3024591, EPI_ISL_3024592, EPI_ISL_3024593, EPI_ISL_3024594, EPI_ISL_3024595, EPI_ISL_3024596, EPI_ISL_3024597, EPI_ISL_3024598, EPI_ISL_3024599, EPI_ISL_3024600, EPI_ISL_3024601, EPI_ISL_3024602, EPI_ISL_3024603, EPI_ISL_3024604, EPI_ISL_3024605, EPI_ISL_3024606, EPI_ISL_3024607, EPI_ISL_3024608, EPI_ISL_3024609, EPI_ISL_3024610, EPI_ISL_3024611, EPI_ISL_3024612, EPI_ISL_3024613, EPI_ISL_3024614, EPI_ISL_3024615, EPI_ISL_3024616, EPI_ISL_3024617, EPI_ISL_3024618, EPI_ISL_3024619, EPI_ISL_3024620, EPI_ISL_3024621, EPI_ISL_3024622, EPI_ISL_3024623, EPI_ISL_3024624, EPI_ISL_3024625, EPI_ISL_3024626, EPI_ISL_3030436, EPI_ISL_3030437, EPI_ISL_3030438, EPI_ISL_3032185, EPI_ISL_3451530, EPI_ISL_3451531, EPI_ISL_3838401, EPI_ISL_3838402, EPI_ISL_3838403, EPI_ISL_3838404, EPI_ISL_3838405, EPI_ISL_3838406, EPI_ISL_3838407, EPI_ISL_3838408, EPI_ISL_3838409, EPI_ISL_3838410, EPI_ISL_3838411, EPI_ISL_3838412, EPI_ISL_3838413, EPI_ISL_3838414, EPI_ISL_3838415, EPI_ISL_3838416, EPI_ISL_3838417, EPI_ISL_3838418, EPI_ISL_3838419, EPI_ISL_3838420, EPI_ISL_3838421, EPI_ISL_3838422, EPI_ISL_3838423, EPI_ISL_3838424, EPI_ISL_3838425, EPI_ISL_3838426, EPI_ISL_4253615, EPI_ISL_4253616, EPI_ISL_4253617, EPI_ISL_4253618, EPI_ISL_4253619, EPI_ISL_4253620, EPI_ISL_4253621, EPI_ISL_4253622, EPI_ISL_4253623, EPI_ISL_4253624, EPI_ISL_4253625, EPI_ISL_4253626, EPI_ISL_4253627, EPI_ISL_4253628, EPI_ISL_4253629, EPI_ISL_4253630, EPI_ISL_4253631, EPI_ISL_4253632, EPI_ISL_4253633, EPI_ISL_4253634, EPI_ISL_4253635, EPI_ISL_4253636, EPI_ISL_4253637, EPI_ISL_4253638, EPI_ISL_4253639, EPI_ISL_4253640, EPI_ISL_4253641, EPI_ISL_4253642, EPI_ISL_4253643, EPI_ISL_4253644, EPI_ISL_4253645, EPI_ISL_4253646, EPI_ISL_4651672, EPI_ISL_4651733, EPI_ISL_4651850, EPI_ISL_4651941, EPI_ISL_4651945, EPI_ISL_4651959, EPI_ISL_4651962, EPI_ISL_4651970, EPI_ISL_5416275, EPI_ISL_5416276, EPI_ISL_5416282, EPI_ISL_5416288, EPI_ISL_5416292, EPI_ISL_5416312, EPI_ISL_5416321, EPI_ISL_5416323, EPI_ISL_5416324, EPI_ISL_5416355, EPI_ISL_5416383, EPI_ISL_5416465, EPI_ISL_5416466, EPI_ISL_5416467, EPI_ISL_5416468, EPI_ISL_5416469, EPI_ISL_5416470, EPI_ISL_5416471, EPI_ISL_5416472, EPI_ISL_6900028, EPI_ISL_6900095, EPI_ISL_6900139, EPI_ISL_6900140, EPI_ISL_6900141, EPI_ISL_6900142, EPI_ISL_6900143, EPI_ISL_6906407, EPI_ISL_7740761, EPI_ISL_7740762, EPI_ISL_7740765, EPI_ISL_7740768, EPI_ISL_7740769, EPI_ISL_7740770, EPI_ISL_7740771, EPI_ISL_7740772, EPI_ISL_7740773, EPI_ISL_7740774, EPI_ISL_7740775, EPI_ISL_7740776, EPI_ISL_7740777, EPI_ISL_7740778, EPI_ISL_7740779, EPI_ISL_7740780, EPI_ISL_7740781, EPI_ISL_7740782, EPI_ISL_7740783, EPI_ISL_7740784, EPI_ISL_7740785, EPI_ISL_7740786, EPI_ISL_7740787, EPI_ISL_7740788, EPI_ISL_7740789, EPI_ISL_7740790, EPI_ISL_7740791, EPI_ISL_7740792, EPI_ISL_7740793, EPI_ISL_7740794, EPI_ISL_7740795, EPI_ISL_7740796, EPI_ISL_7740797, EPI_ISL_7740801, EPI_ISL_7740802, EPI_ISL_7740803, EPI_ISL_7740804, EPI_ISL_7740805, EPI_ISL_7740806, EPI_ISL_7740807, EPI_ISL_7740808, EPI_ISL_7740809, EPI_ISL_7740810, EPI_ISL_7740811, EPI_ISL_7740817, EPI_ISL_7740818, EPI_ISL_7740819, EPI_ISL_7740820, EPI_ISL_7740821, EPI_ISL_7740822, EPI_ISL_7740823, EPI_ISL_7740824, EPI_ISL_7740825, EPI_ISL_7740826, EPI_ISL_7740827, EPI_ISL_7740828, EPI_ISL_7740829, EPI_ISL_7740830, EPI_ISL_7740831, EPI_ISL_7740832, EPI_ISL_7740833, EPI_ISL_7740834, EPI_ISL_7740835, EPI_ISL_7740836, EPI_ISL_7740837, EPI_ISL_7740838, EPI_ISL_7740839, EPI_ISL_7740840, EPI_ISL_7740841, EPI_ISL_7740842, EPI_ISL_7740843, EPI_ISL_7740844, EPI_ISL_7740845, EPI_ISL_7740846, EPI_ISL_7740847, EPI_ISL_7740848, EPI_ISL_7740849, EPI_ISL_7740850, EPI_ISL_7740851, EPI_ISL_7740852, EPI_ISL_7740853, EPI_ISL_7740862, EPI_ISL_7740863, EPI_ISL_7740864, EPI_ISL_7740867, EPI_ISL_7740868, EPI_ISL_7740870, EPI_ISL_7740871, EPI_ISL_7740872, EPI_ISL_7740873, EPI_ISL_7740874, EPI_ISL_7740876, EPI_ISL_7740877, EPI_ISL_7740879, EPI_ISL_7740880, EPI_ISL_7740881, EPI_ISL_7740882, EPI_ISL_7740884, EPI_ISL_7740886, EPI_ISL_7740888, EPI_ISL_7740891, EPI_ISL_7740892, EPI_ISL_7740894, EPI_ISL_7740895, EPI_ISL_7740896, EPI_ISL_7740898, EPI_ISL_7740899 | | | |
| see above | HELEN JOSEPH LABORATORY | National Institute for Communicable Diseases of the National Health Laboratory Service | Amoako DG; Bhiman JN; Everatt J; Ismail A; Mahlangu B; Mnguni A; Mohale T; Ntuli N; Scheepers C; Wolter N |
| EPI_ISL_2271973, EPI_ISL_2271974, EPI_ISL_2271983, EPI_ISL_2271995, EPI_ISL_2271996, EPI_ISL_2271997, EPI_ISL_2271998, EPI_ISL_2272001, EPI_ISL_2272002, EPI_ISL_2272003, EPI_ISL_2272005, EPI_ISL_2272006, EPI_ISL_2272017, EPI_ISL_2272022, EPI_ISL_5510433 | | | |
| see above | Helen Joseph | National Institute for Communicable Diseases of the National Health Laboratory Service | Amoako DG; Bhiman JN; Everatt J; Ismail A; Mahlangu B; Mnguni A; Mohale T; Ntuli N; Scheepers C |
| EPI_ISL_3342734, EPI_ISL_3342735, EPI_ISL_4891523, EPI_ISL_4891634, EPI_ISL_4891635, EPI_ISL_4891643, EPI_ISL_4891658, EPI_ISL_4891662, EPI_ISL_4891779, EPI_ISL_4891780, EPI_ISL_4891781, EPI_ISL_4891782, EPI_ISL_4891783, EPI_ISL_5917851, EPI_ISL_5917869, EPI_ISL_5917880, EPI_ISL_5917886, EPI_ISL_5917956, EPI_ISL_5917991, EPI_ISL_5918006, EPI_ISL_5918016, EPI_ISL_5918018, EPI_ISL_5918025, EPI_ISL_5918034, EPI_ISL_5918065, EPI_ISL_5918093, EPI_ISL_5918155, EPI_ISL_5918156, EPI_ISL_5918157, EPI_ISL_5918158, EPI_ISL_7337440, EPI_ISL_7337441, EPI_ISL_7337442, EPI_ISL_7337443, EPI_ISL_7337444, EPI_ISL_7337445, EPI_ISL_7337446, EPI_ISL_7337447, EPI_ISL_7337448, EPI_ISL_7337449, EPI_ISL_7337450, EPI_ISL_7337451, EPI_ISL_7337452, EPI_ISL_7337453, EPI_ISL_7337454, EPI_ISL_7337455, EPI_ISL_7337456, EPI_ISL_7337457, EPI_ISL_7337458, EPI_ISL_7337459, EPI_ISL_7337460, EPI_ISL_7337461, EPI_ISL_7337462, EPI_ISL_7456431 | | | |
| see above | Helen Joseph Laboratory | National Institute for Communicable Diseases of the National Health Laboratory Service | Amoako DG; Bhiman JN; Everatt J; Ismail A; Mahlangu B; Mnguni A; Mohale T; Ntuli N; Scheepers C; Wolter N |
| EPI_ISL_3859862 | Instituto Nacional de Investigação em Saúde | CERI, Centre for Epidemic Response and Innvoation, Stellenbosch University and KRISP, KZN Research Innovation and Sequencing Platform, UKZN. | Afonso P; David K; Emmanuel SJ; Freitas RH; Giandhari J; Inglês L; Lutucuta S; Miranda J; Morais J; Mufinda M; Naidoo Y; Neto Z; Paulo A Carralero RR Paixão JP; Pereira A; Pillay S; Tegally H; Wilkinson E; de Oliveira T |
| EPI_ISL_2271953, EPI_ISL_2271955, EPI_ISL_2271956, EPI_ISL_2271957, EPI_ISL_2271964, EPI_ISL_2271965, EPI_ISL_2271971, EPI_ISL_2271977, EPI_ISL_2271978, EPI_ISL_2271981, EPI_ISL_2271984, EPI_ISL_2271986, EPI_ISL_2271987, EPI_ISL_2271988, EPI_ISL_2271989, EPI_ISL_2271994, EPI_ISL_2272000, EPI_ISL_2272004, EPI_ISL_2272018 | | | |
| see above | Joubert Clinic | National Institute for Communicable Diseases of the National Health Laboratory Service | Amoako DG; Bhiman JN; Ismail A; Mahlangu B; Mohale T; Ntuli N; Scheepers C |
| EPI_ISL_3342332, EPI_ISL_3342333, EPI_ISL_3342334, EPI_ISL_3342335, EPI_ISL_3342337, EPI_ISL_3342341, EPI_ISL_3342343, EPI_ISL_3342345, EPI_ISL_3342349 | | | |
| see above | Jubilee laboratory | National Institute for Communicable Diseases of the National Health Laboratory Service | Amoako DG; Bhiman JN; Everatt J; Ismail A; Mahlangu B; Mnguni A; Mohale T; Ntuli N; Scheepers C |
| EPI_ISL_2382224, EPI_ISL_3024573, EPI_ISL_3024574, EPI_ISL_8182807 | KIMBERLEY LABORATORY | National Institute for Communicable Diseases of the National Health Laboratory Service | Amoako DG; Bhiman JN; Everatt J; Ismail A; Kekana D; Mahlangu B; Mnguni A; Mohale T; Ntuli N; Scheepers C; Wolter N |
| EPI_ISL_5510405 | KOOKRUS PHC-B | National Institute for Communicable Diseases of the National Health Laboratory Service | Amoako DG; Bhiman JN; Everatt J; Ismail A; Mahlangu B; Mnguni A; Mohale T; Ntuli N; Scheepers C |
| EPI_ISL_2694614, EPI_ISL_2694615, EPI_ISL_2695512, EPI_ISL_2695513, EPI_ISL_2695523, EPI_ISL_2695524, EPI_ISL_5510279, EPI_ISL_5510281, EPI_ISL_5510340, EPI_ISL_5510343, EPI_ISL_5510488 | | | |
| see above | KOPANONG | National Institute for Communicable Diseases of the National Health Laboratory Service | Amoako DG; Bhiman JN; Everatt J; Ismail A; Mahlangu B; Mnguni A; Mohale T; Ntuli N; Scheepers C |
| EPI_ISL_3101489, EPI_ISL_3101490, EPI_ISL_3101491, EPI_ISL_3101492, EPI_ISL_3101493, EPI_ISL_3101494, EPI_ISL_3101495, EPI_ISL_3101496, EPI_ISL_3101497, EPI_ISL_3101498, EPI_ISL_3101499, EPI_ISL_3101500, EPI_ISL_3101501, EPI_ISL_3101502, EPI_ISL_3101503, EPI_ISL_3101504, EPI_ISL_3101505, EPI_ISL_3101506, EPI_ISL_3101507, EPI_ISL_3101508, EPI_ISL_3101509, EPI_ISL_3101510, EPI_ISL_3101511, EPI_ISL_3101512, EPI_ISL_3101513, EPI_ISL_3101514, EPI_ISL_3101515, EPI_ISL_3101516, EPI_ISL_3101517, EPI_ISL_3101518, EPI_ISL_3101519, EPI_ISL_3101520, EPI_ISL_3101521, EPI_ISL_3101522, EPI_ISL_3101523, EPI_ISL_3101524, EPI_ISL_3101525, EPI_ISL_3101526, EPI_ISL_3101527, EPI_ISL_3101528, EPI_ISL_3101529, EPI_ISL_3101530, EPI_ISL_3101531, EPI_ISL_3101532, | | | |

| | | | |
|---|---|---|---|
| EPI_ISL_3101533, EPI_ISL_3101534, EPI_ISL_3101633, EPI_ISL_3101634, EPI_ISL_3149299, EPI_ISL_3149300, EPI_ISL_3149301, EPI_ISL_3149302, EPI_ISL_3149304 | | | |
| see above | KOPANONG HOSPITAL | National Institute for Communicable Diseases of the National Health Laboratory Service | Amoako DG; Bhiman JN; Everatt J; Ismail A; Mahlangu B; Mnguni A; Mohale T; Ntuli N; Scheepers C |
| EPI_ISL_2382260, EPI_ISL_2382261, EPI_ISL_2382262, EPI_ISL_2382263, EPI_ISL_2382264, EPI_ISL_2382265, EPI_ISL_2382266, EPI_ISL_2582987, EPI_ISL_2582990, EPI_ISL_2582993, EPI_ISL_2582996, EPI_ISL_2582999, EPI_ISL_2583001, EPI_ISL_2583005, EPI_ISL_2662662, EPI_ISL_2662663, EPI_ISL_2662664, EPI_ISL_2695759, EPI_ISL_2695760, EPI_ISL_2695761, EPI_ISL_2895766, EPI_ISL_2895767, EPI_ISL_2895768, EPI_ISL_2895769, EPI_ISL_2895770, EPI_ISL_2895771, EPI_ISL_2895799, EPI_ISL_2895800, EPI_ISL_2942283, EPI_ISL_3024643, EPI_ISL_3024644, EPI_ISL_3024645, EPI_ISL_3236955, EPI_ISL_3236956, EPI_ISL_3237104, EPI_ISL_3237105, EPI_ISL_3237106, EPI_ISL_3237254, EPI_ISL_3342222, EPI_ISL_3342223, EPI_ISL_3342224, EPI_ISL_3342225, EPI_ISL_3342226, EPI_ISL_3342227, EPI_ISL_3342228, EPI_ISL_3342229, EPI_ISL_3342230, EPI_ISL_3342231, EPI_ISL_3342232, EPI_ISL_3342233, EPI_ISL_3342234, EPI_ISL_3342235, EPI_ISL_3342236, EPI_ISL_3342237, EPI_ISL_3342238, EPI_ISL_3342239, EPI_ISL_3342240, EPI_ISL_3342241, EPI_ISL_3342242, EPI_ISL_3342243, EPI_ISL_3342244, EPI_ISL_3342245, EPI_ISL_3342246, EPI_ISL_3342248, EPI_ISL_3342249, EPI_ISL_3342250, EPI_ISL_3342251, EPI_ISL_3342252, EPI_ISL_3342253, EPI_ISL_3342254, EPI_ISL_3342256, EPI_ISL_3342422, EPI_ISL_3411463, EPI_ISL_3411466, EPI_ISL_3411467, EPI_ISL_3411469, EPI_ISL_3411770, EPI_ISL_3411810, EPI_ISL_3411811, EPI_ISL_3411812, EPI_ISL_3411813, EPI_ISL_3411814, EPI_ISL_3411815, EPI_ISL_3411816, EPI_ISL_3451255, EPI_ISL_3451256, EPI_ISL_3451491, EPI_ISL_3451492, EPI_ISL_3451493, EPI_ISL_3451494, EPI_ISL_3451495, EPI_ISL_3451496, EPI_ISL_3451497, EPI_ISL_3451498, EPI_ISL_3451499, EPI_ISL_3451500, EPI_ISL_3451501, EPI_ISL_3451502, EPI_ISL_3451503, EPI_ISL_3643814, EPI_ISL_3643821, EPI_ISL_3643830, EPI_ISL_3643833, EPI_ISL_3643840, EPI_ISL_3643846, EPI_ISL_3643870, EPI_ISL_3643876, EPI_ISL_3643877, EPI_ISL_3643887, EPI_ISL_3643889, EPI_ISL_3643900, EPI_ISL_3643901, EPI_ISL_3643910, EPI_ISL_3643911, EPI_ISL_3643912, EPI_ISL_3643914, EPI_ISL_3643917, EPI_ISL_3643924, EPI_ISL_3643926, EPI_ISL_3643932, EPI_ISL_3643933, EPI_ISL_3643934, EPI_ISL_3643950, EPI_ISL_3643960, EPI_ISL_3643961, EPI_ISL_3643963, EPI_ISL_3643966, EPI_ISL_3643969, EPI_ISL_3643974, EPI_ISL_3643980, EPI_ISL_3643981, EPI_ISL_3643982, EPI_ISL_3643986, EPI_ISL_3643987, EPI_ISL_3644019, EPI_ISL_3644020, EPI_ISL_3644021, EPI_ISL_3644022, EPI_ISL_3644023, EPI_ISL_3644024, EPI_ISL_3644025, EPI_ISL_3644026, EPI_ISL_3644027, EPI_ISL_3644028, EPI_ISL_3644029, EPI_ISL_5416274, EPI_ISL_5416342, EPI_ISL_6202362, EPI_ISL_6202425, EPI_ISL_6202743, EPI_ISL_6202820, EPI_ISL_6327770, EPI_ISL_6327771, EPI_ISL_6327848, EPI_ISL_6900144, EPI_ISL_6900145, EPI_ISL_6900146, EPI_ISL_6900147, EPI_ISL_6900148, EPI_ISL_6900149 | | | |
| see above | KOPANONG LABORATORY | National Institute for Communicable Diseases of the National Health Laboratory Service | Amoako DG; Bhiman JN; Everatt J; Ismail A; Mahlangu B; Mnguni A; Mohale T; Ntuli N; Scheepers C |
| EPI_ISL_5510467, EPI_ISL_5510468, EPI_ISL_5510482 | Kathleen Subramoney | National Institute for Communicable Diseases of the National Health Laboratory Service | Amoako DG; Bhiman JN; Everatt J; Ismail A; Mahlangu B; Mnguni A; Mohale T; Ntuli N; Scheepers C |
| EPI_ISL_5510359, EPI_ISL_5510402, EPI_ISL_5510438, EPI_ISL_5510441, EPI_ISL_5510465 | Kopanong | National Institute for Communicable Diseases of the National Health Laboratory Service | Amoako DG; Bhiman JN; Everatt J; Ismail A; Mahlangu B; Mnguni A; Mohale T; Ntuli N; Scheepers C |
| EPI_ISL_5917867, EPI_ISL_5917931, EPI_ISL_5917976, EPI_ISL_5917993, EPI_ISL_5918007, EPI_ISL_5918012, EPI_ISL_5918148, EPI_ISL_7337381, EPI_ISL_7337385, EPI_ISL_7337386, EPI_ISL_7337387, EPI_ISL_7337388, EPI_ISL_7456413, EPI_ISL_7456414, EPI_ISL_7456415, EPI_ISL_7456416, EPI_ISL_7456418, EPI_ISL_7456429 | | | |
| see above | Kopanong Laboratory | National Institute for Communicable Diseases of the National Health Laboratory Service | Amoako DG; Bhiman JN; Everatt J; Ismail A; Mahlangu B; Mnguni A; Mohale T; Ntuli N; Scheepers C; Wolter N |
| EPI_ISL_2463301, EPI_ISL_2463302, EPI_ISL_2463303, EPI_ISL_2463304, EPI_ISL_2463305, EPI_ISL_2463306, EPI_ISL_2463307, EPI_ISL_2463308, EPI_ISL_2463309, EPI_ISL_2463310, EPI_ISL_3236957, EPI_ISL_3236989, EPI_ISL_3236990, EPI_ISL_3236991, EPI_ISL_3236992, EPI_ISL_3237107, EPI_ISL_3237108, EPI_ISL_3237109, EPI_ISL_3237110, EPI_ISL_3237111, EPI_ISL_3237112, EPI_ISL_3237113, EPI_ISL_3237167, EPI_ISL_3237168, EPI_ISL_3237169, EPI_ISL_3237170, EPI_ISL_3237171, EPI_ISL_3237172, EPI_ISL_3237258, EPI_ISL_3643990, EPI_ISL_6647956, EPI_ISL_6647957, EPI_ISL_6647958, EPI_ISL_6647959, EPI_ISL_6647960, EPI_ISL_6647961, EPI_ISL_6647962, EPI_ISL_6698792, EPI_ISL_6704863, EPI_ISL_6704864, EPI_ISL_6704865, EPI_ISL_6704866, EPI_ISL_6704867, EPI_ISL_6704868, EPI_ISL_6704869, EPI_ISL_6704870, EPI_ISL_6704871, EPI_ISL_6704872, EPI_ISL_6704873, EPI_ISL_6704874, EPI_ISL_6704875, EPI_ISL_6704876, EPI_ISL_7701115, EPI_ISL_7701117, EPI_ISL_7701120, EPI_ISL_7701122, EPI_ISL_7701125, EPI_ISL_7701127, EPI_ISL_7701129, EPI_ISL_7701131, EPI_ISL_7701134, EPI_ISL_7701136, EPI_ISL_7701138 | | | |
| see above | LANCET LABORATORY | National Institute for Communicable Diseases of the National Health Laboratory Service | Amoako DG; Bhiman JN; Everatt J; Glass A; Ismail A; Mahlangu B; Mnguni A; Mohale T; Ntuli N; Scheepers C; Sisonke Team; Viana R; Wolter N |
| EPI_ISL_2433705, EPI_ISL_2433706, EPI_ISL_2433707, EPI_ISL_2433708, EPI_ISL_2433709, EPI_ISL_2433710, EPI_ISL_2433711, EPI_ISL_2433713, EPI_ISL_2433716, EPI_ISL_2433717, EPI_ISL_2433719, EPI_ISL_2433720, EPI_ISL_2433721, EPI_ISL_2433722, EPI_ISL_2433723, EPI_ISL_2433724, EPI_ISL_2433850, EPI_ISL_2433851, EPI_ISL_2433852, EPI_ISL_2433853, EPI_ISL_2433854, EPI_ISL_2433855, EPI_ISL_2433856, EPI_ISL_2433857, EPI_ISL_2433858, EPI_ISL_2433859, EPI_ISL_2433860, EPI_ISL_2433864, EPI_ISL_2433865, EPI_ISL_2433866, EPI_ISL_2433870, EPI_ISL_2433871, EPI_ISL_2433872, EPI_ISL_2433873, EPI_ISL_2433874, EPI_ISL_2433875, EPI_ISL_2433876, EPI_ISL_2433877, EPI_ISL_2433878, EPI_ISL_2433879, EPI_ISL_2433880, EPI_ISL_2433881, EPI_ISL_2433882, EPI_ISL_2433883, EPI_ISL_2433884, EPI_ISL_2433885, EPI_ISL_2433886, EPI_ISL_2433887, EPI_ISL_2433888, EPI_ISL_2433889, EPI_ISL_2433890, EPI_ISL_2433891, EPI_ISL_2433892, EPI_ISL_2433893, EPI_ISL_2433895, EPI_ISL_2433896, EPI_ISL_2433897, EPI_ISL_2433899, EPI_ISL_2433900, EPI_ISL_2433901, EPI_ISL_2433903, EPI_ISL_2433904, EPI_ISL_2433905, EPI_ISL_2433909, EPI_ISL_2433910, EPI_ISL_2433911, EPI_ISL_2433912, EPI_ISL_2433913, EPI_ISL_2433914, EPI_ISL_2433915, EPI_ISL_2433916, EPI_ISL_2433918, EPI_ISL_2433919, EPI_ISL_2433920, EPI_ISL_2433922, EPI_ISL_2433923, EPI_ISL_2433924, EPI_ISL_2433925, EPI_ISL_2443995, EPI_ISL_2443996 | | | |
| see above | LANCET, LABORATORIES | National Institute for Communicable Diseases of the National Health Laboratory Service | Amoako DG; Bhiman JN; Ismail A; Mahlangu B; Mohale T; Ntuli N; Scheepers C |
| EPI_ISL_2828731, EPI_ISL_2828732, EPI_ISL_2828733, EPI_ISL_5510283, EPI_ISL_5510348 | Lancet | National Institute for Communicable Diseases of the National Health Laboratory Service | Amoako DG; Bhiman JN; Everatt J; Ismail A; Mahlangu B; Mnguni A; Mohale T; Ntuli N; Scheepers C |
| EPI_ISL_2013048, EPI_ISL_2013049, EPI_ISL_2013050, EPI_ISL_2013051, EPI_ISL_2013052, EPI_ISL_2013053, EPI_ISL_6913991, EPI_ISL_6913992, EPI_ISL_6913993, EPI_ISL_6913994, EPI_ISL_6913995, EPI_ISL_6913996, EPI_ISL_6913997, EPI_ISL_6913998, EPI_ISL_6913999, EPI_ISL_6914000, EPI_ISL_6914001, EPI_ISL_6914002, EPI_ISL_6914003, EPI_ISL_6914004, EPI_ISL_6914005, EPI_ISL_6914006, EPI_ISL_6914007 | | | |
| see above | Lancet Laboratories | National Institute for Communicable Diseases of the National Health Laboratory Service | Amoako DG; Bhiman JN; Everatt J; Glass A; Ismail A; Mahlangu B; Mnguni A; Mohale T; Ntuli N; Scheepers C; Viana R; Wolter N |
| EPI_ISL_2984866, EPI_ISL_2984867, EPI_ISL_2984868, EPI_ISL_2984869, EPI_ISL_2984870, EPI_ISL_2984871, EPI_ISL_2984872, EPI_ISL_2984873, EPI_ISL_3073888, EPI_ISL_3073889, EPI_ISL_3073890, EPI_ISL_3073891, EPI_ISL_3073892, EPI_ISL_3073893, EPI_ISL_3073894, EPI_ISL_3073895, EPI_ISL_3073896, EPI_ISL_3073897, EPI_ISL_3073898, EPI_ISL_3073899, EPI_ISL_3073900, EPI_ISL_3073901, EPI_ISL_3073910, EPI_ISL_3073911, EPI_ISL_3074049, EPI_ISL_3074050, EPI_ISL_3074051, EPI_ISL_3074052, EPI_ISL_3074053, EPI_ISL_3074054, EPI_ISL_3074055, EPI_ISL_3131775, EPI_ISL_3149307 | | | |
| see above | Lancet Laboratory | National Institute for Communicable Diseases of the National Health Laboratory Service | Amoako DG; Bhiman JN; Everatt J; Ismail A; Mahlangu B; Mnguni A; Mohale T; Ntuli N; Scheepers C |
| EPI_ISL_3024571, EPI_ISL_3024572, EPI_ISL_3237114, EPI_ISL_3237115 | MANKWENG LABORATORY | National Institute for Communicable Diseases of the National Health Laboratory Service | Amoako DG; Bhiman JN; Everatt J; Ismail A; Mahlangu B; Mnguni A; Mohale T; Ntuli N; Scheepers C |
| EPI_ISL_3024682 | MILPARK HOSPITAL | National Institute for Communicable Diseases of the National Health Laboratory Service | Amoako DG; Bhiman JN; Everatt J; Ismail A; Mahlangu B; Mnguni A; Mohale T; Ntuli N; Scheepers C; Sisonke Team |
| EPI_ISL_2271968 | Matikwana Hospital | National Institute for Communicable Diseases of the National Health Laboratory Service | Amoako DG; Bhiman JN; Ismail A; Mahlangu B; Mohale T; Ntuli N; Scheepers C |
| EPI_ISL_5918058, EPI_ISL_5918154, EPI_ISL_6913986, EPI_ISL_7456404 | Mobile Labs Gauteng | National Institute for Communicable Diseases of the National Health Laboratory Service | Amoako DG; Bhiman JN; Everatt J; Ismail A; Mahlangu B; Mnguni A; Mohale T; Ntuli N; Scheepers C; Wolter N |
| EPI_ISL_2710249, EPI_ISL_2710250, EPI_ISL_2710251, EPI_ISL_2710252, EPI_ISL_2710253, EPI_ISL_2710254, EPI_ISL_2710255, EPI_ISL_2710256, EPI_ISL_2710257, EPI_ISL_2710258, EPI_ISL_2710259, EPI_ISL_2710260, EPI_ISL_2710261, EPI_ISL_2710262, EPI_ISL_2710263, EPI_ISL_2710264, EPI_ISL_2710265, EPI_ISL_2710266, EPI_ISL_2710267, EPI_ISL_2710268, EPI_ISL_2710269, EPI_ISL_2710270, EPI_ISL_2710271, EPI_ISL_2710272, EPI_ISL_2710273, EPI_ISL_2710274, EPI_ISL_2710275, EPI_ISL_2710276, EPI_ISL_2710277, EPI_ISL_2710278, EPI_ISL_2710279, EPI_ISL_2710280, EPI_ISL_2710281, EPI_ISL_2710282, EPI_ISL_2710283, EPI_ISL_2710284, EPI_ISL_2710285, EPI_ISL_2710286, EPI_ISL_2710287, EPI_ISL_2710288, EPI_ISL_2710289, EPI_ISL_2710290, EPI_ISL_2710291, EPI_ISL_2710292, EPI_ISL_2710293, EPI_ISL_2710294, EPI_ISL_2710295, EPI_ISL_2710296, EPI_ISL_2710297, EPI_ISL_2710298, EPI_ISL_2710299, EPI_ISL_2710300, EPI_ISL_2710301, EPI_ISL_2710302, EPI_ISL_2710303, EPI_ISL_2710304, EPI_ISL_2710305, EPI_ISL_2726854, EPI_ISL_2726855, EPI_ISL_2726856, EPI_ISL_2770439, EPI_ISL_2770440, EPI_ISL_2770441, EPI_ISL_2770442, EPI_ISL_2770443, EPI_ISL_2770444, EPI_ISL_2770445, EPI_ISL_2770446, EPI_ISL_2770447, EPI_ISL_2770448, EPI_ISL_2770449, EPI_ISL_2770450, EPI_ISL_2770451, EPI_ISL_2770452, EPI_ISL_2770453, EPI_ISL_2770454, EPI_ISL_2770455, EPI_ISL_2770456, EPI_ISL_2770457, EPI_ISL_2770458, EPI_ISL_2770459, EPI_ISL_2770460, EPI_ISL_2770461, EPI_ISL_2770462, EPI_ISL_2770463, EPI_ISL_2770464, EPI_ISL_2770465, EPI_ISL_2770466, EPI_ISL_2770467, EPI_ISL_2770468, EPI_ISL_2770469, EPI_ISL_2770470, EPI_ISL_2770471, EPI_ISL_2770472, EPI_ISL_2770473, EPI_ISL_2770474, EPI_ISL_2770475, EPI_ISL_2770476, EPI_ISL_2770477, EPI_ISL_2770478, EPI_ISL_2770479, EPI_ISL_2770480, EPI_ISL_2770481, EPI_ISL_2770482, EPI_ISL_2770483, EPI_ISL_2770484, EPI_ISL_2770485, EPI_ISL_2770486, EPI_ISL_2770487, EPI_ISL_2770488, EPI_ISL_4572817, EPI_ISL_4572818, EPI_ISL_4572819, EPI_ISL_4572820, EPI_ISL_4572821, EPI_ISL_4572822, EPI_ISL_4572823, EPI_ISL_4572824, EPI_ISL_4572825, EPI_ISL_4572826, EPI_ISL_4572827, EPI_ISL_4572828, EPI_ISL_4572831, EPI_ISL_4572832, EPI_ISL_4572833, EPI_ISL_4572834, EPI_ISL_4572835, EPI_ISL_4572836, EPI_ISL_4572840, EPI_ISL_4572842, EPI_ISL_4572843, EPI_ISL_4572844, EPI_ISL_4572845, EPI_ISL_4572846, EPI_ISL_4572847, EPI_ISL_4572848, EPI_ISL_4572849, EPI_ISL_4572850, EPI_ISL_4572851, EPI_ISL_4572852, EPI_ISL_4572853, EPI_ISL_4572854, EPI_ISL_4572855, EPI_ISL_4572856, EPI_ISL_4572857, EPI_ISL_4572860, EPI_ISL_4572861, EPI_ISL_4572862, EPI_ISL_4572864, EPI_ISL_4572865, EPI_ISL_4572866, EPI_ISL_4572867 | | | |
| see above | NHLS Charlotte Maxeke Johannesburg Academic Hospital and the University of the Witwatersrand | KRISP, KZN Research Innovation and Sequencing Platform | Bulelani Manene; Florette Treurnicht; Giandhari Jennifer; Kathleen Subramoney; Naidoo Yeshnee; Pillay Sureshnee; San James; Sisonke; Tegally Houriiyah; Tshabuila Derek; Wilkinson Eduan; Yajna Ramphal; de Oliveira Tulio |
| EPI_ISL_2841643, EPI_ISL_2841644, EPI_ISL_2841645, EPI_ISL_2841646, EPI_ISL_2841647, EPI_ISL_2841648, EPI_ISL_2841649, EPI_ISL_3132494, EPI_ISL_3132495, EPI_ISL_3132496, EPI_ISL_3132497, EPI_ISL_3132498, EPI_ISL_3132499, EPI_ISL_3132500, EPI_ISL_3132501, EPI_ISL_3132502, EPI_ISL_3132503, EPI_ISL_3132504, EPI_ISL_3132505, EPI_ISL_3132506, EPI_ISL_3132507, EPI_ISL_3132508, EPI_ISL_3132509, EPI_ISL_3132510, EPI_ISL_3132511, EPI_ISL_3132512, EPI_ISL_3132513, EPI_ISL_3132514, EPI_ISL_3132515, EPI_ISL_3132516, EPI_ISL_3132517, EPI_ISL_3132518, EPI_ISL_3132519, EPI_ISL_3132520, EPI_ISL_3132521, EPI_ISL_3132522, EPI_ISL_3132523, EPI_ISL_3132524, EPI_ISL_3132525, EPI_ISL_3132526, EPI_ISL_3132527, EPI_ISL_3132528, EPI_ISL_3132529, EPI_ISL_3132530, EPI_ISL_3132531, EPI_ISL_3132532, EPI_ISL_3132533, EPI_ISL_3132534, EPI_ISL_3132535, EPI_ISL_3132536, EPI_ISL_3132537, EPI_ISL_3132538, EPI_ISL_3132539, EPI_ISL_3132540, EPI_ISL_3132541, EPI_ISL_3132542, EPI_ISL_3132543, EPI_ISL_3132544, EPI_ISL_3132545, EPI_ISL_3132546, EPI_ISL_3132547, EPI_ISL_3132548, EPI_ISL_3132549, EPI_ISL_3132550, EPI_ISL_3132551, EPI_ISL_3132552, EPI_ISL_3132553, EPI_ISL_3132554, EPI_ISL_3132555, EPI_ISL_3132556, EPI_ISL_3132557, EPI_ISL_3132558, EPI_ISL_3132559, EPI_ISL_3132560, EPI_ISL_3132561, EPI_ISL_3132562, EPI_ISL_3132563, EPI_ISL_3132564, EPI_ISL_3132565, EPI_ISL_3132566, EPI_ISL_3132567, EPI_ISL_3132568, EPI_ISL_3132569, EPI_ISL_3132570, EPI_ISL_3132571, EPI_ISL_3132572, EPI_ISL_3132573, EPI_ISL_3132623, EPI_ISL_3132624, EPI_ISL_3132625, EPI_ISL_3132626, EPI_ISL_3132627 | | | |
| see above | NHLS Charlotte Maxeke Johannesburg Academic Hospital and the University of the Witwatersrand | KRISP, KZn Research Innovation and Sequencing Platform | Bulelani Manene; Florette Treurnicht; Giandhari Jennifer; Kathleen Subramoney; Naidoo Yeshnee; Pillay Sureshnee; San James; Tegally Houriiyah; Tshabuila Derek; Wilkinson Eduan; Yajna Ramphal; de Oliveira Tulio |
| EPI_ISL_4473159, EPI_ISL_4473163, EPI_ISL_4473165, EPI_ISL_4473166, EPI_ISL_4473174, EPI_ISL_4473175, EPI_ISL_4473177, EPI_ISL_4473181, EPI_ISL_4473183, EPI_ISL_4473186, EPI_ISL_4473189, EPI_ISL_4473208, EPI_ISL_4473211, EPI_ISL_4473218, EPI_ISL_4473219, EPI_ISL_4473221, EPI_ISL_5098656, EPI_ISL_5098663, EPI_ISL_5098665, EPI_ISL_5098669, EPI_ISL_5098679, EPI_ISL_5098684, EPI_ISL_5098692, EPI_ISL_5098700, EPI_ISL_5098706, EPI_ISL_5098710, EPI_ISL_5098711, EPI_ISL_5098717, EPI_ISL_5098720, EPI_ISL_5098726, EPI_ISL_5098736, EPI_ISL_5098743, EPI_ISL_5098751, EPI_ISL_5098759, EPI_ISL_5098807, EPI_ISL_5098812, EPI_ISL_5098820, EPI_ISL_5098828, EPI_ISL_5098833, EPI_ISL_5098838, EPI_ISL_5098847, EPI_ISL_5098853, EPI_ISL_5098857, EPI_ISL_5098864, EPI_ISL_5098890, EPI_ISL_5098899, EPI_ISL_5098906, EPI_ISL_5098927, EPI_ISL_5098931, EPI_ISL_5098934, EPI_ISL_5098964, EPI_ISL_5098971, EPI_ISL_5098977, EPI_ISL_5098981, EPI_ISL_5098993, EPI_ISL_5099000, EPI_ISL_5099020, EPI_ISL_5099027, EPI_ISL_5099032, EPI_ISL_5099041, EPI_ISL_5099047, EPI_ISL_5099057, EPI_ISL_5099077, EPI_ISL_5099085, EPI_ISL_5099154, EPI_ISL_5099155, EPI_ISL_5099171, EPI_ISL_5099172, EPI_ISL_5099173, EPI_ISL_5099204, EPI_ISL_5099216, EPI_ISL_5429023, EPI_ISL_5429024, EPI_ISL_5429025, EPI_ISL_5429026, EPI_ISL_5429027, EPI_ISL_5429034, EPI_ISL_5429035, EPI_ISL_5429036, EPI_ISL_5429038, EPI_ISL_5429039, EPI_ISL_5429040, EPI_ISL_5429041, EPI_ISL_5429042, EPI_ISL_5429043, EPI_ISL_5429044, EPI_ISL_5429045, EPI_ISL_5429049, EPI_ISL_6422264, EPI_ISL_6422265, EPI_ISL_6422266, EPI_ISL_6422267, EPI_ISL_6422268, EPI_ISL_6422269, EPI_ISL_6422305, EPI_ISL_6422306, EPI_ISL_6422307, EPI_ISL_6422308, EPI_ISL_6422309, EPI_ISL_6422310, EPI_ISL_6422311, EPI_ISL_6422312, EPI_ISL_6422313, EPI_ISL_6422314, EPI_ISL_6422315, EPI_ISL_6422316, EPI_ISL_6422317, EPI_ISL_6422318 | | | |
| see above | NHLS Charlotte Maxeke Johannesburg Hospital | CERI, Centre for Epidemic Response and Innvoation, Stellenbosch University and KRISP, KZN Research Innovation and Sequencing Platform, UKZN. | Emmanuel SJ; Florette Treurnicht; Giandhari J; Giandhari Jennifer; Kathleen Subramoney; Naidoo Y; Naidoo Yeshnee; Pillay S; Pillay Sureshnee; San James; Tegally H; Tegally Houriiyah; Tshiabuila Derek; Upasana Ramphal; Wilkinson E; Wilkinson Eduan; Yajna Ramphal; de Oliveira T; de Oliveira Tulio |
| EPI_ISL_3728977, EPI_ISL_3728981, EPI_ISL_3729063, EPI_ISL_3729085, EPI_ISL_3729089, EPI_ISL_3729093, EPI_ISL_3729097, EPI_ISL_3729110, EPI_ISL_3729113, EPI_ISL_3729114, EPI_ISL_3729116, EPI_ISL_3729117, EPI_ISL_3729119, EPI_ISL_3729120, EPI_ISL_3729121, EPI_ISL_3729122, EPI_ISL_3729123, EPI_ISL_3729127, EPI_ISL_3729128, EPI_ISL_3729129, EPI_ISL_3729130, EPI_ISL_3729133, EPI_ISL_3729134, EPI_ISL_3729135, EPI_ISL_3729136, EPI_ISL_3729137, EPI_ISL_3729139, EPI_ISL_3729140, EPI_ISL_3729141, EPI_ISL_3729143, EPI_ISL_3729144, EPI_ISL_3729148, EPI_ISL_3729149, EPI_ISL_3729151, EPI_ISL_3729152, EPI_ISL_3729154, EPI_ISL_3729157, EPI_ISL_3729161, EPI_ISL_3729168, EPI_ISL_3729172, EPI_ISL_3729176, EPI_ISL_3729180, EPI_ISL_3729188, EPI_ISL_3729199, EPI_ISL_3729223, EPI_ISL_3729230, EPI_ISL_3729239, EPI_ISL_3729243, EPI_ISL_3729247, EPI_ISL_3729250, EPI_ISL_3859855, EPI_ISL_3859857, EPI_ISL_3859866, EPI_ISL_3859868, EPI_ISL_3859869, EPI_ISL_3859873, EPI_ISL_3859880, EPI_ISL_3859881, EPI_ISL_3859884, EPI_ISL_3859885, EPI_ISL_3859887, EPI_ISL_3859888, EPI_ISL_3859890, EPI_ISL_3859894, EPI_ISL_3859926, EPI_ISL_3859930, EPI_ISL_3859933, EPI_ISL_3859940, EPI_ISL_3859942, EPI_ISL_3859944, EPI_ISL_3859945, EPI_ISL_3859950, EPI_ISL_3859954, EPI_ISL_3859958, EPI_ISL_3859962, EPI_ISL_3859966, EPI_ISL_3859969, EPI_ISL_3859971, EPI_ISL_3859972, EPI_ISL_3859974, EPI_ISL_3859978, EPI_ISL_3859982, EPI_ISL_3859986, EPI_ISL_3859996, EPI_ISL_3859998, EPI_ISL_3859999, EPI_ISL_3860010, EPI_ISL_3860012, EPI_ISL_3860014, EPI_ISL_3860015, EPI_ISL_3860037, EPI_ISL_3860040, EPI_ISL_3860047, EPI_ISL_3860057, EPI_ISL_3860059, EPI_ISL_3860060, EPI_ISL_3860064, EPI_ISL_3860068, EPI_ISL_3860070, EPI_ISL_3860076, EPI_ISL_3860078, EPI_ISL_3860090, EPI_ISL_3860098, EPI_ISL_3860100, EPI_ISL_3860110, EPI_ISL_3860114, EPI_ISL_3860124, EPI_ISL_3860128, EPI_ISL_3860130, EPI_ISL_3860132, EPI_ISL_3860133, EPI_ISL_3860135, EPI_ISL_3860137, EPI_ISL_3860141, EPI_ISL_3860143, EPI_ISL_3860145, EPI_ISL_3860147, EPI_ISL_3860148, EPI_ISL_3860150, EPI_ISL_3860152, EPI_ISL_3860154, EPI_ISL_3860155, EPI_ISL_3860157, EPI_ISL_3860159, EPI_ISL_3860161, EPI_ISL_3860163, EPI_ISL_3860165, EPI_ISL_3860167, EPI_ISL_3860168, EPI_ISL_4572296, EPI_ISL_4572297, EPI_ISL_4572298, EPI_ISL_4572299, EPI_ISL_4572300, EPI_ISL_4572301, EPI_ISL_4572302, EPI_ISL_4572303, EPI_ISL_4572304, EPI_ISL_4572305, EPI_ISL_4572306, EPI_ISL_4572307, EPI_ISL_4572308, EPI_ISL_4572320, EPI_ISL_4572321, EPI_ISL_4572325, EPI_ISL_4572327, EPI_ISL_4572328, EPI_ISL_4572337, EPI_ISL_4572340, EPI_ISL_4572341, EPI_ISL_4572344, EPI_ISL_4572345, EPI_ISL_4572346, EPI_ISL_4572347, EPI_ISL_4572349, EPI_ISL_4572350, EPI_ISL_4572352, EPI_ISL_4572355, EPI_ISL_4572363, EPI_ISL_4572371, EPI_ISL_4572372, EPI_ISL_4572375, EPI_ISL_4572376, EPI_ISL_4572381, EPI_ISL_4572382, EPI_ISL_4572383, EPI_ISL_4572384, EPI_ISL_4572386, EPI_ISL_4572389 | | | |
| see above | NHLS Charlotte Maxeke Johannesburg Hospital | KRISP, KZN Research Innovation and Sequencing Platform | Emmanuel S; Florette Treurnicht; Giandhari J; Giandhari Jennifer; Kathleen Subramoney; Naidoo Yeshnee; Pillay S; Pillay Sureshnee; San James; Tegally H; Tegally Houriiyah; Tshabuila Derek; Wilkinson E; Wilkinson Eduan; Yajna Ramphal; de Oliveira T; de Oliveira Tulio |
| EPI_ISL_3722215, EPI_ISL_3722216, EPI_ISL_3722217, EPI_ISL_3722218, EPI_ISL_3722219, EPI_ISL_3722220, EPI_ISL_3722221, EPI_ISL_3722222, EPI_ISL_3722223, EPI_ISL_3722224, EPI_ISL_3722225, EPI_ISL_3722226, EPI_ISL_3722227, EPI_ISL_3722228, EPI_ISL_3722229, EPI_ISL_3722230, EPI_ISL_3722231, EPI_ISL_3722232, EPI_ISL_3722233, EPI_ISL_3722234, EPI_ISL_3722235, EPI_ISL_3722236, | | | |

| | | | |
|---|---|---|---|
| EPI_ISL_3722237, EPI_ISL_3722238, EPI_ISL_3722239, EPI_ISL_3722240, EPI_ISL_3722241, EPI_ISL_3722242, EPI_ISL_3722243, EPI_ISL_3722244, EPI_ISL_3722245, EPI_ISL_3722246, EPI_ISL_3722247, EPI_ISL_3722248, EPI_ISL_3722249, EPI_ISL_3722250, EPI_ISL_3722251, EPI_ISL_3722252, EPI_ISL_3722253, EPI_ISL_3722254, EPI_ISL_3722255, EPI_ISL_3722256, EPI_ISL_3722257, EPI_ISL_3722258, EPI_ISL_3722259, EPI_ISL_3722260, EPI_ISL_3722261, EPI_ISL_3722262, EPI_ISL_3722263, EPI_ISL_3722264, EPI_ISL_3722265, EPI_ISL_3722266, EPI_ISL_3722267, EPI_ISL_3722268, EPI_ISL_3722269, EPI_ISL_3722270, EPI_ISL_3722271, EPI_ISL_3722272, EPI_ISL_3722273, EPI_ISL_3722274, EPI_ISL_3722275, EPI_ISL_3722276, EPI_ISL_3722277, EPI_ISL_3722278, EPI_ISL_3799009, EPI_ISL_3799010, EPI_ISL_3799013, EPI_ISL_3799014, EPI_ISL_3799016, EPI_ISL_3799017, EPI_ISL_3799018, EPI_ISL_3799019, EPI_ISL_3799022, EPI_ISL_3799027, EPI_ISL_3799028, EPI_ISL_3799035, EPI_ISL_3799036, EPI_ISL_3799056, EPI_ISL_3799062, EPI_ISL_3799063, EPI_ISL_3799064, EPI_ISL_3799066, EPI_ISL_3799071, EPI_ISL_3799083, EPI_ISL_3799085, EPI_ISL_3799089, EPI_ISL_3799095, EPI_ISL_3799104, EPI_ISL_3799116, EPI_ISL_3799122, EPI_ISL_3799124, EPI_ISL_3799128, EPI_ISL_3799137, EPI_ISL_3799139, EPI_ISL_3799143, EPI_ISL_3799145, EPI_ISL_3799151, EPI_ISL_3799153, EPI_ISL_3799155, EPI_ISL_3799156, EPI_ISL_3799158, EPI_ISL_3799159, EPI_ISL_3799160, EPI_ISL_3799161, EPI_ISL_3799162, EPI_ISL_3799165, EPI_ISL_3799166, EPI_ISL_3799168, EPI_ISL_3799169, EPI_ISL_3799170, EPI_ISL_3799171, EPI_ISL_4121605, EPI_ISL_4121607, EPI_ISL_4121610, EPI_ISL_4121621, EPI_ISL_4121634, EPI_ISL_4121656, EPI_ISL_4121659, EPI_ISL_4121668, EPI_ISL_4121671, EPI_ISL_4121675, EPI_ISL_4121681, EPI_ISL_4121687, EPI_ISL_4121710, EPI_ISL_4121713, EPI_ISL_4121728, EPI_ISL_4121731, EPI_ISL_4121733, EPI_ISL_4121740, EPI_ISL_4121764, EPI_ISL_4121770, EPI_ISL_4121779, EPI_ISL_4121785, EPI_ISL_4336962, EPI_ISL_4336965, EPI_ISL_4336968, EPI_ISL_4336970 | | | |
| see above | NHLS Charlotte Maxeke Johannesburg Hospital | KRISP, KZn Research Innovation and Sequencing Platform | Emmanuel S; Florette Treurnicht; Giandhari J; Giandhari Jennifer; Kathleen Subramoney; Naidoo Yeshnee; Pillay S; Pillay Sureshnee; San James; Tegally H; Tegally Houriiyah; Tshabuila Derek; Tshiabuila Derek; Wilkinson E; Wilkinson Eduan; Yajna Ramphal; de Oliveira T; de Oliveira Tulio |
| EPI_ISL_3447706, EPI_ISL_3447707, EPI_ISL_3447708, EPI_ISL_3447713, EPI_ISL_3447714, EPI_ISL_3447715, EPI_ISL_3447718, EPI_ISL_3447728, EPI_ISL_3447729, EPI_ISL_3447734, EPI_ISL_3447737, EPI_ISL_3447738, EPI_ISL_3447739, EPI_ISL_3447743, EPI_ISL_3447744, EPI_ISL_3447746, EPI_ISL_3447748, EPI_ISL_3447754, EPI_ISL_3447760, EPI_ISL_3447761, EPI_ISL_3447762, EPI_ISL_3447763, EPI_ISL_3447764, EPI_ISL_3447765, EPI_ISL_3447766, EPI_ISL_3447767, EPI_ISL_3447768, EPI_ISL_3447769, EPI_ISL_3447770, EPI_ISL_3447771, EPI_ISL_3447772, EPI_ISL_3447773, EPI_ISL_3447774, EPI_ISL_3447775, EPI_ISL_3447776, EPI_ISL_3447777, EPI_ISL_3447778, EPI_ISL_3447780, EPI_ISL_3447781, EPI_ISL_3447783, EPI_ISL_3447784, EPI_ISL_3447785, EPI_ISL_3447786, EPI_ISL_3447787, EPI_ISL_3447788, EPI_ISL_3447789, EPI_ISL_3447790, EPI_ISL_3447791, EPI_ISL_3447792, EPI_ISL_3447793, EPI_ISL_3447794, EPI_ISL_3447795, EPI_ISL_3447796, EPI_ISL_3447798, EPI_ISL_3447799, EPI_ISL_3447800, EPI_ISL_3447803, EPI_ISL_3447804, EPI_ISL_3447806, EPI_ISL_3447807, EPI_ISL_3447808, EPI_ISL_3447809 | | | |
| see above | NHLS_JHB | KRISP, KZN Research Innovation and Sequencing Platform | Bulelani Manene; Florette Treurnicht; Giandhari Jennifer; Kathleen Subramoney; Naidoo Yeshnee; Pillay Sureshnee; San James; Tegally Houriiyah; Tshiabuila Derek; Wilkinson Eduan; Yajna Ramphal; de Oliveira Tulio |
| EPI_ISL_3261914, EPI_ISL_3261915, EPI_ISL_3261916, EPI_ISL_3261917, EPI_ISL_3261918, EPI_ISL_3261919, EPI_ISL_3261920, EPI_ISL_3261921, EPI_ISL_3261922, EPI_ISL_3261923 | | | |
| see above | NHLS_JHB (Ship 4) | KRISP, KZN Research Innovation and Sequencing Platform | Giandhari Jennifer; Naidoo Yeshnee; Pillay Sureshnee; San James; Tegally Houriiyah; Tshabuila Derek; Wilkinson Eduan; Yajna Ramphal; de Oliveira Tulio |
| EPI_ISL_4253647 | NICD staff | National Institute for Communicable Diseases of the National Health Laboratory Service | Amoako DG; Bhiman JN; Everatt J; Ismail A; Mahlangu B; Mnguni A; Mohale T; Ntuli N; Scheepers C; Sisonke Team |
| EPI_ISL_5510364 | Natalspruit | National Institute for Communicable Diseases of the National Health Laboratory Service | Amoako DG; Bhiman JN; Everatt J; Ismail A; Mahlangu B; Mnguni A; Mohale T; Ntuli N; Scheepers C |
| EPI_ISL_1363124, EPI_ISL_1363125, EPI_ISL_1363126, EPI_ISL_1363127, EPI_ISL_1363128, EPI_ISL_1363129, EPI_ISL_1363130, EPI_ISL_1363131, EPI_ISL_1363132, EPI_ISL_1363133, EPI_ISL_1363134, EPI_ISL_1363135, EPI_ISL_1363136, EPI_ISL_1363137, EPI_ISL_1363138, EPI_ISL_1363139, EPI_ISL_1363140, EPI_ISL_1363141, EPI_ISL_1363142, EPI_ISL_1363143, EPI_ISL_1363144, EPI_ISL_1363145, EPI_ISL_1372562 | | | |
| see above | National Health Laboratory Services | National Health Laboratory Services | Arshad Ismail; Florette Treurnicht; Kathleen Subramoney |
| EPI_ISL_6795188, EPI_ISL_6795189, EPI_ISL_6795190, EPI_ISL_6795191, EPI_ISL_6795192, EPI_ISL_6795193, EPI_ISL_6795194, EPI_ISL_7715346, EPI_ISL_7715347, EPI_ISL_7715348, EPI_ISL_7715349, EPI_ISL_7715350, EPI_ISL_7715351, EPI_ISL_7715352, EPI_ISL_7715353, EPI_ISL_7715354, EPI_ISL_7715355, EPI_ISL_7715356, EPI_ISL_7715357, EPI_ISL_7715358, EPI_ISL_7715359, EPI_ISL_7715360, EPI_ISL_7715361, EPI_ISL_7715362, EPI_ISL_7715363, EPI_ISL_7715364, EPI_ISL_7715365, EPI_ISL_7715366, EPI_ISL_7715367, EPI_ISL_7715368, EPI_ISL_7715369, EPI_ISL_7715370, EPI_ISL_7715371, EPI_ISL_7715372, EPI_ISL_7715373, EPI_ISL_7715374, EPI_ISL_7715375, EPI_ISL_7715376, EPI_ISL_7715377, EPI_ISL_7715378, EPI_ISL_7715379, EPI_ISL_7715380, EPI_ISL_7715381, EPI_ISL_7715382, EPI_ISL_7715383, EPI_ISL_7715384, EPI_ISL_7715385, EPI_ISL_7715386, EPI_ISL_7715387, EPI_ISL_7715388, EPI_ISL_7715389, EPI_ISL_7715390, EPI_ISL_7715391, EPI_ISL_7715392, EPI_ISL_7715393, EPI_ISL_7715394, EPI_ISL_7715395, EPI_ISL_7715396, EPI_ISL_7715397, EPI_ISL_7715398, EPI_ISL_7715399, EPI_ISL_7715400, EPI_ISL_7715401, EPI_ISL_7715402, EPI_ISL_7747503, EPI_ISL_7747504 | | | |
| see above | National Health Laboratory Services, Virology | CERI, Centre for Epidemic Response and Innvoation, Stellenbosch University and KRISP, KZN Research Innovation and Sequencing Platform, UKZN. | Arisha Maharaj; Avani Bharuthram; Florette Treurnicht; Giandhari J; Kathleen Subramoney; Naidoo Y; Nkhensani Mtileni; Pillay S; Ramphal U; Ramphal Y; San JE; Tegally H; Tshiabuila D; Wilkinson E; de Oliveira T |
| EPI_ISL_7852833, EPI_ISL_7852834, EPI_ISL_7852835, EPI_ISL_7852836, EPI_ISL_7852837, EPI_ISL_7852838, EPI_ISL_7852839, EPI_ISL_7852840, EPI_ISL_7852841, EPI_ISL_7852842, EPI_ISL_7852843, EPI_ISL_7852844, EPI_ISL_7852845, EPI_ISL_7852846, EPI_ISL_7852847, EPI_ISL_7852848, EPI_ISL_7852849, EPI_ISL_7852850, EPI_ISL_7852851, EPI_ISL_7852852, EPI_ISL_7852853, EPI_ISL_7852854, EPI_ISL_7852855, EPI_ISL_7852856, EPI_ISL_7852857, EPI_ISL_7852858, EPI_ISL_7852859, EPI_ISL_7852860, EPI_ISL_7852861, EPI_ISL_7852862, EPI_ISL_7852863, EPI_ISL_7852864, EPI_ISL_7852865, EPI_ISL_7852866, EPI_ISL_7852867, EPI_ISL_7852868, EPI_ISL_7852869, EPI_ISL_7852870, EPI_ISL_7852871, EPI_ISL_7852872, EPI_ISL_7852873, EPI_ISL_7852874, EPI_ISL_7852875, EPI_ISL_7852876, EPI_ISL_7852877, EPI_ISL_7852878, EPI_ISL_7852879, EPI_ISL_7852880, EPI_ISL_7852881, EPI_ISL_7852882, EPI_ISL_7852883, EPI_ISL_7852884, EPI_ISL_7852885, EPI_ISL_7852886, EPI_ISL_7852887, EPI_ISL_7852888, EPI_ISL_7852889, EPI_ISL_7852890, EPI_ISL_7852891 | | | |
| see above | National Health Laboratory Services, Virology | National Health Laboratory Services, Virology | Avani Bharuthram; Florette Treurnicht; Kathleen Subramoney; Nkhensani Mtileni |
| EPI_ISL_6699728, EPI_ISL_6699729, EPI_ISL_6699730, EPI_ISL_6699731, EPI_ISL_6699732, EPI_ISL_6699733, EPI_ISL_6699734, EPI_ISL_6699735, EPI_ISL_6699736, EPI_ISL_6699737, EPI_ISL_6699738, EPI_ISL_6699739, EPI_ISL_6699740, EPI_ISL_6699741, EPI_ISL_6699742, EPI_ISL_6699743, EPI_ISL_6699744, EPI_ISL_6699745, EPI_ISL_6699746, EPI_ISL_6699747, EPI_ISL_6699748, EPI_ISL_6699749, EPI_ISL_6699750, EPI_ISL_6699751, EPI_ISL_6699752, EPI_ISL_6699753, EPI_ISL_6699754, EPI_ISL_6699755, EPI_ISL_6699756, EPI_ISL_6699757, EPI_ISL_6699758, EPI_ISL_6699759, EPI_ISL_6699760, EPI_ISL_6699761, EPI_ISL_6699762, EPI_ISL_6699763, EPI_ISL_6699764, EPI_ISL_6699765, EPI_ISL_6699766, EPI_ISL_6699767, EPI_ISL_6699768, EPI_ISL_6699769, EPI_ISL_6699770, EPI_ISL_6699771, EPI_ISL_6782043, EPI_ISL_6782048, EPI_ISL_6782055, EPI_ISL_6782056, EPI_ISL_6782066, EPI_ISL_6782071, EPI_ISL_6782079, EPI_ISL_6782080, EPI_ISL_6782084, EPI_ISL_6782090, EPI_ISL_6782091, EPI_ISL_6782092, EPI_ISL_6810481, EPI_ISL_6810482, EPI_ISL_6810483, EPI_ISL_6810484, EPI_ISL_6810485, EPI_ISL_6810486, EPI_ISL_6810487, EPI_ISL_7015157, EPI_ISL_7015158, EPI_ISL_7015159, EPI_ISL_7015160, EPI_ISL_7015161, EPI_ISL_7015162, EPI_ISL_7015163, EPI_ISL_7015164, EPI_ISL_7015165, EPI_ISL_7015166, EPI_ISL_7015167, EPI_ISL_7015168, EPI_ISL_7015169, EPI_ISL_7015170, EPI_ISL_7417114, EPI_ISL_7417129, EPI_ISL_7417159, EPI_ISL_7417169, EPI_ISL_7417267, EPI_ISL_7417285, EPI_ISL_7417376, EPI_ISL_7417383, EPI_ISL_7417384 | | | |
| see above | National Health Laboratory Services, Virology, Charlotte Maxeke Johannesburg hospital, Parktown, Johannesburg, Gauteng | CERI, Centre for Epidemic Response and Innvoation, Stellenbosch University and KRISP, KZN Research Innovation and Sequencing Platform, UKZN. | Amoaka D; Arisha Maharaj; Avani Bharuthram; Bester P; Bhiman J; Engelbrecht S; Everatt J; Florette Treurnicht; Giandhari J; Goedhals D; Hardie D; Hsiao M; Iranzadeh A; Kathleen Subramoney; Lessells R; Makatini Z; Maponga T; Mdlalose N; Mlisana K; Moir M; NGS-SA (Scheepers C; Naidoo Y; Nkhensani Mtileni; Nyaga M) Giandhari J; Oluwakemi M; Pillay S; Preiser W; Ramphal U; Ramphal Y; San JE; Tegally H; Tshiabuila D; Venter M; Wilkinson E; Williamson C; de Oliveira T; von Gottberg A |
| EPI_ISL_4891489, EPI_ISL_4891510, EPI_ISL_4891522, EPI_ISL_4891528, EPI_ISL_4891579, EPI_ISL_4891586, EPI_ISL_4891588, EPI_ISL_4891595, EPI_ISL_4891610, EPI_ISL_4891613, EPI_ISL_4891639, EPI_ISL_4891645, EPI_ISL_4891656, EPI_ISL_4891661, EPI_ISL_4891667, EPI_ISL_4891844, EPI_ISL_4891850, EPI_ISL_4891851, EPI_ISL_4891852, EPI_ISL_5917845, EPI_ISL_5917864, EPI_ISL_5917908, EPI_ISL_5918045, EPI_ISL_5918057, EPI_ISL_5918182, EPI_ISL_5918183, EPI_ISL_5918184, EPI_ISL_5919750, EPI_ISL_7456447, EPI_ISL_7456448, EPI_ISL_7456452, EPI_ISL_7456455, EPI_ISL_7456456, EPI_ISL_7456457 | | | |
| see above | National Institute For Communicable Diseases Of The National Health Laboratory Service | National Institute for Communicable Diseases of the National Health Laboratory Service | Amoako DG; Bhiman JN; Everatt J; Ismail A; Mahlangu B; Mnguni A; Mohale T; Ntuli N; Scheepers C; Wolter N |
| EPI_ISL_2444091, EPI_ISL_2662708, EPI_ISL_2662709, EPI_ISL_2662710, EPI_ISL_2662711, EPI_ISL_2662712, EPI_ISL_2662713, EPI_ISL_2662714, EPI_ISL_2662715, EPI_ISL_2662716, EPI_ISL_2662717, EPI_ISL_2662718, EPI_ISL_2662719, EPI_ISL_2662720, EPI_ISL_2662721, EPI_ISL_2662722, EPI_ISL_2662723, EPI_ISL_2662724, EPI_ISL_2662725, EPI_ISL_2984801, EPI_ISL_3101635, EPI_ISL_3101636, EPI_ISL_3101637, EPI_ISL_3101638, EPI_ISL_3101656, EPI_ISL_3101657, EPI_ISL_3101658, EPI_ISL_3101659, EPI_ISL_3101660, EPI_ISL_3101661, EPI_ISL_3101662, EPI_ISL_3101663, EPI_ISL_3101664, EPI_ISL_3149306, EPI_ISL_3219783, EPI_ISL_3219784, EPI_ISL_3219785, EPI_ISL_3219786, EPI_ISL_3219787, EPI_ISL_3219788, EPI_ISL_3219789, EPI_ISL_3219790, EPI_ISL_3219791, EPI_ISL_3219792, EPI_ISL_3219793, EPI_ISL_3219794, EPI_ISL_3219795, EPI_ISL_3219796, EPI_ISL_3219797, EPI_ISL_3219798, EPI_ISL_3219799, EPI_ISL_3219800, EPI_ISL_3219801, EPI_ISL_3219802, EPI_ISL_3219803, EPI_ISL_3219804, EPI_ISL_3219805, EPI_ISL_3219806, EPI_ISL_3219807, EPI_ISL_3219808, EPI_ISL_3219809, EPI_ISL_3219810, EPI_ISL_3219811, EPI_ISL_3219812, EPI_ISL_3219813, EPI_ISL_3219814, EPI_ISL_3219815, EPI_ISL_3219816, EPI_ISL_3219817, EPI_ISL_3219818, EPI_ISL_3219819, EPI_ISL_3219820, EPI_ISL_3219821, EPI_ISL_3219822, EPI_ISL_3219823, EPI_ISL_3219824, EPI_ISL_3219825, EPI_ISL_3219826, EPI_ISL_3219827, EPI_ISL_3219828, EPI_ISL_3219829, EPI_ISL_3219830, EPI_ISL_3219831, EPI_ISL_3219832, EPI_ISL_3219833, EPI_ISL_3219835, EPI_ISL_3219836, EPI_ISL_3219837, EPI_ISL_3219838, EPI_ISL_3219839, EPI_ISL_3219840, EPI_ISL_3219841, EPI_ISL_3219842, EPI_ISL_3219843, EPI_ISL_3219844, EPI_ISL_3219845, EPI_ISL_3219846, EPI_ISL_3219847, EPI_ISL_3219848, EPI_ISL_3219849, EPI_ISL_3219850, EPI_ISL_3219851, EPI_ISL_3219852, EPI_ISL_3219853, EPI_ISL_3219854, EPI_ISL_3219855, EPI_ISL_3219856, EPI_ISL_3219857, EPI_ISL_3219858, EPI_ISL_3219859, EPI_ISL_3219860, EPI_ISL_3219861, EPI_ISL_3219862, EPI_ISL_3219863, EPI_ISL_3219864, EPI_ISL_3219865, EPI_ISL_3219878, EPI_ISL_3219879, EPI_ISL_3219880, EPI_ISL_3219881, EPI_ISL_3219882, EPI_ISL_3219883, EPI_ISL_3219884, EPI_ISL_3219885, EPI_ISL_3219886, EPI_ISL_3281585, EPI_ISL_3281586, EPI_ISL_3281588, EPI_ISL_3281589, EPI_ISL_3281590, EPI_ISL_3281591, EPI_ISL_3281592, EPI_ISL_3281593, EPI_ISL_3281594, EPI_ISL_3281595, EPI_ISL_3281596, EPI_ISL_3281597, EPI_ISL_3281598, EPI_ISL_3281599, EPI_ISL_3281600, EPI_ISL_3281601, EPI_ISL_3281602, EPI_ISL_3342435, EPI_ISL_3342436, EPI_ISL_3342437, EPI_ISL_3342438, EPI_ISL_3342439, EPI_ISL_3342440, EPI_ISL_3342441, EPI_ISL_3411817, EPI_ISL_3451570, EPI_ISL_3451571, EPI_ISL_3451572, EPI_ISL_3451573, EPI_ISL_3451574, EPI_ISL_3451575, EPI_ISL_3451576, EPI_ISL_3451577, EPI_ISL_3451593, EPI_ISL_3451634, EPI_ISL_3451636, EPI_ISL_3451637, EPI_ISL_3643852, EPI_ISL_3643957, EPI_ISL_3644044, EPI_ISL_3644066, EPI_ISL_3717965, EPI_ISL_3717994, EPI_ISL_3718045, EPI_ISL_3718046, EPI_ISL_3718047, EPI_ISL_3718066, EPI_ISL_4029915, EPI_ISL_4029941, EPI_ISL_4029956, EPI_ISL_4029973, EPI_ISL_4030023, EPI_ISL_4030024, EPI_ISL_4030069, EPI_ISL_4030084, EPI_ISL_4030100, EPI_ISL_4253671, EPI_ISL_4651683, EPI_ISL_4651747, EPI_ISL_4651872, EPI_ISL_4652276, EPI_ISL_5477158, EPI_ISL_5477595, EPI_ISL_5477673, EPI_ISL_6204411, EPI_ISL_6886555, EPI_ISL_6886558, EPI_ISL_7605585, EPI_ISL_7605586, EPI_ISL_7605593, EPI_ISL_7605594, EPI_ISL_7605629, EPI_ISL_7605630, EPI_ISL_7605631, EPI_ISL_7605632, EPI_ISL_7605637, EPI_ISL_7605638, EPI_ISL_7605666, EPI_ISL_7605720, EPI_ISL_7605759, EPI_ISL_7605762, EPI_ISL_7605765, EPI_ISL_7605767, EPI_ISL_7605768, EPI_ISL_7605770, EPI_ISL_7605771, EPI_ISL_8002879, EPI_ISL_8002880, EPI_ISL_8002883, EPI_ISL_8002884, EPI_ISL_8002885, EPI_ISL_8002886, EPI_ISL_8002889, EPI_ISL_8002890, EPI_ISL_8002891, EPI_ISL_8002893, EPI_ISL_8002894, EPI_ISL_8002895, EPI_ISL_8002896, EPI_ISL_8002899, EPI_ISL_8002902, EPI_ISL_8002903, EPI_ISL_8002904, EPI_ISL_8002905, EPI_ISL_8002907, EPI_ISL_8002908, EPI_ISL_8002909, EPI_ISL_8002910, EPI_ISL_8002911, EPI_ISL_8002912, EPI_ISL_8051873, EPI_ISL_8051874, EPI_ISL_8051875, EPI_ISL_8051876, EPI_ISL_8051877, EPI_ISL_8051882, EPI_ISL_8051883, EPI_ISL_8051884, EPI_ISL_8051885 | | | |
| see above | National Institute for Communicable Diseases of the National Health Laboratory Service | National Institute for Communicable Diseases of the National Health Laboratory Service | Amoako DG; Bhiman JN; Everatt J; Ismail A; Kekana D; Mahlangu B; Mnguni A; Mohale T; Ntuli N; Scheepers C; Sisonke Team; Wolter N |
| EPI_ISL_939656, EPI_ISL_939657, EPI_ISL_939658, EPI_ISL_939659, EPI_ISL_940067, EPI_ISL_1239850, EPI_ISL_1239882, EPI_ISL_1827555, EPI_ISL_1827556, EPI_ISL_1827557, EPI_ISL_1827558, EPI_ISL_1827559, EPI_ISL_1827560, EPI_ISL_1827561, EPI_ISL_1827562, EPI_ISL_1827563, EPI_ISL_1827564, EPI_ISL_1827565, EPI_ISL_1827566, EPI_ISL_1827567, EPI_ISL_1827568, EPI_ISL_1827569, EPI_ISL_1827570, EPI_ISL_1827571, EPI_ISL_1827572, EPI_ISL_1827573, EPI_ISL_1827574, EPI_ISL_1827575, EPI_ISL_1827576, EPI_ISL_1827577, EPI_ISL_1827578, EPI_ISL_1827579, EPI_ISL_1827580, EPI_ISL_1827581, EPI_ISL_1827582, EPI_ISL_1827583, EPI_ISL_1827584, EPI_ISL_1827585, EPI_ISL_1827586, EPI_ISL_1827587, EPI_ISL_1827588, EPI_ISL_1827589, EPI_ISL_1827590, EPI_ISL_1827591, EPI_ISL_1827592, EPI_ISL_1827593, EPI_ISL_1827594, EPI_ISL_1827595, EPI_ISL_1827596, EPI_ISL_1827597, EPI_ISL_1827598, EPI_ISL_1827599, EPI_ISL_1827600, EPI_ISL_1827601, EPI_ISL_1827602, EPI_ISL_1827603, EPI_ISL_1827604, EPI_ISL_1827605, EPI_ISL_1827606, EPI_ISL_1827607, EPI_ISL_1827608, EPI_ISL_1827609, EPI_ISL_1827610, EPI_ISL_1827611, EPI_ISL_1827612, EPI_ISL_1827613, EPI_ISL_1827614, EPI_ISL_1827615, EPI_ISL_1827616, EPI_ISL_1827618, EPI_ISL_1827620, EPI_ISL_1827621, EPI_ISL_1827622, EPI_ISL_1827623, EPI_ISL_1827624, EPI_ISL_1827625, EPI_ISL_1827626, EPI_ISL_1827627, EPI_ISL_1827628, EPI_ISL_1827629, EPI_ISL_1827630, EPI_ISL_1827631, EPI_ISL_1827632, EPI_ISL_1827633, EPI_ISL_1914786, EPI_ISL_1914787, EPI_ISL_1914788, EPI_ISL_1914789, EPI_ISL_1914790, EPI_ISL_1914791, EPI_ISL_1914792, EPI_ISL_1914793, EPI_ISL_1914794, EPI_ISL_1914795, EPI_ISL_1914796, EPI_ISL_1914797, EPI_ISL_1914798, EPI_ISL_1914799, EPI_ISL_1914800, EPI_ISL_1914801, EPI_ISL_1914802, EPI_ISL_1914803, EPI_ISL_1914804, EPI_ISL_1914805, EPI_ISL_1914806, EPI_ISL_1914807, EPI_ISL_1914808, EPI_ISL_1914809, EPI_ISL_1914810, EPI_ISL_1914811, EPI_ISL_1914812, EPI_ISL_1914813, EPI_ISL_1914814, EPI_ISL_1914815, EPI_ISL_1914816, EPI_ISL_1914817, EPI_ISL_1914818, EPI_ISL_1914819, EPI_ISL_1914820, EPI_ISL_1914821, EPI_ISL_1914822, EPI_ISL_1914823, EPI_ISL_1914824, EPI_ISL_1914825, EPI_ISL_1914826, EPI_ISL_1914827, EPI_ISL_1914828, EPI_ISL_1914829, EPI_ISL_1914830, EPI_ISL_1914831, EPI_ISL_1914832, EPI_ISL_1914833, EPI_ISL_1914834, EPI_ISL_1914835, EPI_ISL_1914836, EPI_ISL_1914837, EPI_ISL_1914838, EPI_ISL_1914839, EPI_ISL_1914840, EPI_ISL_1914841, EPI_ISL_1914842, EPI_ISL_1914843, EPI_ISL_1914845, EPI_ISL_1914848, EPI_ISL_1914849, EPI_ISL_1914850, EPI_ISL_1914851, EPI_ISL_1914852, EPI_ISL_1914853, EPI_ISL_1914854, EPI_ISL_1914855, EPI_ISL_1914856, EPI_ISL_1914857, EPI_ISL_1914858, EPI_ISL_1914859, EPI_ISL_1914860, EPI_ISL_1914861, EPI_ISL_1914862, EPI_ISL_1914863, EPI_ISL_1914864, EPI_ISL_1914865, EPI_ISL_1914866, EPI_ISL_1914867, EPI_ISL_1914868, EPI_ISL_1914869, EPI_ISL_1914870, EPI_ISL_1914871, EPI_ISL_1914872, EPI_ISL_1914873, EPI_ISL_1914874, EPI_ISL_1914875, EPI_ISL_1914876, EPI_ISL_1914877, EPI_ISL_1914878, EPI_ISL_1914879, EPI_ISL_1914880, EPI_ISL_1914881, EPI_ISL_1914882, EPI_ISL_1914883, EPI_ISL_1914884, EPI_ISL_1914885, EPI_ISL_1914887, EPI_ISL_1914888, EPI_ISL_1914889, EPI_ISL_1914890, EPI_ISL_1914891, EPI_ISL_1914892, EPI_ISL_1914893, EPI_ISL_1914894, EPI_ISL_1914895, EPI_ISL_1914896, EPI_ISL_1914897, EPI_ISL_1914898, EPI_ISL_1914899, EPI_ISL_1914900, EPI_ISL_1914901, EPI_ISL_1914902, EPI_ISL_1914903, EPI_ISL_1914904, EPI_ISL_1914905, EPI_ISL_1914906, EPI_ISL_1914907, EPI_ISL_1914908, EPI_ISL_1914909, EPI_ISL_1914910, EPI_ISL_1914911, EPI_ISL_1914912, EPI_ISL_1914913, EPI_ISL_1914914, EPI_ISL_1914915, EPI_ISL_1914916, EPI_ISL_1914917, EPI_ISL_1914918, EPI_ISL_1914919, EPI_ISL_1914920, EPI_ISL_1914921, EPI_ISL_1914922, EPI_ISL_1914923, EPI_ISL_1914924, EPI_ISL_1914925, EPI_ISL_1914926, EPI_ISL_1914927, EPI_ISL_1914928, EPI_ISL_1914929, EPI_ISL_1914930, EPI_ISL_1914932, EPI_ISL_1914933, EPI_ISL_1914934, EPI_ISL_1914935 | | | |
| see above | National Institute for Communicable Diseases,National Health Laboratory Services, Gauteng, South Africa | National Institute for Communicable Diseases of the National Health Laboratory Service | Allam M; Amoako DG; Bhiman JN; Ismail A; Mahlangu B; Mohale T; Ntuli N; Scheepers C |
| EPI_ISL_4575097, EPI_ISL_4575118, EPI_ISL_4575178, EPI_ISL_4575179 | Netcare Linksfield | KRISP, KZN Research Innovation and Sequencing Platform | Emmanuel SJ; Giandhari J; Lessells R; Naidoo Y; Pillay S; Ramphal U; Tegally H; Wilkinson E; de Oliveira T |
| EPI_ISL_2086211, EPI_ISL_2086212, EPI_ISL_2086213, EPI_ISL_2086214, EPI_ISL_2086215, EPI_ISL_2086216, EPI_ISL_2086217, EPI_ISL_2086218, EPI_ISL_2086219 | | | |
| see above | Netcare Linksfield Hospital | KRISP, KZN Research Innovation and Sequencing Platform | Emmanuel S; Emmanuel SJ; Giandhari J; Lessells R; Naidoo Y; Pillay S; Ramphal U; Tegally H; Wilkinson E; de Oliveira T |
| EPI_ISL_4575055, EPI_ISL_4575199 | Netcare Milpark | KRISP, KZN Research Innovation and Sequencing Platform | Emmanuel SJ; Giandhari J; Lessells R; Naidoo Y; Pillay S; Ramphal U; Tegally H; Wilkinson E; de Oliveira T |
| EPI_ISL_4575056 | Netcare Olivedale Hospital | KRISP, KZN Research Innovation and Sequencing Platform | Emmanuel SJ; Giandhari J; Lessells R; Naidoo Y; Pillay S; Ramphal U; Tegally H; Wilkinson E; de Oliveira T |
| EPI_ISL_5510409 | ORT Mobile lab | National Institute for Communicable Diseases of the National Health Laboratory Service | Amoako DG; Bhiman JN; Everatt J; Ismail A; Mahlangu B; Mnguni A; Mohale T; Ntuli N; Scheepers C |
| EPI_ISL_2086977, EPI_ISL_2086978 | PATH CARE | National Institute for Communicable Diseases of the National Health Laboratory Service | Amoako DG; Bhiman JN; Ismail A; Mahlangu B; Mohale T; Ntuli N; Scheepers C |
| EPI_ISL_3838570, EPI_ISL_3838571, EPI_ISL_4651723, EPI_ISL_4651764, EPI_ISL_4651842, EPI_ISL_4652073, EPI_ISL_4652083, EPI_ISL_6327912 | | | |
| see above | PATHCARE | National Institute for Communicable Diseases of the National Health Laboratory Service | Amoako DG; Bhiman JN; Everatt J; Ismail A; Mahlangu B; Mnguni A; Mohale T; Ntuli N; Scheepers C |
| EPI_ISL_2695583, EPI_ISL_2695584, EPI_ISL_2695585, EPI_ISL_2695586, EPI_ISL_2695587 | PATHCARE LABORATORY | National Institute for Communicable Diseases of the National Health Laboratory Service | Amoako DG; Bhiman JN; Everatt J; Ismail A; Mahlangu B; Mnguni A; Mohale T; Ntuli N; Scheepers C |

| | | | |
|---|---|---|---|
| EPI_ISL_8051886, EPI_ISL_8051887, EPI_ISL_8051888 | PATHCARe LABORATORY | National Institute for Communicable Diseases of the National Health Laboratory Service | Amoako DG; Bhiman JN; Everatt J; Ismail A; Kekana D; Mahlangu B; Mnguni A; Mohale T; Ntuli N; Scheepers C; Wolter N |
| EPI_ISL_2709978 | PHOLOSONG LABORATORY | National Institute for Communicable Diseases of the National Health Laboratory Service | Amoako DG; Bhiman JN; Everatt J; Ismail A; Mahlangu B; Mnguni A; Mohale T; Ntuli N; Scheepers C |
| EPI_ISL_3237004, EPI_ISL_3237005, EPI_ISL_3237006, EPI_ISL_3237007, EPI_ISL_3237008, EPI_ISL_3237009, EPI_ISL_3237010, EPI_ISL_3237011, EPI_ISL_3237012, EPI_ISL_3237013, EPI_ISL_3237016, EPI_ISL_3237017, EPI_ISL_3237018, EPI_ISL_3237019, EPI_ISL_3237020, EPI_ISL_3237197, EPI_ISL_3237198, EPI_ISL_3237199, EPI_ISL_3237200, EPI_ISL_3237201, EPI_ISL_3237202, EPI_ISL_3237203, EPI_ISL_3237204, EPI_ISL_3237205, EPI_ISL_3237206, EPI_ISL_3237207, EPI_ISL_3237208, EPI_ISL_3237218, EPI_ISL_3237219, EPI_ISL_3237220, EPI_ISL_3237221 | | | |
| see above | PORT ELIZABETH | National Institute for Communicable Diseases of the National Health Laboratory Service | Amoako DG; Bhiman JN; Everatt J; Ismail A; Mahlangu B; Mnguni A; Mohale T; Ntuli N; Scheepers C |
| EPI_ISL_3411695 | PORT ELIZABETH LABORATORY | National Institute for Communicable Diseases of the National Health Laboratory Service | Amoako DG; Bhiman JN; Everatt J; Ismail A; Mahlangu B; Mnguni A; Mohale T; Ntuli N; Scheepers C |
| EPI_ISL_3717896, EPI_ISL_3717898, EPI_ISL_3717903, EPI_ISL_3717907, EPI_ISL_3717909, EPI_ISL_3717911, EPI_ISL_3717920, EPI_ISL_3717932, EPI_ISL_3717935, EPI_ISL_3717942, EPI_ISL_3717946, EPI_ISL_3717948, EPI_ISL_3717972, EPI_ISL_3717975, EPI_ISL_3717979, EPI_ISL_3717980, EPI_ISL_3717982, EPI_ISL_3717993, EPI_ISL_3717999, EPI_ISL_3718000, EPI_ISL_3718038, EPI_ISL_3718039, EPI_ISL_3718040, EPI_ISL_3718041, EPI_ISL_3718042, EPI_ISL_3718043, EPI_ISL_3718044, EPI_ISL_5510391, EPI_ISL_5917989, EPI_ISL_5918035 | | | |
| see above | Pathcare | National Institute for Communicable Diseases of the National Health Laboratory Service | Amoako DG; Bhiman JN; Everatt J; Ismail A; Mahlangu B; Mnguni A; Mohale T; Ntuli N; Scheepers C; Sisonke Team |
| EPI_ISL_2828734, EPI_ISL_2828735, EPI_ISL_2828736, EPI_ISL_2828737, EPI_ISL_2828738, EPI_ISL_2828739, EPI_ISL_2828740, EPI_ISL_2828741, EPI_ISL_2828742, EPI_ISL_2828743, EPI_ISL_2828744, EPI_ISL_2828745, EPI_ISL_2828746, EPI_ISL_2828747, EPI_ISL_2828748, EPI_ISL_2828749, EPI_ISL_2828750, EPI_ISL_2828751, EPI_ISL_2828752, EPI_ISL_2828753, EPI_ISL_2828754, EPI_ISL_2828755, EPI_ISL_2828756, EPI_ISL_2828757, EPI_ISL_2828758, EPI_ISL_3101583, EPI_ISL_3101584, EPI_ISL_3101585, EPI_ISL_3101586, EPI_ISL_3101587, EPI_ISL_3101588, EPI_ISL_3101589, EPI_ISL_3101590, EPI_ISL_3101591, EPI_ISL_3411556, EPI_ISL_3411557, EPI_ISL_3411558, EPI_ISL_3411559, EPI_ISL_3411560, EPI_ISL_3411561, EPI_ISL_3411562, EPI_ISL_3411563, EPI_ISL_3411564, EPI_ISL_3411565, EPI_ISL_3411566, EPI_ISL_3411567, EPI_ISL_3411568, EPI_ISL_3411569, EPI_ISL_3411570, EPI_ISL_3411576, EPI_ISL_3411577, EPI_ISL_3411578, EPI_ISL_3411579, EPI_ISL_3411581, EPI_ISL_3411582, EPI_ISL_3411583, EPI_ISL_3411681, EPI_ISL_5510334, EPI_ISL_5510349, EPI_ISL_5510486 | | | |
| see above | Pathcare Vaal | National Institute for Communicable Diseases of the National Health Laboratory Service | Amoako DG; Bhiman JN; Everatt J; Ismail A; Mahlangu B; Mnguni A; Mohale T; Ntuli N; Scheepers C |
| EPI_ISL_2984940, EPI_ISL_2984941, EPI_ISL_2984942, EPI_ISL_2984943, EPI_ISL_2984944, EPI_ISL_2984945, EPI_ISL_2984946, EPI_ISL_2984947, EPI_ISL_2984948, EPI_ISL_2984949, EPI_ISL_2984950, EPI_ISL_2984951, EPI_ISL_2984952, EPI_ISL_2984953, EPI_ISL_2988410 | | | |
| see above | Pathcare Vaal Laboratory | National Institute for Communicable Diseases of the National Health Laboratory Service | Amoako DG; Bhiman JN; Everatt J; Ismail A; Mahlangu B; Mnguni A; Mohale T; Ntuli N; Scheepers C |
| EPI_ISL_3236988, EPI_ISL_3237163, EPI_ISL_3237164, EPI_ISL_3237165, EPI_ISL_3237166 | Pathcare Vaal laboratory | National Institute for Communicable Diseases of the National Health Laboratory Service | Amoako DG; Bhiman JN; Everatt J; Ismail A; Mahlangu B; Mnguni A; Mohale T; Ntuli N; Scheepers C |
| EPI_ISL_2013036, EPI_ISL_2013038, EPI_ISL_2013039, EPI_ISL_2013041, EPI_ISL_2013042 | Pathcare-Vermaak Centurion | National Institute for Communicable Diseases of the National Health Laboratory Service | Amoako DG; Bhiman JN; Glass A; Gottberg A; Mahlangu B; Mohale T; Ntuli N; Oliveira TD; Scheepers C; Tegally H; Viana R |
| EPI_ISL_5510288, EPI_ISL_5510290, EPI_ISL_5510371, EPI_ISL_5510418, EPI_ISL_5510464, EPI_ISL_5510466, EPI_ISL_5510475, EPI_ISL_5510485 | | | |
| see above | Pathcare/Vermaak Centurion | National Institute for Communicable Diseases of the National Health Laboratory Service | Amoako DG; Bhiman JN; Everatt J; Ismail A; Mahlangu B; Mnguni A; Mohale T; Ntuli N; Scheepers C |
| EPI_ISL_3838601, EPI_ISL_3838602, EPI_ISL_3838604, EPI_ISL_3838606, EPI_ISL_3838607, EPI_ISL_3838608, EPI_ISL_3838610, EPI_ISL_3838611, EPI_ISL_3838612, EPI_ISL_3838613, EPI_ISL_3838614, EPI_ISL_3838615, EPI_ISL_3838616, EPI_ISL_3838617, EPI_ISL_3838618, EPI_ISL_3838619, EPI_ISL_3838620, EPI_ISL_3838621, EPI_ISL_3838622, EPI_ISL_3838623, EPI_ISL_3838624, EPI_ISL_3838625, EPI_ISL_3838626, EPI_ISL_3838627, EPI_ISL_3838628, EPI_ISL_3838629, EPI_ISL_3838630, EPI_ISL_3838631, EPI_ISL_3838632, EPI_ISL_3838633, EPI_ISL_3838634, EPI_ISL_3838635, EPI_ISL_3838636, EPI_ISL_3838637, EPI_ISL_3838639, EPI_ISL_3838640, EPI_ISL_3838641, EPI_ISL_3838642, EPI_ISL_3838643, EPI_ISL_3838644, EPI_ISL_3838645, EPI_ISL_3838646, EPI_ISL_3838647, EPI_ISL_3838648, EPI_ISL_3838649, EPI_ISL_3838650, EPI_ISL_3838651, EPI_ISL_3838652, EPI_ISL_3838654, EPI_ISL_3838655, EPI_ISL_3838656, EPI_ISL_3838657, EPI_ISL_3838658, EPI_ISL_4253648, EPI_ISL_4253649, EPI_ISL_4253650, EPI_ISL_4253651, EPI_ISL_4253652, EPI_ISL_4253653, EPI_ISL_4253654, EPI_ISL_4253655, EPI_ISL_4253656, EPI_ISL_4253657, EPI_ISL_4253658, EPI_ISL_4253659, EPI_ISL_4253660, EPI_ISL_4253661, EPI_ISL_4253662, EPI_ISL_4253663, EPI_ISL_4253664, EPI_ISL_4253665, EPI_ISL_4253666, EPI_ISL_4253667, EPI_ISL_4253668, EPI_ISL_4253669, EPI_ISL_4253670, EPI_ISL_4651691, EPI_ISL_4651856, EPI_ISL_4651857, EPI_ISL_4651865, EPI_ISL_4652202, EPI_ISL_4652208, EPI_ISL_4652210, EPI_ISL_4652211, EPI_ISL_4652215, EPI_ISL_4652231, EPI_ISL_4652237, EPI_ISL_4652248, EPI_ISL_4652253, EPI_ISL_4652265, EPI_ISL_4652270 | | | |
| see above | Private/Pathcare | National Institute for Communicable Diseases of the National Health Laboratory Service | Amoako DG; Bhiman JN; Everatt J; Ismail A; Mahlangu B; Mnguni A; Mohale T; Ntuli N; Scheepers C; Sisonke Team |
| EPI_ISL_4651817, EPI_ISL_4651980 | ROB FERREIRA LABORATORY | National Institute for Communicable Diseases of the National Health Laboratory Service | Amoako DG; Bhiman JN; Everatt J; Ismail A; Mahlangu B; Mnguni A; Mohale T; Ntuli N; Scheepers C |
| EPI_ISL_3236982, EPI_ISL_3236983, EPI_ISL_3236984, EPI_ISL_3236985, EPI_ISL_3236993, EPI_ISL_3236994, EPI_ISL_3236995, EPI_ISL_3236996, EPI_ISL_3236997, EPI_ISL_3236998, EPI_ISL_3236999, EPI_ISL_3237001, EPI_ISL_3237002, EPI_ISL_3237003, EPI_ISL_3237150, EPI_ISL_3237151, EPI_ISL_3237152, EPI_ISL_3237153, EPI_ISL_3237173, EPI_ISL_3237174, EPI_ISL_3237175, EPI_ISL_3237176, EPI_ISL_3237177, EPI_ISL_3237178, EPI_ISL_3237179, EPI_ISL_3237180, EPI_ISL_3237181, EPI_ISL_3237182, EPI_ISL_3237183, EPI_ISL_3237184, EPI_ISL_3237185, EPI_ISL_3237186, EPI_ISL_3237187, EPI_ISL_3237188, EPI_ISL_3237189, EPI_ISL_3237190, EPI_ISL_3237191, EPI_ISL_3237192, EPI_ISL_3237193, EPI_ISL_3237194, EPI_ISL_3237195, EPI_ISL_3237196, EPI_ISL_5416476 | | | |
| see above | RUSTENBURG LABORATORY | National Institute for Communicable Diseases of the National Health Laboratory Service | Amoako DG; Bhiman JN; Everatt J; Ismail A; Mahlangu B; Mnguni A; Mohale T; Ntuli N; Scheepers C |
| EPI_ISL_4891541 | Rob Ferreira Laboratory | National Institute for Communicable Diseases of the National Health Laboratory Service | Amoako DG; Bhiman JN; Everatt J; Ismail A; Mahlangu B; Mnguni A; Mohale T; Ntuli N; Scheepers C |
| EPI_ISL_7545637, EPI_ISL_7545638, EPI_ISL_7545639, EPI_ISL_7545640, EPI_ISL_7545641, EPI_ISL_7545642, EPI_ISL_7545643, EPI_ISL_7545644, EPI_ISL_7545645, EPI_ISL_7545646, EPI_ISL_7545647, EPI_ISL_7545648, EPI_ISL_7545649, EPI_ISL_7545650, EPI_ISL_7545651 | | | |
| see above | SAMRC | CERI, Centre for Epidemic Response and Innvoation, Stellenbosch University and KRISP, KZN Research Innovation and Sequencing Platform, UKZN. | Arisha Maharaj; Giandhari J; MRC; Naidoo Y; Pillay S; Ramphal U; Ramphal Y; San JE; Tegally H; Tshiabuila D; Wilkinson E; de Oliveira T |
| EPI_ISL_5510477 | SOUTH RAND HOSPITAL | National Institute for Communicable Diseases of the National Health Laboratory Service | Amoako DG; Bhiman JN; Everatt J; Ismail A; Mahlangu B; Mnguni A; Mohale T; Ntuli N; Scheepers C |
| EPI_ISL_2086909, EPI_ISL_2086914, EPI_ISL_2086916, EPI_ISL_2086917, EPI_ISL_2086918, EPI_ISL_2086919, EPI_ISL_2086920, EPI_ISL_2086925, EPI_ISL_2086926, EPI_ISL_2086927, EPI_ISL_2086928, EPI_ISL_2086929, EPI_ISL_2086930 | | | |
| see above | Sebokeng Lab | National Institute for Communicable Diseases of the National Health Laboratory Service | Amoako DG; Bhiman JN; Ismail A; Mahlangu B; Mohale T; Ntuli N; Scheepers C |
| EPI_ISL_3267722, EPI_ISL_3267723, EPI_ISL_3267724, EPI_ISL_3267726, EPI_ISL_3267727, EPI_ISL_3267728, EPI_ISL_3267729, EPI_ISL_3267730, EPI_ISL_3267731, EPI_ISL_3267732, EPI_ISL_3267733, EPI_ISL_3267734, EPI_ISL_3267735, EPI_ISL_3267737, EPI_ISL_3267738, EPI_ISL_3267739, EPI_ISL_3267742, EPI_ISL_3267743, EPI_ISL_3267744, EPI_ISL_3267745, EPI_ISL_3267746, EPI_ISL_3267747 | | | |
| see above | Sisonke - BARC SA | CERI, Centre for Epidemic Response and Innovation, Stellenbosch Univeristy & KRISP, KZN Research Innovation and Sequencing Platform | Giandhari Jennifer; Naidoo Yeshnee; Pillay Sureshnee; San James; Tegally Houriiyah; Tshabuila Derek; Wilkinson Eduan; Yajna Ramphal; de Oliveira Tulio |
| EPI_ISL_2694611, EPI_ISL_2694612, EPI_ISL_2694613, EPI_ISL_2694616, EPI_ISL_2694617, EPI_ISL_2694618, EPI_ISL_2694619, EPI_ISL_2694620, EPI_ISL_2694621, EPI_ISL_2694622, EPI_ISL_2694623, EPI_ISL_2694624, EPI_ISL_2694625, EPI_ISL_2694626, EPI_ISL_2694632, EPI_ISL_2694678, EPI_ISL_2694679, EPI_ISL_2694680, EPI_ISL_2694681, EPI_ISL_2694682, EPI_ISL_2694683, EPI_ISL_2694684, EPI_ISL_2694685, EPI_ISL_2694686, EPI_ISL_2694687, EPI_ISL_2694688, EPI_ISL_2694689, EPI_ISL_2694690, EPI_ISL_2694691, EPI_ISL_2694692, EPI_ISL_2694693, EPI_ISL_2694694, EPI_ISL_2694695, EPI_ISL_2694696, EPI_ISL_2694697, EPI_ISL_2694698, EPI_ISL_2694699, EPI_ISL_2694700, EPI_ISL_2694701, EPI_ISL_2694702, EPI_ISL_2694703, EPI_ISL_2694704, EPI_ISL_2694705, EPI_ISL_2694706, EPI_ISL_2694707, EPI_ISL_2694708, EPI_ISL_2694709, EPI_ISL_2694710, EPI_ISL_2694711, EPI_ISL_2694712, EPI_ISL_2694713, EPI_ISL_2694714, EPI_ISL_2694715, EPI_ISL_2694716, EPI_ISL_2694717, EPI_ISL_2694718, EPI_ISL_2694719, EPI_ISL_2694720, EPI_ISL_2694721, EPI_ISL_2694722, EPI_ISL_2694723, EPI_ISL_2694724, EPI_ISL_2694725, EPI_ISL_2694726, EPI_ISL_2695504, EPI_ISL_2695505, EPI_ISL_2695507, EPI_ISL_2695508, EPI_ISL_2695509, EPI_ISL_2695510, EPI_ISL_2695511, EPI_ISL_2695514, EPI_ISL_2695515, EPI_ISL_2695516, EPI_ISL_2695517, EPI_ISL_2695518, EPI_ISL_2695519, EPI_ISL_2695520, EPI_ISL_2695521, EPI_ISL_2695522, EPI_ISL_2695525, EPI_ISL_2695526, EPI_ISL_2695527, EPI_ISL_2695528, EPI_ISL_2695529, EPI_ISL_3101535, EPI_ISL_3101537, EPI_ISL_3101538, EPI_ISL_3101539, EPI_ISL_3101540, EPI_ISL_3101541, EPI_ISL_3101542, EPI_ISL_3101543, EPI_ISL_3101544, EPI_ISL_3101545, EPI_ISL_3101546, EPI_ISL_3101547, EPI_ISL_3101548, EPI_ISL_3101549, EPI_ISL_3101550, EPI_ISL_3101551, EPI_ISL_3101552, EPI_ISL_3101553, EPI_ISL_3101554, EPI_ISL_3101555, EPI_ISL_3101556, EPI_ISL_3101557, EPI_ISL_3101558, EPI_ISL_3101559, EPI_ISL_3101560, EPI_ISL_3101561, EPI_ISL_3101562, EPI_ISL_3101563, EPI_ISL_3101564, EPI_ISL_3101565, EPI_ISL_3101566, EPI_ISL_3101567, EPI_ISL_3101568, EPI_ISL_3101569, EPI_ISL_3101570, EPI_ISL_5510280, EPI_ISL_5510282, EPI_ISL_5510301, EPI_ISL_5510305, EPI_ISL_5510341, EPI_ISL_5510421, EPI_ISL_5510474, EPI_ISL_5510481 | | | |
| see above | TAMBO MEMORIAL | National Institute for Communicable Diseases of the National Health Laboratory Service | Amoako DG; Bhiman JN; Everatt J; Ismail A; Mahlangu B; Mnguni A; Mohale T; Ntuli N; Scheepers C; Sisonke Team |
| EPI_ISL_2153480, EPI_ISL_2153481, EPI_ISL_2153482, EPI_ISL_2662486, EPI_ISL_2662487, EPI_ISL_2662488, EPI_ISL_2662489, EPI_ISL_2662490, EPI_ISL_2662491, EPI_ISL_2662556, EPI_ISL_2662557, EPI_ISL_2662558, EPI_ISL_2662559, EPI_ISL_2662560, EPI_ISL_2662561, EPI_ISL_2662562, EPI_ISL_2662563, EPI_ISL_2662564, EPI_ISL_2662565, EPI_ISL_2662566, EPI_ISL_2662567, EPI_ISL_2662568, EPI_ISL_2662569, EPI_ISL_2662570, EPI_ISL_2662571, EPI_ISL_2662572, EPI_ISL_2662573, EPI_ISL_2662574, EPI_ISL_2662575, EPI_ISL_2662576, EPI_ISL_2662577, EPI_ISL_2662578, EPI_ISL_2662579, EPI_ISL_2662580, EPI_ISL_2662581, EPI_ISL_2662582, EPI_ISL_2662583, EPI_ISL_2662584, EPI_ISL_2662585, EPI_ISL_2662586, EPI_ISL_2662587, EPI_ISL_2662588, EPI_ISL_2662589, EPI_ISL_2662590, EPI_ISL_2662591, EPI_ISL_2662592, EPI_ISL_2662593, EPI_ISL_2662594, EPI_ISL_2662595, EPI_ISL_2662596, EPI_ISL_2662597, EPI_ISL_2662598, EPI_ISL_2662599, EPI_ISL_2662600, EPI_ISL_2662601, EPI_ISL_2662602, EPI_ISL_2662603, EPI_ISL_2662604, EPI_ISL_2662605, EPI_ISL_2662606, EPI_ISL_2662607, EPI_ISL_2662608, EPI_ISL_2662609, EPI_ISL_2662610, EPI_ISL_2662611, EPI_ISL_2662612, EPI_ISL_2662613, EPI_ISL_2662614, EPI_ISL_2662615, EPI_ISL_2662616, EPI_ISL_2662617, EPI_ISL_2695772, EPI_ISL_2695773, EPI_ISL_2695774, EPI_ISL_2695775, EPI_ISL_2695776, EPI_ISL_2695777, EPI_ISL_2695778, EPI_ISL_2695779, EPI_ISL_2695780, EPI_ISL_2695781, EPI_ISL_2695782, EPI_ISL_2695783, EPI_ISL_2695784, EPI_ISL_2709943, EPI_ISL_2709944, EPI_ISL_2709945, EPI_ISL_2709946, EPI_ISL_2709947, EPI_ISL_2709948, EPI_ISL_2709949, EPI_ISL_2709950, EPI_ISL_2709951, EPI_ISL_2709952, EPI_ISL_2709953, EPI_ISL_2709954, EPI_ISL_2709955, EPI_ISL_2709956, EPI_ISL_2709957, EPI_ISL_2709958, EPI_ISL_2709959, EPI_ISL_2709960, EPI_ISL_2709961, EPI_ISL_2709962, EPI_ISL_2709963, EPI_ISL_2709964, EPI_ISL_2709965, EPI_ISL_2709966, EPI_ISL_2709967, EPI_ISL_2709968, EPI_ISL_2709969, EPI_ISL_2709970, EPI_ISL_2709972, EPI_ISL_2709973, EPI_ISL_2709974, EPI_ISL_2709975, EPI_ISL_2709976, EPI_ISL_2709977, EPI_ISL_2709979, EPI_ISL_2709980, EPI_ISL_2709981, EPI_ISL_2709982, EPI_ISL_2709983, EPI_ISL_2709984, EPI_ISL_2709985, EPI_ISL_2709986, EPI_ISL_2709987, EPI_ISL_2709988, EPI_ISL_2709989, EPI_ISL_2709990, EPI_ISL_2709991, EPI_ISL_2709992, EPI_ISL_2709993, EPI_ISL_2709994, EPI_ISL_2827037, EPI_ISL_2827038, EPI_ISL_2827039, EPI_ISL_2827040, EPI_ISL_2827041, EPI_ISL_2827042, EPI_ISL_2827043, EPI_ISL_2827044, EPI_ISL_2827045, EPI_ISL_2827046, EPI_ISL_2827047, EPI_ISL_2827048, EPI_ISL_2827049, EPI_ISL_2827050, EPI_ISL_2827051, EPI_ISL_2827052, EPI_ISL_2827053, EPI_ISL_2827054, EPI_ISL_2827055, EPI_ISL_2827056, EPI_ISL_2827057, EPI_ISL_2827058, EPI_ISL_2827059, EPI_ISL_2827060, EPI_ISL_2827061, EPI_ISL_2827062, EPI_ISL_2827063, EPI_ISL_2827064, EPI_ISL_2827065, EPI_ISL_2827066, EPI_ISL_2827067, EPI_ISL_2827068, EPI_ISL_2827069, EPI_ISL_2827142, EPI_ISL_2827143, EPI_ISL_2827144, EPI_ISL_2827145, EPI_ISL_2827146, EPI_ISL_2827147, EPI_ISL_2827148, EPI_ISL_2827149, EPI_ISL_2827150, EPI_ISL_2827151, EPI_ISL_2827152, EPI_ISL_2827153, EPI_ISL_2827154, EPI_ISL_2827155, EPI_ISL_3024627, EPI_ISL_3024628, EPI_ISL_3024629, EPI_ISL_3024630, EPI_ISL_3024631, EPI_ISL_3024632, EPI_ISL_3024633, EPI_ISL_3024634, EPI_ISL_3024635, EPI_ISL_3024636, EPI_ISL_3024637, EPI_ISL_3024638, EPI_ISL_3024639, EPI_ISL_3024640, EPI_ISL_3024641, EPI_ISL_3024642, EPI_ISL_3074030, EPI_ISL_3074032, EPI_ISL_3074033, EPI_ISL_3074034, EPI_ISL_3074035, EPI_ISL_3074036, EPI_ISL_3074037, EPI_ISL_3074038, EPI_ISL_3074039, EPI_ISL_3074040, EPI_ISL_3074041, EPI_ISL_3236935, EPI_ISL_3236936, EPI_ISL_3236937, EPI_ISL_3236938, EPI_ISL_3236939, EPI_ISL_3236940, EPI_ISL_3236941, EPI_ISL_3236942, EPI_ISL_3236943, EPI_ISL_3236944, EPI_ISL_3236945, EPI_ISL_3236946, EPI_ISL_3236948, EPI_ISL_3236949, EPI_ISL_3236950, EPI_ISL_3236951, EPI_ISL_3236952, EPI_ISL_3236953, EPI_ISL_3236954, EPI_ISL_3237059, EPI_ISL_3237060, EPI_ISL_3237061, EPI_ISL_3237062, EPI_ISL_3237063, EPI_ISL_3237064, EPI_ISL_3237065, EPI_ISL_3237066, EPI_ISL_3237067, EPI_ISL_3237068, EPI_ISL_3237069, EPI_ISL_3237070, EPI_ISL_3237071, EPI_ISL_3237072, EPI_ISL_3237073, EPI_ISL_3237074, EPI_ISL_3237075, EPI_ISL_3237076, EPI_ISL_3237077, EPI_ISL_3237078, EPI_ISL_3237079, EPI_ISL_3237080, EPI_ISL_3237081, EPI_ISL_3237082, EPI_ISL_3237083, EPI_ISL_3237084, EPI_ISL_3237085, EPI_ISL_3237086, EPI_ISL_3237087, EPI_ISL_3237088, EPI_ISL_3237089, EPI_ISL_3237090, EPI_ISL_3237091, EPI_ISL_3237092, EPI_ISL_3237093, EPI_ISL_3237094, EPI_ISL_3237095, EPI_ISL_3237096, EPI_ISL_3237097, EPI_ISL_3237098, EPI_ISL_3237099, EPI_ISL_3237100, EPI_ISL_3237101, EPI_ISL_3237102, EPI_ISL_3237103, EPI_ISL_3342257, EPI_ISL_3342258, EPI_ISL_3342259, EPI_ISL_3342260, EPI_ISL_3342261, EPI_ISL_3342262, EPI_ISL_3342263, EPI_ISL_3342264, EPI_ISL_3342265, EPI_ISL_3411468, EPI_ISL_3451184, EPI_ISL_3451185, EPI_ISL_3451186, EPI_ISL_3451187, EPI_ISL_3451188, EPI_ISL_3451189, EPI_ISL_3451190, EPI_ISL_3451191, EPI_ISL_3451192, EPI_ISL_3451193, EPI_ISL_3451194, EPI_ISL_3451195, EPI_ISL_3451196, EPI_ISL_3451197, EPI_ISL_3451198, EPI_ISL_3451199, EPI_ISL_3451200, EPI_ISL_3451201, EPI_ISL_3451202, EPI_ISL_3451203, EPI_ISL_3451204, EPI_ISL_3451205, EPI_ISL_3451206, EPI_ISL_3451207, EPI_ISL_3451208, EPI_ISL_3451209, EPI_ISL_3451210, EPI_ISL_3451211, EPI_ISL_3451212, EPI_ISL_3451213, EPI_ISL_3451214, EPI_ISL_3451215, EPI_ISL_3451216, EPI_ISL_3451505, EPI_ISL_3451506, EPI_ISL_3451507, EPI_ISL_3451508, EPI_ISL_3451509, EPI_ISL_3451510, EPI_ISL_3451511, EPI_ISL_3451512, EPI_ISL_3451513, EPI_ISL_3451514, EPI_ISL_3451515, EPI_ISL_3451516, EPI_ISL_3451517, EPI_ISL_3451518, EPI_ISL_3451519, EPI_ISL_3451520, EPI_ISL_3451521, EPI_ISL_3451522, EPI_ISL_3451523, EPI_ISL_3451524, EPI_ISL_3451525, EPI_ISL_3451526, EPI_ISL_3451527, EPI_ISL_3451528, EPI_ISL_3451529, EPI_ISL_3717901, EPI_ISL_3717929, EPI_ISL_3717931, EPI_ISL_3717952, EPI_ISL_3717955, EPI_ISL_3717961, EPI_ISL_3717964, EPI_ISL_3717998, EPI_ISL_3718033, EPI_ISL_3718034, EPI_ISL_5196436, EPI_ISL_5196518, EPI_ISL_5196531, EPI_ISL_5196555, EPI_ISL_5196563, EPI_ISL_5196564, EPI_ISL_5196582, EPI_ISL_5196611, EPI_ISL_5196625, EPI_ISL_5416267, EPI_ISL_5416269, EPI_ISL_5416272, EPI_ISL_5416277, EPI_ISL_5416280, EPI_ISL_5416281, EPI_ISL_5416285, EPI_ISL_5416287, EPI_ISL_5416304, EPI_ISL_5416305, EPI_ISL_5416313, EPI_ISL_5416320, EPI_ISL_5416325, EPI_ISL_5416328, EPI_ISL_5416331, EPI_ISL_5416363, EPI_ISL_5416365, EPI_ISL_5416473, EPI_ISL_5416474, EPI_ISL_5416477, EPI_ISL_5416478, EPI_ISL_5416479, EPI_ISL_5416480, EPI_ISL_5416481, EPI_ISL_5416482, EPI_ISL_5416483, EPI_ISL_5416484, EPI_ISL_5416485, EPI_ISL_5416486, EPI_ISL_5416487, EPI_ISL_5416488, EPI_ISL_5416489, EPI_ISL_5416490, EPI_ISL_5416491, EPI_ISL_5416492, EPI_ISL_5416493, EPI_ISL_5416494, EPI_ISL_5416495, EPI_ISL_5416496, EPI_ISL_5416497, EPI_ISL_5416498, EPI_ISL_5416499, EPI_ISL_5416500, EPI_ISL_5416501, EPI_ISL_5416502, EPI_ISL_5416503, EPI_ISL_5416504, EPI_ISL_5416505, EPI_ISL_5416506, EPI_ISL_5416507, EPI_ISL_5603066, EPI_ISL_5603067, EPI_ISL_5603068, EPI_ISL_5603069, EPI_ISL_5603070, EPI_ISL_5603074, EPI_ISL_5603085, EPI_ISL_5603133, EPI_ISL_5603137, EPI_ISL_5603138, EPI_ISL_5603145, EPI_ISL_5603153, EPI_ISL_5603160, EPI_ISL_5603162, EPI_ISL_5603168, EPI_ISL_5603173, EPI_ISL_5603174, EPI_ISL_5603179, EPI_ISL_5603189, EPI_ISL_6201577, EPI_ISL_6202177, EPI_ISL_6202187, EPI_ISL_6202234, EPI_ISL_6202310, EPI_ISL_6202316, EPI_ISL_6202339, EPI_ISL_6202387, EPI_ISL_6202400, EPI_ISL_6202430, EPI_ISL_6202435, EPI_ISL_6202453, EPI_ISL_6202476, EPI_ISL_6202480, EPI_ISL_6202495, EPI_ISL_6202503, EPI_ISL_6202511, EPI_ISL_6202519, EPI_ISL_6202531, EPI_ISL_6202556, EPI_ISL_6202565, EPI_ISL_6202574, EPI_ISL_6202583, EPI_ISL_6202597, EPI_ISL_6202606, EPI_ISL_6202617, EPI_ISL_6202630, EPI_ISL_6202657, EPI_ISL_6202661, EPI_ISL_6202749, EPI_ISL_6202812, EPI_ISL_6203025, EPI_ISL_6203207, EPI_ISL_6203304, EPI_ISL_6203356, EPI_ISL_6203359, EPI_ISL_6203417, EPI_ISL_6203461, EPI_ISL_6203484, EPI_ISL_6203593, EPI_ISL_6203605, EPI_ISL_6900134, EPI_ISL_6900135, EPI_ISL_6906374, EPI_ISL_6906375, EPI_ISL_6906383 | | | |
| see above | TAMBO MEMORIAL LABORATORY | National Institute for Communicable Diseases of the National Health Laboratory Service | Amoako DG; Bhiman JN; Everatt J; Ismail A; Mahlangu B; Mnguni A; Mohale T; Ntuli N; Scheepers C; Wolter N |

| | | | |
|---|---|---|---|
| EPI_ISL_5510487 | THUTHUKANI CLINIC | National Institute for Communicable Diseases of the National Health Laboratory Service | Amoako DG; Bhiman JN; Everatt J; Ismail A; Mahlangu B; Mnguni A; Mohale T; Ntuli N; Scheepers C |
| EPI_ISL_3236977, EPI_ISL_3236978, EPI_ISL_3237116, EPI_ISL_3237135, EPI_ISL_3237136, EPI_ISL_3237137, EPI_ISL_3237138, EPI_ISL_5196441, EPI_ISL_5196442, EPI_ISL_5196528, EPI_ISL_7971571, EPI_ISL_7971580 | | | |
| see above | TSHEPONG LABORATORY | National Institute for Communicable Diseases of the National Health Laboratory Service | Amoako DG; Bhiman JN; Everatt J; Ismail A; Kekana D; Mahlangu B; Mnguni A; Mohale T; Ntuli N; Scheepers C; Wolter N |
| EPI_ISL_5510389 | TSHWANE ACADEMIC | National Institute for Communicable Diseases of the National Health Laboratory Service | Amoako DG; Bhiman JN; Everatt J; Ismail A; Mahlangu B; Mnguni A; Mohale T; Ntuli N; Scheepers C |
| EPI_ISL_2285301, EPI_ISL_2285302, EPI_ISL_2285304, EPI_ISL_2285305, EPI_ISL_2285306, EPI_ISL_2285307, EPI_ISL_2285308, EPI_ISL_2285309, EPI_ISL_2285310, EPI_ISL_2285311, EPI_ISL_2285312, EPI_ISL_2285313, EPI_ISL_3838566, EPI_ISL_3838567, EPI_ISL_3838568, EPI_ISL_4651617, EPI_ISL_4651628, EPI_ISL_4651636, EPI_ISL_4651647, EPI_ISL_4651651, EPI_ISL_4651659, EPI_ISL_4651677, EPI_ISL_4651704, EPI_ISL_4651724, EPI_ISL_4651726, EPI_ISL_4651752, EPI_ISL_4651755, EPI_ISL_4651787, EPI_ISL_4651810, EPI_ISL_4651898, EPI_ISL_4651906, EPI_ISL_4651919, EPI_ISL_4651984, EPI_ISL_4651996, EPI_ISL_4652005, EPI_ISL_4652015, EPI_ISL_4652027, EPI_ISL_4652041, EPI_ISL_4652046, EPI_ISL_4652055, EPI_ISL_4652062, EPI_ISL_4652065, EPI_ISL_4652284 | | | |
| see above | TSHWANE ACADEMIC LABORATORY | National Institute for Communicable Diseases of the National Health Laboratory Service | Amoako DG; Bhiman JN; Everatt J; Ismail A; Mahlangu B; Mnguni A; Mohale T; Ntuli N; Scheepers C |
| EPI_ISL_3342730, EPI_ISL_3342731, EPI_ISL_3342732, EPI_ISL_3342733, EPI_ISL_5510313, EPI_ISL_5510314, EPI_ISL_5510362, EPI_ISL_5510374, EPI_ISL_5510386, EPI_ISL_5510439, EPI_ISL_5510455 | | | |
| see above | Tambo Memorial | National Institute for Communicable Diseases of the National Health Laboratory Service | Amoako DG; Bhiman JN; Everatt J; Ismail A; Mahlangu B; Mnguni A; Mohale T; Ntuli N; Scheepers C |
| EPI_ISL_5917846, EPI_ISL_5917847, EPI_ISL_5917852, EPI_ISL_5917854, EPI_ISL_5917858, EPI_ISL_5917866, EPI_ISL_5917868, EPI_ISL_5917881, EPI_ISL_5917892, EPI_ISL_5917895, EPI_ISL_5917896, EPI_ISL_5917936, EPI_ISL_5917950, EPI_ISL_5917967, EPI_ISL_5917968, EPI_ISL_5917973, EPI_ISL_5917977, EPI_ISL_5917978, EPI_ISL_5917985, EPI_ISL_5918003, EPI_ISL_5918022, EPI_ISL_5918023, EPI_ISL_5918024, EPI_ISL_5918026, EPI_ISL_5918033, EPI_ISL_5918038, EPI_ISL_5918040, EPI_ISL_5918046, EPI_ISL_5918048, EPI_ISL_5918051, EPI_ISL_5918053, EPI_ISL_5918054, EPI_ISL_5918056, EPI_ISL_5918066, EPI_ISL_5918071, EPI_ISL_5918080, EPI_ISL_5918085, EPI_ISL_5918088, EPI_ISL_5918098, EPI_ISL_5918103, EPI_ISL_5918104, EPI_ISL_5918149, EPI_ISL_5918150, EPI_ISL_5918152, EPI_ISL_5918153, EPI_ISL_5919754, EPI_ISL_5919765, EPI_ISL_5919775, EPI_ISL_5919777, EPI_ISL_5919782, EPI_ISL_5919787, EPI_ISL_5919790, EPI_ISL_5919855, EPI_ISL_5919861, EPI_ISL_5919870, EPI_ISL_5919878, EPI_ISL_5919888, EPI_ISL_5919895, EPI_ISL_5919902, EPI_ISL_5919910, EPI_ISL_5919913, EPI_ISL_5919917, EPI_ISL_6913967, EPI_ISL_6913968, EPI_ISL_6913969, EPI_ISL_6913970, EPI_ISL_6913971, EPI_ISL_6913972, EPI_ISL_6913973, EPI_ISL_6913974, EPI_ISL_6913975, EPI_ISL_6913976, EPI_ISL_6913977, EPI_ISL_6913978, EPI_ISL_6913979, EPI_ISL_6913981, EPI_ISL_6913982, EPI_ISL_6913983, EPI_ISL_6913984, EPI_ISL_6913985, EPI_ISL_6913987, EPI_ISL_6913988, EPI_ISL_6913989, EPI_ISL_6913990, EPI_ISL_7337382, EPI_ISL_7337383, EPI_ISL_7337384, EPI_ISL_7337390, EPI_ISL_7337391, EPI_ISL_7337392, EPI_ISL_7337393, EPI_ISL_7337394, EPI_ISL_7337395, EPI_ISL_7337396, EPI_ISL_7337397, EPI_ISL_7337398, EPI_ISL_7337399, EPI_ISL_7337400, EPI_ISL_7337401, EPI_ISL_7337402, EPI_ISL_7337403, EPI_ISL_7337404, EPI_ISL_7337405, EPI_ISL_7337406, EPI_ISL_7337407, EPI_ISL_7337408, EPI_ISL_7337409, EPI_ISL_7337410, EPI_ISL_7337411, EPI_ISL_7337412, EPI_ISL_7337413, EPI_ISL_7337414, EPI_ISL_7337415, EPI_ISL_7337416, EPI_ISL_7456405, EPI_ISL_7456406, EPI_ISL_7456407, EPI_ISL_7456408, EPI_ISL_7456409, EPI_ISL_7456410, EPI_ISL_7456417, EPI_ISL_7456419, EPI_ISL_7456420, EPI_ISL_7456421, EPI_ISL_7456422, EPI_ISL_7456425, EPI_ISL_7456426, EPI_ISL_7456427, EPI_ISL_7456428, EPI_ISL_7456430, EPI_ISL_7456432, EPI_ISL_7456433, EPI_ISL_7456434, EPI_ISL_7456435, EPI_ISL_7456436, EPI_ISL_7456437, EPI_ISL_7456438, EPI_ISL_7456439, EPI_ISL_7456440, EPI_ISL_7456441, EPI_ISL_7456442, EPI_ISL_7456445, EPI_ISL_7456446 | | | |
| see above | Tambo Memorial Laboratory | National Institute for Communicable Diseases of the National Health Laboratory Service | Amoako DG; Bhiman JN; Everatt J; Ismail A; Mahlangu B; Mnguni A; Mohale T; Ntuli N; Scheepers C; Wolter N |
| EPI_ISL_5510390 | Tembisa | National Institute for Communicable Diseases of the National Health Laboratory Service | Amoako DG; Bhiman JN; Everatt J; Ismail A; Mahlangu B; Mnguni A; Mohale T; Ntuli N; Scheepers C |
| EPI_ISL_2271958, EPI_ISL_2271959, EPI_ISL_2271960, EPI_ISL_2271961, EPI_ISL_2271963, EPI_ISL_2271969, EPI_ISL_2271975, EPI_ISL_2271985, EPI_ISL_2272009, EPI_ISL_2272012 | | | |
| see above | Tshepong Hospital | National Institute for Communicable Diseases of the National Health Laboratory Service | Amoako DG; Bhiman JN; Ismail A; Mahlangu B; Mohale T; Ntuli N; Scheepers C |
| EPI_ISL_5918113 | Tshepong Laboratory | National Institute for Communicable Diseases of the National Health Laboratory Service | Amoako DG; Bhiman JN; Everatt J; Ismail A; Mahlangu B; Mnguni A; Mohale T; Ntuli N; Scheepers C |
| EPI_ISL_1324121, EPI_ISL_1324126 | UW Virology Lab | UW Virology Lab | Alexander Greninger; Hong Xie; Keith R Jerome; Lasata Shrestha; Margaret Mills; Meei-Li Huang; Michelle Lin; Noah Baker; Pavitra Roychoudhury; Saraswathi Sathees; Sean Ellis; Shah Mohamed Bakhash |
| EPI_ISL_3237038, EPI_ISL_3237039, EPI_ISL_3237043, EPI_ISL_3237044, EPI_ISL_3237243, EPI_ISL_3237244, EPI_ISL_3237251, EPI_ISL_3237252, EPI_ISL_3237253, EPI_ISL_3411587, EPI_ISL_3411588, EPI_ISL_3411589, EPI_ISL_3411590, EPI_ISL_3411591, EPI_ISL_3411592, EPI_ISL_3411593, EPI_ISL_3411594, EPI_ISL_3411595, EPI_ISL_3411596, EPI_ISL_3411597, EPI_ISL_3411598, EPI_ISL_3411599, EPI_ISL_3411600, EPI_ISL_3411601, EPI_ISL_3411602, EPI_ISL_3411603, EPI_ISL_3411604, EPI_ISL_3411605, EPI_ISL_3411606, EPI_ISL_3411607, EPI_ISL_3411608, EPI_ISL_5510431 | | | |
| see above | University Pretoria | National Institute for Communicable Diseases of the National Health Laboratory Service | Adriano Mendes; Amoako DG; Amy Strydom; Bhiman JN; Everatt J; Ismail A; Mahlangu B; Marietjie Venter; Micheala Davids; Mnguni A; Mohale T; Ntuli N; Scheepers C; Sim Mayaphi |
| EPI_ISL_6308308, EPI_ISL_6308309, EPI_ISL_6308310, EPI_ISL_6308311, EPI_ISL_6308315, EPI_ISL_6308316, EPI_ISL_6308320, EPI_ISL_6308327, EPI_ISL_6308336, EPI_ISL_6308340, EPI_ISL_6308341, EPI_ISL_6308351, EPI_ISL_6308356 | | | |
| see above | University of Pretoria | CERI, Centre for Epidemic Response and Innvoation, Stellenbosch University and KRISP, KZN Research Innovation and Sequencing Platform, UKZN. | Adriano Mendes; Amy Strydom; Giandhari Jennifer; Marietjie Venter; Micheala Davids; Naidoo Yeshnee; Pillay Sureshnee; San James; Sim Mayaphi; Tegally Houriiyah; Tshiabuila Derek; Wilkinson Eduan; Yajna Ramphal; de Oliveira Tulio |
| EPI_ISL_2382656, EPI_ISL_2382657, EPI_ISL_2382658, EPI_ISL_2382659, EPI_ISL_2382660, EPI_ISL_2382661, EPI_ISL_2382662, EPI_ISL_2382664, EPI_ISL_2382665, EPI_ISL_2382666, EPI_ISL_2382667, EPI_ISL_2382668, EPI_ISL_2382670, EPI_ISL_2382671, EPI_ISL_2382673 | | | |
| see above | VERMAAK | National Institute for Communicable Diseases of the National Health Laboratory Service | Amoako DG; Bhiman JN; Ismail A; Mahlangu B; Mohale T; Ntuli N; Scheepers C |
| EPI_ISL_2662476, EPI_ISL_2662477, EPI_ISL_2662478, EPI_ISL_2662479, EPI_ISL_2662480, EPI_ISL_2662482, EPI_ISL_2662483, EPI_ISL_2662484, EPI_ISL_2662485, EPI_ISL_2662530, EPI_ISL_2662537, EPI_ISL_7740759, EPI_ISL_7740763, EPI_ISL_7740766, EPI_ISL_7740854, EPI_ISL_7740857, EPI_ISL_7740858, EPI_ISL_7740859, EPI_ISL_7740860, EPI_ISL_7740861, EPI_ISL_7740866, EPI_ISL_7740885, EPI_ISL_7740889, EPI_ISL_7740890, EPI_ISL_7740897, EPI_ISL_7971522, EPI_ISL_7971523, EPI_ISL_7971524, EPI_ISL_7971525, EPI_ISL_7971526, EPI_ISL_7971527, EPI_ISL_7971528, EPI_ISL_7971529, EPI_ISL_7971530, EPI_ISL_7971531, EPI_ISL_7971532, EPI_ISL_7971533, EPI_ISL_7971534, EPI_ISL_7971535, EPI_ISL_7971536, EPI_ISL_7971537, EPI_ISL_7971538, EPI_ISL_7971539, EPI_ISL_7971540, EPI_ISL_7971541, EPI_ISL_7971542, EPI_ISL_7971543, EPI_ISL_7971544, EPI_ISL_7971545, EPI_ISL_7971546, EPI_ISL_7971547, EPI_ISL_7971548, EPI_ISL_7971549, EPI_ISL_7971550, EPI_ISL_7971551, EPI_ISL_7971552, EPI_ISL_7971553, EPI_ISL_7971554, EPI_ISL_7971555, EPI_ISL_7971556 | | | |
| see above | VERMAAK LABORATORY | National Institute for Communicable Diseases of the National Health Laboratory Service | Amoako DG; Bhiman JN; Everatt J; Ismail A; Kekana D; Mahlangu B; Mnguni A; Mohale T; Ntuli N; Scheepers C; Wolter N |
| EPI_ISL_2494710, EPI_ISL_2494715, EPI_ISL_2494717, EPI_ISL_2494718, EPI_ISL_2494719, EPI_ISL_2494723, EPI_ISL_2494724, EPI_ISL_2494726, EPI_ISL_2494732, EPI_ISL_2494733, EPI_ISL_2494735, EPI_ISL_2494737, EPI_ISL_2494742, EPI_ISL_2494754, EPI_ISL_2494757, EPI_ISL_2494760, EPI_ISL_2494765, EPI_ISL_2494768, EPI_ISL_2494788, EPI_ISL_2494902, EPI_ISL_2494919, EPI_ISL_2494933, EPI_ISL_2494941, EPI_ISL_2494947, EPI_ISL_2494962, EPI_ISL_2494967, EPI_ISL_2494968, EPI_ISL_2494970, EPI_ISL_2494973, EPI_ISL_2494974, EPI_ISL_2494976, EPI_ISL_2494981, EPI_ISL_2494987, EPI_ISL_2494988, EPI_ISL_2494991, EPI_ISL_2494996, EPI_ISL_2494997, EPI_ISL_2495000, EPI_ISL_2495001, EPI_ISL_2495002, EPI_ISL_2495004, EPI_ISL_2495006, EPI_ISL_2495010, EPI_ISL_2495011, EPI_ISL_2495015, EPI_ISL_2495018, EPI_ISL_2495019, EPI_ISL_2495020, EPI_ISL_2495022, EPI_ISL_2495024, EPI_ISL_2495030, EPI_ISL_2495031, EPI_ISL_2495033, EPI_ISL_3447812, EPI_ISL_3447814, EPI_ISL_3447815, EPI_ISL_3447816, EPI_ISL_3447823, EPI_ISL_3447825, EPI_ISL_3447843, EPI_ISL_3447844, EPI_ISL_3447845, EPI_ISL_3447846, EPI_ISL_3447847, EPI_ISL_3447848, EPI_ISL_3447849, EPI_ISL_3447852, EPI_ISL_3447853, EPI_ISL_3447854, EPI_ISL_3447863, EPI_ISL_3447864, EPI_ISL_3447865, EPI_ISL_3447866, EPI_ISL_3447867, EPI_ISL_3447868, EPI_ISL_3447881, EPI_ISL_3728985, EPI_ISL_3728989, EPI_ISL_3728993, EPI_ISL_3728996, EPI_ISL_3729000, EPI_ISL_3729005, EPI_ISL_3729009, EPI_ISL_3729013, EPI_ISL_3729016, EPI_ISL_3729021, EPI_ISL_3729024, EPI_ISL_3729028, EPI_ISL_3729032, EPI_ISL_3729036, EPI_ISL_3729047, EPI_ISL_3729054, EPI_ISL_3729072, EPI_ISL_3729076, EPI_ISL_3729080, EPI_ISL_3729101, EPI_ISL_3729106, EPI_ISL_3729115, EPI_ISL_3729118, EPI_ISL_3729124, EPI_ISL_3729125, EPI_ISL_3729126, EPI_ISL_3729131, EPI_ISL_3729132, EPI_ISL_3729138, EPI_ISL_3729142, EPI_ISL_3729145, EPI_ISL_3729146, EPI_ISL_3729147, EPI_ISL_3729150, EPI_ISL_3729153, EPI_ISL_3729164, EPI_ISL_3729183, EPI_ISL_3729192, EPI_ISL_3729196, EPI_ISL_3729203, EPI_ISL_3729208, EPI_ISL_3729212, EPI_ISL_3729216, EPI_ISL_3729220, EPI_ISL_3729227, EPI_ISL_3729235 | | | |
| see above | Vaccines and Infectious Diseases Analytics Research Unit (VIDA) | KRISP, KZN Research Innovation and Sequencing Platform | Baillie Vicky; Giandhari Jennifer; Madhi Shabir; Naidoo Yeshnee; Pillay Sureshnee; San James; Tegally Houriiyah; Tshabuila Derek; Wilkinson Eduan; Yajna Ramphal; de Oliveira Tulio; du Plessis Jeanine |
| EPI_ISL_2360362, EPI_ISL_2360371, EPI_ISL_2360376, EPI_ISL_2360393, EPI_ISL_2360399, EPI_ISL_2360402, EPI_ISL_2360407, EPI_ISL_2360410, EPI_ISL_2360414, EPI_ISL_2360415, EPI_ISL_2360416, EPI_ISL_2360417, EPI_ISL_2360468, EPI_ISL_2360519, EPI_ISL_2360520, EPI_ISL_2360521, EPI_ISL_2360522, EPI_ISL_2360523, EPI_ISL_2360531, EPI_ISL_2360533, EPI_ISL_2360536, EPI_ISL_2360537, EPI_ISL_2360538, EPI_ISL_2360539, EPI_ISL_2360540, EPI_ISL_2360541, EPI_ISL_2360542, EPI_ISL_2360544, EPI_ISL_2360545, EPI_ISL_2360546, EPI_ISL_2360548, EPI_ISL_2360549, EPI_ISL_2360555, EPI_ISL_2360556, EPI_ISL_2360558, EPI_ISL_2360560, EPI_ISL_2360561, EPI_ISL_2360562, EPI_ISL_2360564, EPI_ISL_2360567, EPI_ISL_2360568, EPI_ISL_2360569, EPI_ISL_2360577, EPI_ISL_2360578, EPI_ISL_2360579, EPI_ISL_2360582, EPI_ISL_2360583, EPI_ISL_2360584, EPI_ISL_2360585, EPI_ISL_2360588, EPI_ISL_2360590, EPI_ISL_2360599, EPI_ISL_2360604, EPI_ISL_2360605, EPI_ISL_2360610, EPI_ISL_2360611, EPI_ISL_2360615, EPI_ISL_2360618, EPI_ISL_2360619, EPI_ISL_2360622, EPI_ISL_2360623, EPI_ISL_2360624, EPI_ISL_2360625, EPI_ISL_2360626, EPI_ISL_2360627, EPI_ISL_2360629, EPI_ISL_2360630, EPI_ISL_2360648, EPI_ISL_2360650, EPI_ISL_2360656, EPI_ISL_2360657, EPI_ISL_2360668, EPI_ISL_2360672, EPI_ISL_2360673, EPI_ISL_2360674, EPI_ISL_2360685, EPI_ISL_2360686, EPI_ISL_2360688, EPI_ISL_2360691, EPI_ISL_2360696, EPI_ISL_2360700, EPI_ISL_2360710, EPI_ISL_2360715, EPI_ISL_2360720, EPI_ISL_2360727, EPI_ISL_2360728, EPI_ISL_2360729, EPI_ISL_2360734, EPI_ISL_2360747, EPI_ISL_2360748, EPI_ISL_2360749, EPI_ISL_2360750, EPI_ISL_2360751, EPI_ISL_2360752, EPI_ISL_2360753, EPI_ISL_2360754, EPI_ISL_2360755, EPI_ISL_2360756, EPI_ISL_2360757, EPI_ISL_2360759, EPI_ISL_2360760, EPI_ISL_2360767, EPI_ISL_2360769, EPI_ISL_2360770, EPI_ISL_2360777, EPI_ISL_2360779, EPI_ISL_2360780, EPI_ISL_2360781, EPI_ISL_2360790, EPI_ISL_2360791, EPI_ISL_2360793, EPI_ISL_2360798, EPI_ISL_2360799, EPI_ISL_2360801, EPI_ISL_2360802, EPI_ISL_2360804, EPI_ISL_2360805, EPI_ISL_2360806, EPI_ISL_2360807, EPI_ISL_2360808, EPI_ISL_2360810, EPI_ISL_2360811, EPI_ISL_2360812, EPI_ISL_2360813, EPI_ISL_2360814, EPI_ISL_2360815, EPI_ISL_2360819, EPI_ISL_2360824, EPI_ISL_2360827, EPI_ISL_2360830, EPI_ISL_2360831, EPI_ISL_2360832, EPI_ISL_2360834, EPI_ISL_2360835, EPI_ISL_2360855, EPI_ISL_2360857, EPI_ISL_2360859, EPI_ISL_2360860, EPI_ISL_2360861, EPI_ISL_2360867, EPI_ISL_2360869, EPI_ISL_2360870, EPI_ISL_2360873, EPI_ISL_2360874, EPI_ISL_2360875, EPI_ISL_2360879, EPI_ISL_2360880, EPI_ISL_2360884, EPI_ISL_2360891, EPI_ISL_2360901, EPI_ISL_2360906, EPI_ISL_2360911, EPI_ISL_2360912, EPI_ISL_2360913, EPI_ISL_2360922, EPI_ISL_2360927, EPI_ISL_2360930, EPI_ISL_2360931, EPI_ISL_2360932, EPI_ISL_2360933, EPI_ISL_2360937, EPI_ISL_2360939, EPI_ISL_2360945, EPI_ISL_2360946, EPI_ISL_2360947, EPI_ISL_2360952, EPI_ISL_2360953, EPI_ISL_2360956, EPI_ISL_2360957, EPI_ISL_2360958, EPI_ISL_2360959, EPI_ISL_2360960, EPI_ISL_2360961, EPI_ISL_2360965, EPI_ISL_2360967, EPI_ISL_2360968, EPI_ISL_2360969, EPI_ISL_2360970, EPI_ISL_2360971, EPI_ISL_2360972, EPI_ISL_2360975, EPI_ISL_2360977, EPI_ISL_2360978, EPI_ISL_2360980, EPI_ISL_2360988, EPI_ISL_2360989, EPI_ISL_2360990, EPI_ISL_2360991, EPI_ISL_2360992, EPI_ISL_2360994, EPI_ISL_2360995, EPI_ISL_2360996, EPI_ISL_2360997, EPI_ISL_2360998, EPI_ISL_2361005, EPI_ISL_2361006, EPI_ISL_2361007, EPI_ISL_2361008, EPI_ISL_2361010, EPI_ISL_2361012, EPI_ISL_2361015, EPI_ISL_2361016, EPI_ISL_2361017, EPI_ISL_2361019, EPI_ISL_2361020, EPI_ISL_2361021, EPI_ISL_2361022, EPI_ISL_2361023, EPI_ISL_2361025, EPI_ISL_2361029, EPI_ISL_2361030, EPI_ISL_2361031, EPI_ISL_2361032, EPI_ISL_2361033, EPI_ISL_2361034, EPI_ISL_2361035, EPI_ISL_2361046, EPI_ISL_2361059, EPI_ISL_2361062, EPI_ISL_2361063, EPI_ISL_2361064, EPI_ISL_2361065, EPI_ISL_2361066, EPI_ISL_2361068, EPI_ISL_2494357, EPI_ISL_2494359, EPI_ISL_2494360, EPI_ISL_2494365, EPI_ISL_2494366, EPI_ISL_2494367, EPI_ISL_2494371, EPI_ISL_2494372, EPI_ISL_2494373, EPI_ISL_2494378, EPI_ISL_2494379, EPI_ISL_2494380, EPI_ISL_2494381, EPI_ISL_2494382, EPI_ISL_2494388, EPI_ISL_2494391, EPI_ISL_2494392, EPI_ISL_2494394, EPI_ISL_2494396, EPI_ISL_2494398, EPI_ISL_2494402, EPI_ISL_2494405, EPI_ISL_2494407, EPI_ISL_2494409, EPI_ISL_2494412, EPI_ISL_2494413, EPI_ISL_2494416, EPI_ISL_2494420, EPI_ISL_2494423, EPI_ISL_2494425, EPI_ISL_2494426, EPI_ISL_2494429, EPI_ISL_2494430, EPI_ISL_2494432, EPI_ISL_2494433, EPI_ISL_2494434, EPI_ISL_2494435, EPI_ISL_2494436, EPI_ISL_2494438, EPI_ISL_2494439, EPI_ISL_2494442, EPI_ISL_2494443, EPI_ISL_2494444, EPI_ISL_2494448, EPI_ISL_2494456, EPI_ISL_2494460, EPI_ISL_2494461, EPI_ISL_2494466, EPI_ISL_2494468, EPI_ISL_2494471, EPI_ISL_2494472, EPI_ISL_2494478, EPI_ISL_2494479, EPI_ISL_2494482, EPI_ISL_2494484, EPI_ISL_2494487, EPI_ISL_2494488, EPI_ISL_2494489, EPI_ISL_2494494, EPI_ISL_2494495, EPI_ISL_2494512, EPI_ISL_2494513, EPI_ISL_2494526, EPI_ISL_2494528, EPI_ISL_2494529, EPI_ISL_2494534, EPI_ISL_2494537, EPI_ISL_2494539, EPI_ISL_2494549, EPI_ISL_2494552, EPI_ISL_2494553, EPI_ISL_2494558, EPI_ISL_2494559, EPI_ISL_2494562, EPI_ISL_2494563, EPI_ISL_2494567, EPI_ISL_2494575, EPI_ISL_2494577, EPI_ISL_2494578, EPI_ISL_2494580, EPI_ISL_2494582, EPI_ISL_2494586, EPI_ISL_2494589, EPI_ISL_2494590, EPI_ISL_2494591, EPI_ISL_2494593, EPI_ISL_2494594, EPI_ISL_2494601, EPI_ISL_2494603, EPI_ISL_2494609, EPI_ISL_2494616, EPI_ISL_2494627, EPI_ISL_2494630 | | | |
| see above | Vaccines and Infectious Diseases Analytics Research Unit (VIDA) | KRISP, KZn Research Innovation and Sequencing Platform | Baillie Vicky; Giandhari Jennifer; Madhi Shabir; Naidoo Yeshnee; Pillay Sureshnee; San James; Tegally Houriiyah; Tshabuila Derek; Wilkinson Eduan; de Oliveira Tulio; du Plessis Jeanine |
| EPI_ISL_940850, EPI_ISL_940851, EPI_ISL_940852, EPI_ISL_940853, EPI_ISL_940854, EPI_ISL_940855, EPI_ISL_940856, EPI_ISL_940858, EPI_ISL_940859, EPI_ISL_940861, EPI_ISL_940862, EPI_ISL_940863, EPI_ISL_940865, EPI_ISL_940866, EPI_ISL_940867, EPI_ISL_940868, EPI_ISL_940869, EPI_ISL_940870, EPI_ISL_940871, EPI_ISL_940874, EPI_ISL_940875, EPI_ISL_940876, EPI_ISL_940877, EPI_ISL_940878, EPI_ISL_940879, EPI_ISL_940880, EPI_ISL_940881, EPI_ISL_940882, EPI_ISL_940884, EPI_ISL_940885, EPI_ISL_940886, EPI_ISL_940887, EPI_ISL_940888, EPI_ISL_940889, EPI_ISL_940890, EPI_ISL_940891 | | | |
| see above | Vaccines and Infectious Diseases Analytics Research Unit (VIDA) | KRISP, KZN Research Innovation and Sequencing Platform | Baillie Vicky; Giandhari Jennifer; Madhi Shabir; Naidoo Yeshnee; Pillay Sureshnee; Tegally Houriiyah; de Oliveira Tulio; du Plessis Jeanine |
| EPI_ISL_1132669, EPI_ISL_1132670, EPI_ISL_1132671, EPI_ISL_1132672, EPI_ISL_1132673, EPI_ISL_1132674, EPI_ISL_1132675, EPI_ISL_1132676, EPI_ISL_1132677, EPI_ISL_1132678, EPI_ISL_1132679, EPI_ISL_1132680, EPI_ISL_1132681, EPI_ISL_1132682, EPI_ISL_1132683, EPI_ISL_1132684, EPI_ISL_1132685, EPI_ISL_1132686, EPI_ISL_1132687, EPI_ISL_1132688, EPI_ISL_1132689, EPI_ISL_1132690, EPI_ISL_1132691, EPI_ISL_1132692, EPI_ISL_1132693, EPI_ISL_1132694, EPI_ISL_1132695, EPI_ISL_1132696, EPI_ISL_1132697, EPI_ISL_1132698, EPI_ISL_1132699, EPI_ISL_1132700, EPI_ISL_1132701, EPI_ISL_1132702, EPI_ISL_1132703, EPI_ISL_1132704, EPI_ISL_1132705, EPI_ISL_1132706, EPI_ISL_1132707, EPI_ISL_1132708, EPI_ISL_1132709, EPI_ISL_1132710, EPI_ISL_1132711, EPI_ISL_1132712, EPI_ISL_1132713, EPI_ISL_1132714, EPI_ISL_1132715, EPI_ISL_1132716, EPI_ISL_1132717, EPI_ISL_1132718, EPI_ISL_1132719, EPI_ISL_1132720, EPI_ISL_1132721, EPI_ISL_1132722, EPI_ISL_1132723, EPI_ISL_1132724, EPI_ISL_1132725, EPI_ISL_1132726, EPI_ISL_1132727, EPI_ISL_1132728 | | | |
| see above | Vaccines and Infectious Diseases Analytics Research Unit (VIDA) | KRISP, KZn Research Innovation and Sequencing Platform | Baillie Vicky; Giandhari Jennifer; Madhi Shabir; Naidoo Yeshnee; Pillay Sureshnee; Tegally Houriiyah; de Oliveira Tulio; du Plessis Jeanine |
| EPI_ISL_5917944, EPI_ISL_5917979, EPI_ISL_5918019, EPI_ISL_5918027 | Vermaak | National Institute for Communicable Diseases of the National Health Laboratory Service | Amoako DG; Bhiman JN; Everatt J; Ismail A; Mahlangu B; Mnguni A; Mohale T; Ntuli N; Scheepers C |
| EPI_ISL_2086968, EPI_ISL_2086969, EPI_ISL_2086972, EPI_ISL_2086979, EPI_ISL_2086981, EPI_ISL_2086983, EPI_ISL_2582726, EPI_ISL_2582730, EPI_ISL_2582733, EPI_ISL_2582736, EPI_ISL_2582739, EPI_ISL_2582742, EPI_ISL_2582744, EPI_ISL_2582746, EPI_ISL_2582748, EPI_ISL_2582749, EPI_ISL_2582752, EPI_ISL_2582757, EPI_ISL_2582761, EPI_ISL_2582765, EPI_ISL_2582769, EPI_ISL_2582772, EPI_ISL_2582774, EPI_ISL_2582776, EPI_ISL_2582781, EPI_ISL_2582786, EPI_ISL_2582790, EPI_ISL_2582792, EPI_ISL_2582794, EPI_ISL_2582797, EPI_ISL_2582800, EPI_ISL_2582803, EPI_ISL_2582806 | | | |
| see above | Vermaak & Partners Laboratory | National Institute for Communicable Diseases of the National Health Laboratory Service | Amoako DG; Bhiman JN; Everatt J; Ismail A; Mahlangu B; Mnguni A; Mohale T; Ntuli N; Scheepers C |
| EPI_ISL_2841660, EPI_ISL_2841661, EPI_ISL_2841662, EPI_ISL_2841663, EPI_ISL_2841664, EPI_ISL_2841665, EPI_ISL_2841666, EPI_ISL_2841667, EPI_ISL_2841668, EPI_ISL_2841669, EPI_ISL_2841670, EPI_ISL_2841671, EPI_ISL_2841672, EPI_ISL_2841673, EPI_ISL_2841674, EPI_ISL_2841675, EPI_ISL_2841676, EPI_ISL_2841677, EPI_ISL_2841678, EPI_ISL_2841679, EPI_ISL_2841680, EPI_ISL_2841681, EPI_ISL_2841682, EPI_ISL_2841683, EPI_ISL_2841684, EPI_ISL_2841685, EPI_ISL_2841686, EPI_ISL_2841687, EPI_ISL_2841688, EPI_ISL_2841689, EPI_ISL_2841690, EPI_ISL_2841691, EPI_ISL_2841692, EPI_ISL_2841693, EPI_ISL_2841694, EPI_ISL_2841695, EPI_ISL_2841696, EPI_ISL_2841697, EPI_ISL_2841698, EPI_ISL_2841699, EPI_ISL_2841700 | | | |
| see above | Wits Vaccines & Infectious Diseases Analytics (VIDA) Research Unit | KRISP, KZn Research Innovation and Sequencing Platform | Baillie Vicky; Giandhari Jennifer; Madhi Shabir; Naidoo Yeshnee; Pillay Sureshnee; San James; Tegally Houriiyah; Wilkinson Eduan; de Oliveira Tulio; du Plessis Jeanine |

| | | | |
|---|---|---|---|
| EPI_ISL_6261993 | ZARV, Department Medical Virology, University of Pretoria | ZARV, Department Mdeical Virology, University of Pretoria | Adriano Mendes; Amy Strydom; Katja Koeppel; Lia Rotherham and Marietjie Venter |
| EPI_ISL_6261983, EPI_ISL_6261987, EPI_ISL_6261989, EPI_ISL_6261996 | ZARV, Department Medical Virology, University of Pretoria | ZARV, Department Medical Virology, University of Pretoria | Adriano Mendes; Amy Strydom; Katja Koeppel; Lia Rotherham and Marietjie Venter |
| EPI_ISL_4474416, EPI_ISL_4474417, EPI_ISL_4474418, EPI_ISL_4474419, EPI_ISL_4474420, EPI_ISL_4474421, EPI_ISL_4474422, EPI_ISL_4474423, EPI_ISL_4474424, EPI_ISL_4474425, EPI_ISL_4474426, EPI_ISL_4474427, EPI_ISL_4474428, EPI_ISL_4474429, EPI_ISL_4474430, EPI_ISL_4474431, EPI_ISL_4474432 | | | |
| see above | ZARV/NHLS, Department Medical Virology, University of Pretoria | CERI, Centre for Epidemic Response and Innovation | Adriano Mendes; Amy Strydom; Emmanuel S; Giandhari J; Micheala Davids; Naidoo Yeshnee; Pillay S; Sim Mayaphi and Marietjie Venter; Tegally H; Tshabuila Derek; Wilkinson E; Yajna Ramphal; de Oliveira T |
| EPI_ISL_5098761, EPI_ISL_5098766, EPI_ISL_5098771, EPI_ISL_5098774, EPI_ISL_5098782, EPI_ISL_5098787, EPI_ISL_5098801, EPI_ISL_5098860, EPI_ISL_5098870, EPI_ISL_5098876, EPI_ISL_5098882, EPI_ISL_5098912, EPI_ISL_5098921, EPI_ISL_5098949, EPI_ISL_5098984, EPI_ISL_5098990, EPI_ISL_5099007, EPI_ISL_5099083, EPI_ISL_6795833, EPI_ISL_6795834, EPI_ISL_6795835, EPI_ISL_6795836, EPI_ISL_6795837, EPI_ISL_6795838, EPI_ISL_6795839, EPI_ISL_6795840, EPI_ISL_6795841, EPI_ISL_6795842, EPI_ISL_6795843, EPI_ISL_6795844, EPI_ISL_6795845, EPI_ISL_6795846, EPI_ISL_6795847, EPI_ISL_6795848, EPI_ISL_6795849, EPI_ISL_6795850, EPI_ISL_6825389, EPI_ISL_6825390, EPI_ISL_6825391, EPI_ISL_6825392, EPI_ISL_6825393, EPI_ISL_6825394, EPI_ISL_6825395, EPI_ISL_6825396, EPI_ISL_6825397, EPI_ISL_6825398, EPI_ISL_7015171, EPI_ISL_7015172, EPI_ISL_7015173, EPI_ISL_7015174, EPI_ISL_7015175, EPI_ISL_7015176, EPI_ISL_7015177, EPI_ISL_7015178, EPI_ISL_7015179, EPI_ISL_7015180, EPI_ISL_7015181, EPI_ISL_7015182, EPI_ISL_7015183, EPI_ISL_7015184, EPI_ISL_7015185, EPI_ISL_7015186, EPI_ISL_7015187, EPI_ISL_7015188, EPI_ISL_7015189, EPI_ISL_7015190, EPI_ISL_7015191, EPI_ISL_7015192, EPI_ISL_7015193, EPI_ISL_7015194, EPI_ISL_7015195, EPI_ISL_7015196, EPI_ISL_7015197, EPI_ISL_7015198, EPI_ISL_7015199, EPI_ISL_7015200, EPI_ISL_7015201, EPI_ISL_7015202, EPI_ISL_7015203, EPI_ISL_7015204, EPI_ISL_7015205, EPI_ISL_7015206, EPI_ISL_7015207, EPI_ISL_7015208, EPI_ISL_8128432, EPI_ISL_8128433, EPI_ISL_8128434, EPI_ISL_8128435, EPI_ISL_8128436, EPI_ISL_8128437, EPI_ISL_8128438, EPI_ISL_8128439, EPI_ISL_8128440, EPI_ISL_8128441, EPI_ISL_8128442, EPI_ISL_8128443, EPI_ISL_8128444, EPI_ISL_8128445, EPI_ISL_8128446, EPI_ISL_8128447, EPI_ISL_8128448, EPI_ISL_8128449, EPI_ISL_8128450, EPI_ISL_8128451, EPI_ISL_8128452, EPI_ISL_8128453, EPI_ISL_8128454, EPI_ISL_8128455, EPI_ISL_8128456, EPI_ISL_8128457, EPI_ISL_8128458, EPI_ISL_8128459, EPI_ISL_8128460, EPI_ISL_8128461, EPI_ISL_8128462, EPI_ISL_8128463, EPI_ISL_8128464, EPI_ISL_8128465, EPI_ISL_8128466, EPI_ISL_8128467, EPI_ISL_8128468, EPI_ISL_8128469, EPI_ISL_8128470, EPI_ISL_8128471, EPI_ISL_8128472, EPI_ISL_8128473, EPI_ISL_8128474, EPI_ISL_8128475, EPI_ISL_8128476, EPI_ISL_8128477, EPI_ISL_8128478, EPI_ISL_8128479, EPI_ISL_8128480, EPI_ISL_8128481, EPI_ISL_8128482, EPI_ISL_8128483, EPI_ISL_8128484, EPI_ISL_8128485, EPI_ISL_8128486, EPI_ISL_8128487, EPI_ISL_8128488, EPI_ISL_8128489, EPI_ISL_8128490, EPI_ISL_8128491, EPI_ISL_8128492, EPI_ISL_8128493, EPI_ISL_8128494, EPI_ISL_8128495, EPI_ISL_8128496, EPI_ISL_8128497, EPI_ISL_8128498, EPI_ISL_8128499, EPI_ISL_8128500, EPI_ISL_8128501, EPI_ISL_8128502, EPI_ISL_8128503, EPI_ISL_8128504 | | | |
| see above | ZARV/NHLS, Department Medical Virology, University of Pretoria | CERI, Centre for Epidemic Response and Innvoation, Stellenbosch University and KRISP, KZN Research Innovation and Sequencing Platform, UKZN. | Adriano Mendes; Amoaka D; Amy Strydom; Arisha Maharaj; Bester P; Bhiman J; Engelbrecht S; Everatt J; Giandhari J; Giandhari Jennifer; Goedhals D; Hardie D; Hsiao M; Iranzadeh A; Lessells R; Makatini Z; Maponga T; Mdlalose N; Micheala Davids; Mlisana K; Moir M; NGS-SA (Scheepers C; Naidoo Y; Naidoo Yeshnee; Nyaga M) Giandhari J; Oluwakemi M; Pillay S; Pillay Sureshnee; Preiser W; Ramphal U; Ramphal Y; San JE; San James; Sim Mayaphi and Marietjie Venter; Tegally H; Tegally Houriiyah; Tshiabuila D; Tshiabuila Derek; Upasana Ramphal; Venter M; Wilkinson E; Wilkinson Eduan; Williamson C; Yajna Ramphal; de Oliveira T; de Oliveira Tulio; von Gottberg A |
| EPI_ISL_2727239, EPI_ISL_2727240, EPI_ISL_2727241, EPI_ISL_2727242, EPI_ISL_2727243, EPI_ISL_2727244, EPI_ISL_2727245, EPI_ISL_2727246, EPI_ISL_2727247, EPI_ISL_2727248, EPI_ISL_2727249, EPI_ISL_2727250, EPI_ISL_2727251, EPI_ISL_2727252, EPI_ISL_2727253, EPI_ISL_2727254, EPI_ISL_2727255, EPI_ISL_2727256, EPI_ISL_2727257, EPI_ISL_2727258, EPI_ISL_2727259, EPI_ISL_2727260, EPI_ISL_2727261, EPI_ISL_2727262, EPI_ISL_2727263, EPI_ISL_2727264, EPI_ISL_2727265, EPI_ISL_2727266, EPI_ISL_2727267, EPI_ISL_2727268, EPI_ISL_2727269, EPI_ISL_2727270, EPI_ISL_2727271, EPI_ISL_2727272, EPI_ISL_2727273, EPI_ISL_2727274, EPI_ISL_2727275, EPI_ISL_2727276, EPI_ISL_2727277, EPI_ISL_2727278, EPI_ISL_2727279, EPI_ISL_2727280, EPI_ISL_2727281, EPI_ISL_2727282, EPI_ISL_2727283, EPI_ISL_2727284, EPI_ISL_2727285, EPI_ISL_2727286, EPI_ISL_2727287, EPI_ISL_2727288, EPI_ISL_2727289, EPI_ISL_2727290, EPI_ISL_4572282, EPI_ISL_4572283, EPI_ISL_4572284, EPI_ISL_4572285, EPI_ISL_4572286, EPI_ISL_4572287, EPI_ISL_4572288, EPI_ISL_4572289, EPI_ISL_4572290, EPI_ISL_4572291, EPI_ISL_4572292, EPI_ISL_4572293, EPI_ISL_4572294, EPI_ISL_4572295, EPI_ISL_4572319, EPI_ISL_4572322, EPI_ISL_4572323, EPI_ISL_4572324, EPI_ISL_4572329, EPI_ISL_4572331, EPI_ISL_4572333, EPI_ISL_4572334, EPI_ISL_4572338, EPI_ISL_4572339, EPI_ISL_4572342, EPI_ISL_4572343, EPI_ISL_4572353, EPI_ISL_4572356, EPI_ISL_4572357, EPI_ISL_4572358, EPI_ISL_4572360, EPI_ISL_4572361, EPI_ISL_4572364, EPI_ISL_4572366, EPI_ISL_4572367, EPI_ISL_4572368, EPI_ISL_4572373, EPI_ISL_4572374, EPI_ISL_4572378, EPI_ISL_4572379, EPI_ISL_4572387, EPI_ISL_4572388, EPI_ISL_4575028, EPI_ISL_4575029, EPI_ISL_4575030, EPI_ISL_4575031, EPI_ISL_4575032, EPI_ISL_4575033, EPI_ISL_4575034, EPI_ISL_4575035, EPI_ISL_4575036, EPI_ISL_4575037, EPI_ISL_4575038, EPI_ISL_4575039, EPI_ISL_4575040, EPI_ISL_4575041, EPI_ISL_4575042, EPI_ISL_4575043, EPI_ISL_4575044, EPI_ISL_4575045, EPI_ISL_4575046, EPI_ISL_4575047, EPI_ISL_4575048, EPI_ISL_4575049, EPI_ISL_4575050, EPI_ISL_4575051, EPI_ISL_4575052, EPI_ISL_4575053, EPI_ISL_4575054, EPI_ISL_4575060, EPI_ISL_4575070, EPI_ISL_4575071, EPI_ISL_4575079, EPI_ISL_4575080, EPI_ISL_4575081, EPI_ISL_4575082, EPI_ISL_4575084, EPI_ISL_4575088, EPI_ISL_4575089, EPI_ISL_4575098, EPI_ISL_4575100, EPI_ISL_4575102, EPI_ISL_4575107, EPI_ISL_4575119, EPI_ISL_4575120, EPI_ISL_4575121, EPI_ISL_4575127, EPI_ISL_4575128, EPI_ISL_4575129, EPI_ISL_4575130, EPI_ISL_4575133, EPI_ISL_4575141, EPI_ISL_4575142, EPI_ISL_4575150, EPI_ISL_4575151, EPI_ISL_4575153, EPI_ISL_4575154, EPI_ISL_4575155, EPI_ISL_4575156, EPI_ISL_4575157, EPI_ISL_4575164, EPI_ISL_4575165, EPI_ISL_4575166, EPI_ISL_4575167, EPI_ISL_4575169, EPI_ISL_4575170, EPI_ISL_4575171, EPI_ISL_4575183, EPI_ISL_4575184, EPI_ISL_4575185, EPI_ISL_4575189, EPI_ISL_4575190, EPI_ISL_4575191, EPI_ISL_4575200, EPI_ISL_4575203 | | | |
| see above | ZARV/NHLS, Department Medical Virology, University of Pretoria | KRISP, KZN Research Innovation and Sequencing Platform | Adriano Mendes; Amy Strydom; Emmanuel SJ; Giandhari J; Giandhari Jennifer; Lessells R; Micheala Davids; Naidoo Y; Naidoo Yeshnee; Pillay S; Pillay Sureshnee; Ramphal U; San James; Sim Mayaphi and Marietjie Venter; Tegally H; Tegally Houriiyah; Tshiabuila Derek; Wilkinson E; Wilkinson Eduan; Yajna Ramphal; de Oliveira T; de Oliveira Tulio |
| EPI_ISL_2727328, EPI_ISL_2727329, EPI_ISL_2727330, EPI_ISL_2727331, EPI_ISL_2727332, EPI_ISL_2727333, EPI_ISL_2727334, EPI_ISL_2727335, EPI_ISL_2727336, EPI_ISL_2727337, EPI_ISL_2727338, EPI_ISL_2727339, EPI_ISL_2727340, EPI_ISL_2727341, EPI_ISL_2727342, EPI_ISL_2727343, EPI_ISL_2727344, EPI_ISL_2727345, EPI_ISL_2727346, EPI_ISL_2727347, EPI_ISL_2727348, EPI_ISL_2727349, EPI_ISL_2727350, EPI_ISL_2727351, EPI_ISL_2727352, EPI_ISL_2727353, EPI_ISL_2727354, EPI_ISL_2727355, EPI_ISL_2727356, EPI_ISL_2727357, EPI_ISL_2727358, EPI_ISL_2727359, EPI_ISL_2727360, EPI_ISL_2727361, EPI_ISL_2727362, EPI_ISL_2727363, EPI_ISL_2727364, EPI_ISL_2727365, EPI_ISL_2727366, EPI_ISL_2727367, EPI_ISL_2727368, EPI_ISL_2727369, EPI_ISL_2727370, EPI_ISL_2727371, EPI_ISL_2727372, EPI_ISL_2727373, EPI_ISL_2727374, EPI_ISL_2727375, EPI_ISL_2727376, EPI_ISL_2727377, EPI_ISL_2727378, EPI_ISL_2727379, EPI_ISL_2899741, EPI_ISL_2899742, EPI_ISL_2899743, EPI_ISL_2899744, EPI_ISL_2899745, EPI_ISL_2899746, EPI_ISL_2899747, EPI_ISL_2899748, EPI_ISL_2899749, EPI_ISL_2899750, EPI_ISL_2899751, EPI_ISL_2899752, EPI_ISL_2899753, EPI_ISL_2899754, EPI_ISL_2899755, EPI_ISL_2899756, EPI_ISL_2899757, EPI_ISL_2955421, EPI_ISL_2955422, EPI_ISL_2955423, EPI_ISL_2955424, EPI_ISL_2955425, EPI_ISL_2955426, EPI_ISL_2955427, EPI_ISL_2955428, EPI_ISL_2955429, EPI_ISL_2955430, EPI_ISL_2955431, EPI_ISL_2955432, EPI_ISL_2955433, EPI_ISL_2955434, EPI_ISL_2955435, EPI_ISL_2955436, EPI_ISL_2955437, EPI_ISL_2955438, EPI_ISL_2955439, EPI_ISL_2955440, EPI_ISL_2955441, EPI_ISL_2955442, EPI_ISL_2955443, EPI_ISL_2955444, EPI_ISL_2955445, EPI_ISL_2955446, EPI_ISL_2955447, EPI_ISL_2955448, EPI_ISL_2955449, EPI_ISL_2955450, EPI_ISL_2955451, EPI_ISL_2955452, EPI_ISL_4121618, EPI_ISL_4121678, EPI_ISL_4121684 | | | |
| see above | ZARV/NHLS, Department Medical Virology, University of Pretoria | KRISP, KZn Research Innovation and Sequencing Platform | Adriano Mendes; Amy Strydom; Emmanuel SJ; Giandhari J; Lessells R; Micheala Davids; Naidoo Y; Pillay S; Ramphal U; Sim Mayaphi and Marietjie Venter; Tegally H; Wilkinson E; de Oliveira T |
| EPI_ISL_5510403 | ZONE 3 CLINIC | National Institute for Communicable Diseases of the National Health Laboratory Service | Amoako DG; Bhiman JN; Everatt J; Ismail A; Mahlangu B; Mnguni A; Mohale T; Ntuli N; Scheepers C |
