## Supplemental Table 4 for "The spread of SARS-CoV-2 variant Omicron with the doubling time of 2.0–3.3 days can be explained by immune evasion"

We gratefully acknowledge the following Authors from the Originating laboratories responsible for obtaining the specimens, as well as the Submitting laboratories where the genome data were generated and shared via GISAID, on which this research is based.

All Submitters of data may be contacted directly via www.gisaid.org

Authors are sorted alphabetically.

| Accession ID | Originating Laboratory | Submitting Laboratory | Authors |
|---|---|---|---|
| EPI_ISL_7381191 | AHRI | CERI, Centre for Epidemic Response and Innvoation, Stellenbosch University and KRISP, KZN Research Innovation and Sequencing Platform, UKZN. | Arisha Maharaj; Bernstein Mallory; Cele Sandile; Giandhari J; Karim Farina; Khan Khadija; Moir M; Naidoo Y; Pillay S; Ramphal U; Ramphal Y; San JE; Sigal Alex; Tegally H; Tshiabuila D; Wilkinson E; de Oliveira T; van Wyk S |
| EPI_ISL_2397308, EPI_ISL_2397309, EPI_ISL_2397310, EPI_ISL_2397311, EPI_ISL_2397312, EPI_ISL_2397313 | AHRI | KRISP, KZN Research Innovation and Sequencing Platform | Bernstein M; COMMIT-KZN Team; Cele S; Giandhari J; Gosnell BI; Hanekom W; Karim F; Khan K; Lessells RJ; Manickchund N; Mlisana K; Moosa MYS; Msomi N; Pillay S; Ramphal U; San JE; Sigal A; Singh L; Tegally H; Wilkinson E; de Oliveira T |
| EPI_ISL_3722334, EPI_ISL_3722335, EPI_ISL_3722336, EPI_ISL_3722337, EPI_ISL_3722338, EPI_ISL_3722339 | AHRI | KRISP, KZn Research Innovation and Sequencing Platform | Bernstein Mallory; Cele Sandile; Emmanuel S; Giandhari J; Karim Farina; Khan Khadija; Naidoo Yeshnee; Pillay S; Sigal Alex; Tegally H; Tshabuila Derek; Wilkinson E; Yajna Ramphal; de Oliveira T |
| EPI_ISL_3939064, EPI_ISL_3939068, EPI_ISL_3939088 | AHRI - Sigal Lab | KRISP, KZn Research Innovation and Sequencing Platform | Bernstein Mallory; Cele Sandile; Emmanuel S; Giandhari J; Karim Farina; Khan Khadija; Naidoo Yeshnee; Pillay S; Sigal Alex; Tegally H; Tshabuila Derek; Wilkinson E; Yajna Ramphal; de Oliveira T |
| EPI_ISL_3447712, EPI_ISL_3447752, EPI_ISL_3447779, EPI_ISL_3447802 | AHRI- Alex Sigal Lab | KRISP, KZN Research Innovation and Sequencing Platform | Alex Sigal; Giandhari Jennifer; Mallory Bernstein; Naidoo Yeshnee; Pillay Sureshnee; San E. James; Sandile Cele; Tegally Houriiyah; Tshabuila Derek; Wilkinson Eduan; Yajna Ramphal; de Oliveira Tulio |
| EPI_ISL_3267757 | AHRI- Alex Sigal lab | KRISP, KZN Research Innovation and Sequencing Platform | Alex Sigal; Giandhari Jennifer; Mallory Bernstein; Naidoo Yeshnee; Pillay Sureshnee; San E. James; Sandile Cele; Tegally Houriiyah; Tshabuila Derek; Wilkinson Eduan; Yajna Ramphal; de Oliveira Tulio |
| EPI_ISL_1229367, EPI_ISL_1229368 | AHRI-Sigal | KRISP, KZn Research Innovation and Sequencing Platform | Cele S; Gazy I; Giandhari J; Karim F; Pillay S; Sigal A; Tegally H; Wilkinson E; de Oliveira T |
| EPI_ISL_2493041 | AMPATH | CERI, Centre for Epidemic Response and Innvoation, Stellenbosch University and KRISP, KZN Research Innovation and Sequencing Platform, UKZN. | Emmanuel SJ; Giandhari J; Khan S; Lessells R; Mdlalose K; Naidoo Y; Pillay S; Ramphal U; Tegally H; Wilkinson E; York D; de Oliveira T |
| EPI_ISL_2501090 | AMPATH | KRISP, KZN Research Innovation and Sequencing Platform | Emmanuel SJ; Giandhari J; Khan S; Lessells R; Mdlalose K; Naidoo Y; Pillay S; Ramphal U; Sisonke Team; Tegally H; Wilkinson E; York D; de Oliveira T |
| EPI_ISL_3838572, EPI_ISL_3838573, EPI_ISL_3838574, EPI_ISL_3838575, EPI_ISL_3838576, EPI_ISL_3838577, EPI_ISL_4651553, EPI_ISL_4651560, EPI_ISL_6327732 | | | |
| see above | AMPATH | National Institute for Communicable Diseases of the National Health Laboratory Service | Amoako DG; Bhiman JN; Everatt J; Ismail A; Mahlangu B; Mnguni A; Mohale T; Ntuli N; Scheepers C |
| EPI_ISL_2494961 | Ampath | KRISP, KZN Research Innovation and Sequencing Platform | Emmanuel SJ; Giandhari J; Khan S; Lessells R; Mdlalose K; Naidoo Y; Pillay S; Ramphal U; Tegally H; Wilkinson E; York D; de Oliveira T |
| EPI_ISL_1994608 | Ampath - Kingsway | KRISP, KZN Research Innovation and Sequencing Platform | Emmanuel SJ; Giandhari J; Khan S; Lessells R; Mdlalose K; Naidoo Y; Pillay S; Ramphal U; Tegally H; Wilkinson E; York D; de Oliveira T |
| EPI_ISL_2693088 | Ampath laboratories- Sharks | KRISP, KZN Research Innovation and Sequencing Platform | Giandhari Jennifer; Naidoo Yeshnee; Pillay Sureshnee; San James; Tegally Houriiyah; Tshabuila Derek; Wilkinson Eduan; Yajna Ramphal; de Oliveira Tulio |
| EPI_ISL_860553 | Ampath-Netcare | KRISP, KZn Research Innovation and Sequencing Platform | Giandhari J; Khan S; Lessells R; Mdlalose K; Pillay S; Tegally H; Wilkinson E; York D; de Oliveira T |
| EPI_ISL_2893315, EPI_ISL_2893316 | BARC - Sisonke | KRISP, KZN Research Innovation and Sequencing Platform | Giandhari Jennifer; Naidoo Yeshnee; Pillay Sureshnee; San James; Sisonke; Tegally Houriiyah; Tshabuila Derek; Wilkinson Eduan; Yajna Ramphal; de Oliveira Tulio |
| EPI_ISL_2955501, EPI_ISL_2955502, EPI_ISL_2955503, EPI_ISL_2955504, EPI_ISL_2955505, EPI_ISL_2955506, EPI_ISL_2955507, EPI_ISL_2955508, EPI_ISL_2955509, EPI_ISL_2955510, EPI_ISL_2955511 | | | |
| see above | CAPRISA | KRISP, KZn Research Innovation and Sequencing Platform | Emmanuel SJ; Giandhari J; Lessells R; Naidoo Y; Ngcapu S; Pillay S; Ramphal U; Samsunder N; Sivro A; Tegally H; Wilkinson E; de Oliveira T |
| EPI_ISL_7747505, EPI_ISL_7747506, EPI_ISL_7747507, EPI_ISL_7747508, EPI_ISL_7747509, EPI_ISL_7747510, EPI_ISL_7747511, EPI_ISL_7747512, EPI_ISL_7747513, EPI_ISL_7747514, EPI_ISL_7747515, EPI_ISL_7747516, EPI_ISL_7747517, EPI_ISL_7747518, EPI_ISL_7747519, EPI_ISL_7747520, EPI_ISL_7747521, EPI_ISL_7747522, EPI_ISL_7747523, EPI_ISL_7747524, EPI_ISL_7747525, EPI_ISL_7747526, EPI_ISL_7747527, EPI_ISL_7747528, EPI_ISL_7747529, EPI_ISL_7747530, EPI_ISL_7747531, EPI_ISL_7747532, EPI_ISL_7747533, EPI_ISL_7747534 | | | |
| see above | CAPRISA_HILLCREST | CERI, Centre for Epidemic Response and Innvoation, Stellenbosch University and KRISP, KZN Research Innovation and Sequencing Platform, UKZN. | Aida Sivro; Arisha Maharaj; Giandhari J; Naidoo Y; Natasha Samsunder; Pillay S; Ramphal U; Ramphal Y; San JE; Tegally H; Tshiabuila D; Wilkinson E; de Oliveira T |
| EPI_ISL_2271972, EPI_ISL_2271982, EPI_ISL_2271990, EPI_ISL_2271991, EPI_ISL_2272019, EPI_ISL_2272020, EPI_ISL_2272021 | | | |
| see above | Edendale Gateway Clinic | National Institute for Communicable Diseases of the National Health Laboratory Service | Amoako DG; Bhiman JN; Ismail A; Mahlangu B; Mohale T; Ntuli N; Scheepers C |
| EPI_ISL_4253672 | Edendale Hosiptal | National Institute for Communicable Diseases of the National Health Laboratory Service | Amoako DG; Bhiman JN; Everatt J; Ismail A; Mahlangu B; Mnguni A; Mohale T; Ntuli N; Scheepers C; Sisonke Team |
| EPI_ISL_2086267 | Enthabeni Lancet Laboratories | KRISP, KZN Research Innovation and Sequencing Platform | Emmanuel SJ; Giandhari J; Khan S; Lessells R; Mdlalose K; Naidoo Y; Pillay S; Ramphal U; Tegally H; Wilkinson E; York D; de Oliveira T |
| EPI_ISL_5416464 | KIMBERLEY LABORATORY | National Institute for Communicable Diseases of the National Health Laboratory Service | Amoako DG; Bhiman JN; Everatt J; Ismail A; Mahlangu B; Mnguni A; Mohale T; Ntuli N; Scheepers C |
| EPI_ISL_660221, EPI_ISL_660222, EPI_ISL_660228, EPI_ISL_660229, EPI_ISL_660230, EPI_ISL_660231, EPI_ISL_660232, EPI_ISL_660233, EPI_ISL_660256, EPI_ISL_660257, EPI_ISL_660258, EPI_ISL_660664 | | | |
| see above | KRISP, KZN Research Innovation and Sequencing Platform | KRISP, KZN Research Innovation and Sequencing Platform | Giandhari J; Khan S; Lessells R; Mdlalose K; Pillay S; Tegally H; Wilkinson E; York D; de Oliveira T |
| EPI_ISL_2688547, EPI_ISL_2688548 | KRISP, KZn Research Innovation and Sequencing Platform | KRISP, KZn Research Innovation and Sequencing Platform | Giandhari Jennifer; Naidoo Yeshnee; Pillay Sureshnee; San James; Tegally Houriiyah; Tshabuila Derek; Wilkinson Eduan; Yajna Ramphal; de Oliveira Tulio |
| EPI_ISL_5918117 | Kimberley Laboratory | National Institute for Communicable Diseases of the National Health Laboratory Service | Amoako DG; Bhiman JN; Everatt J; Ismail A; Mahlangu B; Mnguni A; Mohale T; Ntuli N; Scheepers C |
| EPI_ISL_2013037, EPI_ISL_2013040 | Lancet Laboratories | National Institute for Communicable Diseases of the National Health Laboratory Service | Amoako DG; Bhiman JN; Glass A; Gottberg A; Mahlangu B; Mohale T; Ntuli N; Oliveira TD; Scheepers C; Tegally H; Viana R |
| EPI_ISL_736927, EPI_ISL_736930, EPI_ISL_736966, EPI_ISL_736967, EPI_ISL_736968, EPI_ISL_736995 | MDS | KRISP, KZn Research Innovation and Sequencing Platform | ChimukangaraB; Giandhari J; Khan S; Lessells R; Mdlalose K; Pillay S; Tegally H; Wilkinson E; York D; de Oliveira T |
| EPI_ISL_660259, EPI_ISL_660260, EPI_ISL_660261, EPI_ISL_660262, EPI_ISL_660263 | Molecualr Diagnostic Services (MDS) | KRISP, KZN Research Innovation and Sequencing Platform | Giandhari J; Khan S; Lessells R; Mdlalose K; Pillay S; Tegally H; Wilkinson E; York D; de Oliveira T |
| EPI_ISL_1250507, EPI_ISL_1250509, EPI_ISL_1250512, EPI_ISL_1250514, EPI_ISL_1250515, EPI_ISL_1250517, EPI_ISL_1250518, EPI_ISL_1250520, EPI_ISL_1250522, EPI_ISL_1250523, EPI_ISL_1250525, EPI_ISL_1250526, EPI_ISL_1250528, EPI_ISL_1250529, EPI_ISL_1250531, EPI_ISL_1250533, EPI_ISL_1250535, EPI_ISL_1250537, EPI_ISL_1250538, EPI_ISL_1250540, EPI_ISL_1250542, EPI_ISL_1250543, EPI_ISL_1250545, EPI_ISL_1250547, EPI_ISL_1250548, EPI_ISL_1250550, EPI_ISL_1250551, EPI_ISL_1250552 | | | |
| see above | Molecular Diagnostics Services (MDS) | KRISP, KZN Research Innovation and Sequencing Platform | Emmanuel SJ; Giandhari J; Khan S; Lessells R; Maslo C; Mdlalose K; Pillay S; Sitharam L; Tegally H; Wilkinson E; York D; de Oliveira T |
| EPI_ISL_529720, EPI_ISL_529721, EPI_ISL_529722, EPI_ISL_529723, EPI_ISL_529724, EPI_ISL_529725, EPI_ISL_529726, EPI_ISL_529727, EPI_ISL_529728, EPI_ISL_529729, EPI_ISL_529730, EPI_ISL_529731, EPI_ISL_529732, EPI_ISL_529733, EPI_ISL_529734, EPI_ISL_529735, EPI_ISL_529736, EPI_ISL_529737, EPI_ISL_529738, EPI_ISL_529739, EPI_ISL_529740, EPI_ISL_529741, EPI_ISL_529742, EPI_ISL_529743, EPI_ISL_529744, EPI_ISL_529745, EPI_ISL_529746, EPI_ISL_529747, EPI_ISL_529748, EPI_ISL_529749, EPI_ISL_529750, EPI_ISL_529751, EPI_ISL_529752, EPI_ISL_529753, EPI_ISL_529754, EPI_ISL_529755, EPI_ISL_529756, EPI_ISL_529757, EPI_ISL_529758, EPI_ISL_529759, EPI_ISL_529760, EPI_ISL_529761, EPI_ISL_529762, EPI_ISL_529763, EPI_ISL_529764, EPI_ISL_529765, EPI_ISL_529766, EPI_ISL_529767, EPI_ISL_529768, EPI_ISL_529769, EPI_ISL_529770, EPI_ISL_529771, EPI_ISL_529772, EPI_ISL_529773, EPI_ISL_529774, EPI_ISL_529775, EPI_ISL_529776, EPI_ISL_529777, EPI_ISL_529778, EPI_ISL_529779, EPI_ISL_529780, EPI_ISL_529781, EPI_ISL_529782, EPI_ISL_529783, EPI_ISL_529784, EPI_ISL_529785, EPI_ISL_529786, EPI_ISL_529787, EPI_ISL_529788, EPI_ISL_529789, EPI_ISL_529790, EPI_ISL_529791, EPI_ISL_529792, EPI_ISL_529793, EPI_ISL_529794, EPI_ISL_529795, EPI_ISL_529796, EPI_ISL_529797, EPI_ISL_529798, EPI_ISL_529799, EPI_ISL_529800, EPI_ISL_529801, EPI_ISL_529802, EPI_ISL_529803, EPI_ISL_535390, EPI_ISL_535391, EPI_ISL_535392, EPI_ISL_535393, EPI_ISL_535394, EPI_ISL_535395, EPI_ISL_535396, EPI_ISL_535397, EPI_ISL_535398, EPI_ISL_535399, EPI_ISL_535400, EPI_ISL_535401, EPI_ISL_535402, EPI_ISL_535403, EPI_ISL_535404, EPI_ISL_535503, EPI_ISL_535504, EPI_ISL_535505, EPI_ISL_535508, EPI_ISL_535509, EPI_ISL_535510, EPI_ISL_535516, EPI_ISL_535517, EPI_ISL_535518, EPI_ISL_535520, EPI_ISL_535521, EPI_ISL_535523, EPI_ISL_535524, EPI_ISL_535525, EPI_ISL_535526, EPI_ISL_535527, EPI_ISL_535528, EPI_ISL_535529, EPI_ISL_535531, EPI_ISL_535533, EPI_ISL_535535, EPI_ISL_535538, EPI_ISL_535540, EPI_ISL_535542, EPI_ISL_535544, EPI_ISL_535549, EPI_ISL_535550, EPI_ISL_535551, EPI_ISL_602632, EPI_ISL_602633, EPI_ISL_602634, EPI_ISL_602635, EPI_ISL_602636, EPI_ISL_602637, EPI_ISL_602638, EPI_ISL_602639, EPI_ISL_602640, EPI_ISL_602641, EPI_ISL_602642, EPI_ISL_602643, EPI_ISL_602644, EPI_ISL_602645, EPI_ISL_602646, EPI_ISL_602647, EPI_ISL_602648, EPI_ISL_602649, EPI_ISL_602650, EPI_ISL_602651, EPI_ISL_602652, EPI_ISL_602653, EPI_ISL_602654, EPI_ISL_602655, EPI_ISL_602656, EPI_ISL_602657, EPI_ISL_602658, EPI_ISL_602659, EPI_ISL_602660, EPI_ISL_602661, EPI_ISL_602662, EPI_ISL_602663, EPI_ISL_602664, EPI_ISL_602665, EPI_ISL_602666, EPI_ISL_602667, EPI_ISL_602668, EPI_ISL_602669, EPI_ISL_602670, EPI_ISL_602671, EPI_ISL_602672, EPI_ISL_602673, EPI_ISL_602674, EPI_ISL_602675, EPI_ISL_602676, EPI_ISL_602677, EPI_ISL_602678, EPI_ISL_602679, EPI_ISL_602680, EPI_ISL_602681, EPI_ISL_602682, EPI_ISL_602683, EPI_ISL_602684, EPI_ISL_602685, EPI_ISL_602686, EPI_ISL_602687, EPI_ISL_602688, EPI_ISL_602689, EPI_ISL_602690, EPI_ISL_602691, EPI_ISL_602692, EPI_ISL_602693, EPI_ISL_602694, EPI_ISL_602695, EPI_ISL_602696, EPI_ISL_602697, EPI_ISL_602698, EPI_ISL_602699, EPI_ISL_602700, EPI_ISL_602701, EPI_ISL_602702, EPI_ISL_602703, EPI_ISL_602704, EPI_ISL_602705, EPI_ISL_602706, EPI_ISL_602707, EPI_ISL_602708, EPI_ISL_602709, EPI_ISL_602710, EPI_ISL_602711, EPI_ISL_602712, EPI_ISL_602713, EPI_ISL_602714, EPI_ISL_602715, EPI_ISL_602716, EPI_ISL_602717, EPI_ISL_602718, EPI_ISL_602719, EPI_ISL_602720, EPI_ISL_602721, EPI_ISL_602722, EPI_ISL_602723, EPI_ISL_602724, EPI_ISL_602725, EPI_ISL_602726, EPI_ISL_602727, EPI_ISL_602728, EPI_ISL_602729, EPI_ISL_602730, EPI_ISL_602731, EPI_ISL_602732, EPI_ISL_602733, EPI_ISL_602734, EPI_ISL_602735, EPI_ISL_602736, EPI_ISL_602737, EPI_ISL_602738, EPI_ISL_602739, EPI_ISL_602740, EPI_ISL_602741, EPI_ISL_602742, EPI_ISL_602743, EPI_ISL_602744, EPI_ISL_602745, EPI_ISL_602746, EPI_ISL_602747, EPI_ISL_602748, EPI_ISL_602749, EPI_ISL_602750, EPI_ISL_602751, EPI_ISL_602752, EPI_ISL_602753, EPI_ISL_602754, EPI_ISL_602755, EPI_ISL_602756, EPI_ISL_602757, EPI_ISL_602758, EPI_ISL_602759, EPI_ISL_602760, EPI_ISL_602761, EPI_ISL_602762, EPI_ISL_602763, EPI_ISL_602764, EPI_ISL_602765, EPI_ISL_602766, EPI_ISL_602767, EPI_ISL_602768, EPI_ISL_602769, EPI_ISL_602770, EPI_ISL_602771, EPI_ISL_602772, EPI_ISL_602773, EPI_ISL_602774, EPI_ISL_602775, EPI_ISL_602776, EPI_ISL_602777, EPI_ISL_602778, EPI_ISL_602779, EPI_ISL_602780, EPI_ISL_602781, EPI_ISL_602782, EPI_ISL_602783, EPI_ISL_602784, EPI_ISL_602785, EPI_ISL_602786, EPI_ISL_602787, EPI_ISL_602788, EPI_ISL_602789, EPI_ISL_602790, EPI_ISL_602791, EPI_ISL_602792, EPI_ISL_602793, EPI_ISL_602794, EPI_ISL_602795, EPI_ISL_602796, EPI_ISL_602797, EPI_ISL_602798, EPI_ISL_602799, EPI_ISL_602800, EPI_ISL_602801, EPI_ISL_602802, EPI_ISL_602803, EPI_ISL_602804, EPI_ISL_602805, EPI_ISL_602806, EPI_ISL_602807, EPI_ISL_602808, EPI_ISL_602809, EPI_ISL_602810, EPI_ISL_602811, EPI_ISL_602812, EPI_ISL_602813, EPI_ISL_602814, EPI_ISL_602815, EPI_ISL_602816, EPI_ISL_602817, EPI_ISL_602818, EPI_ISL_602819, EPI_ISL_602820, EPI_ISL_602821, EPI_ISL_602822, EPI_ISL_602823, EPI_ISL_602824, EPI_ISL_602825, EPI_ISL_602826, EPI_ISL_602827, EPI_ISL_602828, EPI_ISL_602829, EPI_ISL_602830, EPI_ISL_602831, EPI_ISL_602832, EPI_ISL_602833, EPI_ISL_602834, EPI_ISL_602835, EPI_ISL_602836, EPI_ISL_602837, EPI_ISL_602838, EPI_ISL_602839, EPI_ISL_602840, EPI_ISL_602841, EPI_ISL_602842, EPI_ISL_602843, EPI_ISL_602844, EPI_ISL_602845, EPI_ISL_602846, EPI_ISL_602847, EPI_ISL_602848, EPI_ISL_602849, EPI_ISL_602850, EPI_ISL_602851, EPI_ISL_602852, EPI_ISL_602853, EPI_ISL_602854, EPI_ISL_602855, EPI_ISL_602856, EPI_ISL_602857, EPI_ISL_602858, EPI_ISL_602859, EPI_ISL_602860, EPI_ISL_602861, EPI_ISL_602862, EPI_ISL_602863, EPI_ISL_602864, EPI_ISL_602865, EPI_ISL_602866, EPI_ISL_602867, EPI_ISL_602868, EPI_ISL_602869, EPI_ISL_602870, EPI_ISL_602871, EPI_ISL_602872, EPI_ISL_602873, EPI_ISL_602874, EPI_ISL_602875, EPI_ISL_602876, EPI_ISL_602877, EPI_ISL_602878, EPI_ISL_602879, EPI_ISL_602880, EPI_ISL_602881, EPI_ISL_602882, EPI_ISL_602883, EPI_ISL_602884, EPI_ISL_602885, EPI_ISL_602886, EPI_ISL_602887, EPI_ISL_602888, EPI_ISL_602889, EPI_ISL_602890, EPI_ISL_602891, EPI_ISL_602892, EPI_ISL_602893, EPI_ISL_602894, EPI_ISL_602895, EPI_ISL_602896, EPI_ISL_602897, EPI_ISL_602898, EPI_ISL_602899, EPI_ISL_602900, EPI_ISL_602901, EPI_ISL_602902, EPI_ISL_602903, EPI_ISL_602904, EPI_ISL_602905, EPI_ISL_602906, EPI_ISL_602907, EPI_ISL_602908, EPI_ISL_602909, EPI_ISL_602910, EPI_ISL_602911, EPI_ISL_602912, EPI_ISL_602913, EPI_ISL_602914, EPI_ISL_602915, EPI_ISL_602916, EPI_ISL_602917, EPI_ISL_602918, EPI_ISL_602919, EPI_ISL_602920, EPI_ISL_602921, EPI_ISL_602922, EPI_ISL_602923, EPI_ISL_602924, EPI_ISL_602925, EPI_ISL_602926, EPI_ISL_602927, EPI_ISL_602928, EPI_ISL_602929, EPI_ISL_602930, EPI_ISL_605785, EPI_ISL_605786, EPI_ISL_605787, EPI_ISL_605788, EPI_ISL_605789, EPI_ISL_605790, EPI_ISL_660159, EPI_ISL_660160, EPI_ISL_660161, EPI_ISL_660162, EPI_ISL_660163, EPI_ISL_660164, EPI_ISL_660177, EPI_ISL_660178, EPI_ISL_660179, EPI_ISL_660180, EPI_ISL_660181, EPI_ISL_660182, EPI_ISL_660183, EPI_ISL_660184, EPI_ISL_660185, EPI_ISL_660186, EPI_ISL_660187, EPI_ISL_660188, EPI_ISL_660189, EPI_ISL_660190, EPI_ISL_660191, EPI_ISL_660192, EPI_ISL_660193, EPI_ISL_660194, EPI_ISL_660195, EPI_ISL_660196, EPI_ISL_660197, EPI_ISL_660198, EPI_ISL_660199, EPI_ISL_660200, EPI_ISL_660201, EPI_ISL_660202, EPI_ISL_660203, EPI_ISL_660204, EPI_ISL_660205, EPI_ISL_660206, EPI_ISL_660207, EPI_ISL_660208, EPI_ISL_660209, EPI_ISL_660210, EPI_ISL_660211, EPI_ISL_660212, EPI_ISL_660213, EPI_ISL_660214, EPI_ISL_660215, EPI_ISL_660216, EPI_ISL_660217, EPI_ISL_660218, EPI_ISL_660219, EPI_ISL_660220, EPI_ISL_660223, EPI_ISL_660224, EPI_ISL_660225, EPI_ISL_660226, EPI_ISL_660227, EPI_ISL_660234, EPI_ISL_660235, EPI_ISL_660236, EPI_ISL_660237, EPI_ISL_660238, EPI_ISL_660239, EPI_ISL_660240, EPI_ISL_660241, EPI_ISL_660242, EPI_ISL_660243, EPI_ISL_660244, EPI_ISL_660245, EPI_ISL_660246, EPI_ISL_660247, EPI_ISL_660248, EPI_ISL_660249, EPI_ISL_660250, EPI_ISL_660251, EPI_ISL_660252, EPI_ISL_660253, EPI_ISL_660254, EPI_ISL_660255, EPI_ISL_660616, EPI_ISL_660617, EPI_ISL_660618, EPI_ISL_660619, EPI_ISL_660620, EPI_ISL_660621, EPI_ISL_660622, EPI_ISL_660623, EPI_ISL_660624, EPI_ISL_660625, EPI_ISL_660626, EPI_ISL_660627, EPI_ISL_660628, EPI_ISL_660629, EPI_ISL_660630, EPI_ISL_660631, EPI_ISL_660632, EPI_ISL_660633, EPI_ISL_660634, EPI_ISL_660635, EPI_ISL_660636, EPI_ISL_660637, EPI_ISL_660638, EPI_ISL_660639, EPI_ISL_660640, EPI_ISL_660641, EPI_ISL_660642, EPI_ISL_660643, EPI_ISL_660644, EPI_ISL_660645, EPI_ISL_660646, EPI_ISL_660647, EPI_ISL_660648, EPI_ISL_660649, EPI_ISL_660650, EPI_ISL_660651, EPI_ISL_660652, EPI_ISL_660653, EPI_ISL_660654, EPI_ISL_660655, EPI_ISL_660656, EPI_ISL_660657, EPI_ISL_660658, EPI_ISL_660659, EPI_ISL_660660, EPI_ISL_660661, EPI_ISL_660662, EPI_ISL_660663, EPI_ISL_678570, EPI_ISL_678598, EPI_ISL_678599, EPI_ISL_678600, EPI_ISL_678601, EPI_ISL_678602, EPI_ISL_678603, EPI_ISL_678604, EPI_ISL_678605, EPI_ISL_678606, EPI_ISL_678607, EPI_ISL_678608, EPI_ISL_678609, EPI_ISL_678610, EPI_ISL_678611, EPI_ISL_678612, EPI_ISL_678613, EPI_ISL_678614, EPI_ISL_678615, EPI_ISL_678616, EPI_ISL_678617, EPI_ISL_678618, EPI_ISL_678619, EPI_ISL_678620, EPI_ISL_678621, EPI_ISL_678622, EPI_ISL_678623, EPI_ISL_678624, EPI_ISL_678643, EPI_ISL_736925, EPI_ISL_736926, EPI_ISL_736931, EPI_ISL_736932, EPI_ISL_736933, EPI_ISL_736934, EPI_ISL_736935, EPI_ISL_736940, EPI_ISL_736941, EPI_ISL_736942, EPI_ISL_736943, EPI_ISL_736944, EPI_ISL_736945, EPI_ISL_736946, EPI_ISL_736947, EPI_ISL_736948, EPI_ISL_736949, EPI_ISL_736950, EPI_ISL_736951, EPI_ISL_736952, EPI_ISL_736953, EPI_ISL_736954, EPI_ISL_736955, EPI_ISL_736956, EPI_ISL_736957, EPI_ISL_736958, EPI_ISL_736959, EPI_ISL_736960, EPI_ISL_736984, EPI_ISL_736985, EPI_ISL_736986, EPI_ISL_736987, EPI_ISL_736988, EPI_ISL_736989, EPI_ISL_736990, EPI_ISL_736991, EPI_ISL_736992, EPI_ISL_736993, EPI_ISL_736994, EPI_ISL_825128, EPI_ISL_825129, EPI_ISL_825130, EPI_ISL_825131, EPI_ISL_825132, EPI_ISL_825133, EPI_ISL_825134, EPI_ISL_825135, EPI_ISL_825136, EPI_ISL_825137, EPI_ISL_825138, EPI_ISL_825139, EPI_ISL_825438, EPI_ISL_825439, EPI_ISL_825440, EPI_ISL_825441, EPI_ISL_825442, EPI_ISL_825443, EPI_ISL_825444, EPI_ISL_825445, EPI_ISL_825446, EPI_ISL_825447, EPI_ISL_825448, EPI_ISL_825449, EPI_ISL_825450, EPI_ISL_825451, EPI_ISL_825452, EPI_ISL_825453, EPI_ISL_825454, EPI_ISL_825455, EPI_ISL_825456, EPI_ISL_825457, EPI_ISL_825458, EPI_ISL_825459, EPI_ISL_825460, EPI_ISL_825461, EPI_ISL_825462, EPI_ISL_825463, EPI_ISL_825464, EPI_ISL_825465, EPI_ISL_825466, EPI_ISL_825467, EPI_ISL_825468, EPI_ISL_825469, EPI_ISL_825470, EPI_ISL_825471, EPI_ISL_825472, EPI_ISL_825473, EPI_ISL_825474, EPI_ISL_825475, EPI_ISL_825476, EPI_ISL_825477, EPI_ISL_825478, EPI_ISL_825479, EPI_ISL_825480, EPI_ISL_825481, EPI_ISL_825482, EPI_ISL_825483, EPI_ISL_825484, EPI_ISL_825485, EPI_ISL_825486, EPI_ISL_825487, EPI_ISL_825488, EPI_ISL_825489 | | | |
| see above | NHLS-IALCH | KRISP, KZN Research Innovation and Sequencing Platform | Cele S; ChimukangaraB; Gazy I; Giandhari J; Karim F; Khan S; Lessells R; Mdlalose K; Pillay S; Sigal A; Tegally H; Wilkinson E; York D; de Oliveira T |
| EPI_ISL_860554, EPI_ISL_860555, EPI_ISL_860556, EPI_ISL_860557, EPI_ISL_860558, EPI_ISL_860559, EPI_ISL_860560, EPI_ISL_860561, EPI_ISL_860562, EPI_ISL_860563, EPI_ISL_860564, EPI_ISL_860565, EPI_ISL_860566, EPI_ISL_860567, EPI_ISL_860568, EPI_ISL_860569, EPI_ISL_860570, EPI_ISL_860571, EPI_ISL_860572, EPI_ISL_860573, EPI_ISL_860574, EPI_ISL_860575, EPI_ISL_860576, EPI_ISL_860577, EPI_ISL_860578, EPI_ISL_860579, EPI_ISL_860580, EPI_ISL_860581, EPI_ISL_860582, EPI_ISL_860583, EPI_ISL_860584, EPI_ISL_860585, EPI_ISL_860586, EPI_ISL_860587, EPI_ISL_860588, EPI_ISL_860589, EPI_ISL_860590, EPI_ISL_860591, EPI_ISL_860592, EPI_ISL_860593, EPI_ISL_860594, EPI_ISL_860595, EPI_ISL_860596, EPI_ISL_860597, EPI_ISL_860598, EPI_ISL_860599, EPI_ISL_860600, EPI_ISL_860601, EPI_ISL_860602, EPI_ISL_860603, EPI_ISL_860604, EPI_ISL_860605, EPI_ISL_860606, EPI_ISL_860607, EPI_ISL_860608, EPI_ISL_860609, EPI_ISL_860610, EPI_ISL_860611, EPI_ISL_860612, EPI_ISL_860613, EPI_ISL_860614, EPI_ISL_860615, EPI_ISL_860616, EPI_ISL_860617, EPI_ISL_860618, EPI_ISL_860619, EPI_ISL_860620, EPI_ISL_860621, EPI_ISL_860622, EPI_ISL_860623, EPI_ISL_860624, EPI_ISL_860625, | | | |

| | | | |
|---|---|---|---|
| EPI_ISL_860626, EPI_ISL_860627, EPI_ISL_860628, EPI_ISL_860629, EPI_ISL_860630 | | | |
| see above | NHLS-IALCH | KRISP, KZn Research Innovation and Sequencing Platform | Giandhari J; Khan S; Lessells R; Mdlalose K; Pillay S; Tegally H; Wilkinson E; York D; de Oliveira T |
| EPI_ISL_3447709, EPI_ISL_3447710, EPI_ISL_3447711, EPI_ISL_3447716, EPI_ISL_3447717, EPI_ISL_3447719, EPI_ISL_3447720, EPI_ISL_3447721, EPI_ISL_3447722, EPI_ISL_3447723, EPI_ISL_3447724, EPI_ISL_3447725, EPI_ISL_3447726, EPI_ISL_3447727, EPI_ISL_3447730, EPI_ISL_3447731, EPI_ISL_3447732, EPI_ISL_3447733, EPI_ISL_3447735, EPI_ISL_3447736, EPI_ISL_3447740, EPI_ISL_3447741, EPI_ISL_3447742, EPI_ISL_3447745, EPI_ISL_3447747, EPI_ISL_3447749, EPI_ISL_3447750, EPI_ISL_3447751, EPI_ISL_3447753, EPI_ISL_3447755, EPI_ISL_3447756, EPI_ISL_3447757, EPI_ISL_3447758, EPI_ISL_3447759, EPI_ISL_3447782, EPI_ISL_3447797, EPI_ISL_3447801, EPI_ISL_3447805, EPI_ISL_3447810, EPI_ISL_3447811, EPI_ISL_3447813, EPI_ISL_3447817, EPI_ISL_3447819, EPI_ISL_3447822, EPI_ISL_3447824, EPI_ISL_3447826, EPI_ISL_3447827, EPI_ISL_3447828, EPI_ISL_3447829, EPI_ISL_3447830, EPI_ISL_3447831, EPI_ISL_3447832, EPI_ISL_3447833, EPI_ISL_3447834, EPI_ISL_3447835, EPI_ISL_3447836, EPI_ISL_3447838, EPI_ISL_3447839, EPI_ISL_3447840, EPI_ISL_3447841, EPI_ISL_3447842, EPI_ISL_3447855, EPI_ISL_3447869, EPI_ISL_3447870, EPI_ISL_3447871, EPI_ISL_3447872, EPI_ISL_3447873, EPI_ISL_3447874, EPI_ISL_3447875, EPI_ISL_3447876, EPI_ISL_3447877, EPI_ISL_3447878, EPI_ISL_3447879, EPI_ISL_3447880 | | | |
| see above | NHLS_IALCH | KRISP, KZN Research Innovation and Sequencing Platform | Giandhari Jennifer; Naidoo Yeshnee; Pillay Sureshnee; San James; Tegally Houriiyah; Tshabuila Derek; Tshiabuila Derek; Wilkinson Eduan; Yajna Ramphal; de Oliveira Tulio |
| EPI_ISL_4473168, EPI_ISL_4473194, EPI_ISL_4473195, EPI_ISL_4473198, EPI_ISL_4473201 | NHLS_VIRO | CERI, Centre for Epidemic Response and Innvoation, Stellenbosch University and KRISP, KZN Research Innovation and Sequencing Platform, UKZN. | Giandhari Jennifer; Naidoo Yeshnee; Pillay Sureshnee; San James; Tegally Houriiyah; Tshabuila Derek; Wilkinson Eduan; Yajna Ramphal; de Oliveira Tulio |
| EPI_ISL_2693036, EPI_ISL_2693037, EPI_ISL_2693038, EPI_ISL_2693039, EPI_ISL_2693040, EPI_ISL_2693041, EPI_ISL_2693042, EPI_ISL_2693043, EPI_ISL_2693044, EPI_ISL_2693045, EPI_ISL_2693046, EPI_ISL_2693047, EPI_ISL_2693048, EPI_ISL_2693049, EPI_ISL_2693050, EPI_ISL_2693051, EPI_ISL_2693052, EPI_ISL_2693053, EPI_ISL_2693054, EPI_ISL_2693055, EPI_ISL_2693056, EPI_ISL_2693057, EPI_ISL_2693058, EPI_ISL_2693059, EPI_ISL_2693060, EPI_ISL_2693061, EPI_ISL_2693063, EPI_ISL_2693064, EPI_ISL_2693065, EPI_ISL_2693066, EPI_ISL_2693067, EPI_ISL_2693068, EPI_ISL_2693069, EPI_ISL_2693070, EPI_ISL_2693071, EPI_ISL_2693072, EPI_ISL_2693073, EPI_ISL_2693074, EPI_ISL_2693075, EPI_ISL_2693076, EPI_ISL_2693077, EPI_ISL_2693078, EPI_ISL_2693079, EPI_ISL_2693080, EPI_ISL_2693081, EPI_ISL_2693082, EPI_ISL_2693083, EPI_ISL_2693084, EPI_ISL_2693085, EPI_ISL_2693086, EPI_ISL_2693087, EPI_ISL_2693089, EPI_ISL_2693090, EPI_ISL_2693091, EPI_ISL_2693092, EPI_ISL_2693093, EPI_ISL_2693094, EPI_ISL_2693095, EPI_ISL_3161759, EPI_ISL_3161760, EPI_ISL_3161761, EPI_ISL_3161762, EPI_ISL_3161763, EPI_ISL_3161764, EPI_ISL_3161765, EPI_ISL_3161766, EPI_ISL_3161767, EPI_ISL_3161768, EPI_ISL_3161769, EPI_ISL_3161770, EPI_ISL_3161771, EPI_ISL_3161772, EPI_ISL_3161773, EPI_ISL_3161774, EPI_ISL_3161775, EPI_ISL_3161776, EPI_ISL_3161777, EPI_ISL_3161778, EPI_ISL_3161779, EPI_ISL_3161780, EPI_ISL_3161781, EPI_ISL_3161782, EPI_ISL_3161783, EPI_ISL_3161784, EPI_ISL_3161785, EPI_ISL_3161786, EPI_ISL_3161787, EPI_ISL_3161788, EPI_ISL_3161789, EPI_ISL_3161790, EPI_ISL_3161791, EPI_ISL_3161792, EPI_ISL_3161793, EPI_ISL_3161794, EPI_ISL_3161795, EPI_ISL_3161796, EPI_ISL_3161797, EPI_ISL_3161798, EPI_ISL_3161799, EPI_ISL_3161800, EPI_ISL_3161801, EPI_ISL_3261864, EPI_ISL_3261865, EPI_ISL_3261866, EPI_ISL_3261867, EPI_ISL_3261868, EPI_ISL_3261869, EPI_ISL_3261870, EPI_ISL_3261871, EPI_ISL_3261872, EPI_ISL_3261873, EPI_ISL_3261874, EPI_ISL_3261875, EPI_ISL_3261876, EPI_ISL_3261877, EPI_ISL_3261878, EPI_ISL_3261879, EPI_ISL_3261880, EPI_ISL_3261881, EPI_ISL_3261882, EPI_ISL_3261883, EPI_ISL_3261884, EPI_ISL_3261885, EPI_ISL_3261886, EPI_ISL_3261887, EPI_ISL_3261888, EPI_ISL_3261889, EPI_ISL_3261890, EPI_ISL_3261891, EPI_ISL_3261892, EPI_ISL_3261893, EPI_ISL_3261894, EPI_ISL_3261895, EPI_ISL_3261896, EPI_ISL_3261897, EPI_ISL_3261898, EPI_ISL_3261899, EPI_ISL_3261900, EPI_ISL_3261901, EPI_ISL_3261902, EPI_ISL_3261903, EPI_ISL_3261904, EPI_ISL_3261905, EPI_ISL_3261924, EPI_ISL_3261925, EPI_ISL_3261926, EPI_ISL_3261927, EPI_ISL_3261928, EPI_ISL_3261929, EPI_ISL_3261930, EPI_ISL_3261931, EPI_ISL_3261932, EPI_ISL_3261933, EPI_ISL_3261934, EPI_ISL_3261935, EPI_ISL_3261936, EPI_ISL_3261937, EPI_ISL_3261938, EPI_ISL_3261939, EPI_ISL_3261940, EPI_ISL_3261941, EPI_ISL_3261942, EPI_ISL_3261943, EPI_ISL_3261944, EPI_ISL_3261945, EPI_ISL_3261946, EPI_ISL_3261947, EPI_ISL_3261948, EPI_ISL_3261949, EPI_ISL_3261950, EPI_ISL_3261951, EPI_ISL_3261952, EPI_ISL_3261953, EPI_ISL_3261954, EPI_ISL_3261955, EPI_ISL_3261956, EPI_ISL_3261957, EPI_ISL_3261958, EPI_ISL_3261959, EPI_ISL_3261960, EPI_ISL_3261961, EPI_ISL_3261962, EPI_ISL_3261963, EPI_ISL_3261964, EPI_ISL_3261965, EPI_ISL_3261966, EPI_ISL_3261967, EPI_ISL_3261968, EPI_ISL_3261969, EPI_ISL_3261970, EPI_ISL_3261971, EPI_ISL_3261972, EPI_ISL_3261973, EPI_ISL_3261974, EPI_ISL_3261975, EPI_ISL_3261976, EPI_ISL_3261977, EPI_ISL_3261978, EPI_ISL_3261979, EPI_ISL_3261980, EPI_ISL_3261981, EPI_ISL_3261982, EPI_ISL_3261983 | | | |
| see above | NHLS_VIRO | KRISP, KZN Research Innovation and Sequencing Platform | Giandhari Jennifer; Naidoo Yeshnee; Pillay Sureshnee; San James; Tegally Houriiyah; Tshabuila Derek; Wilkinson Eduan; Yajna Ramphal; de Oliveira Tulio |
| EPI_ISL_2494363, EPI_ISL_2494368, EPI_ISL_2494376, EPI_ISL_2494387, EPI_ISL_2494390, EPI_ISL_2494397, EPI_ISL_2494403, EPI_ISL_2494417, EPI_ISL_2494421, EPI_ISL_2494422, EPI_ISL_2494424, EPI_ISL_2494427, EPI_ISL_2494428, EPI_ISL_2494431, EPI_ISL_2494437, EPI_ISL_2494440, EPI_ISL_2494441, EPI_ISL_2494445, EPI_ISL_2494446, EPI_ISL_2494447, EPI_ISL_2494449, EPI_ISL_2494450, EPI_ISL_2494451, EPI_ISL_2494453, EPI_ISL_2494454, EPI_ISL_2494455, EPI_ISL_2494457, EPI_ISL_2494458, EPI_ISL_2494459, EPI_ISL_2494462, EPI_ISL_2494463, EPI_ISL_2494464, EPI_ISL_2494465, EPI_ISL_2494469, EPI_ISL_2494470, EPI_ISL_2494474, EPI_ISL_2494475, EPI_ISL_2494476, EPI_ISL_2494480, EPI_ISL_2494490, EPI_ISL_2494491, EPI_ISL_2494492, EPI_ISL_2494493, EPI_ISL_2494499, EPI_ISL_2494500, EPI_ISL_2494502, EPI_ISL_2494503, EPI_ISL_2494504, EPI_ISL_2494505, EPI_ISL_2494506, EPI_ISL_2494507, EPI_ISL_2494508, EPI_ISL_2494509, EPI_ISL_2494510, EPI_ISL_2494511, EPI_ISL_2494514, EPI_ISL_2494515, EPI_ISL_2494516, EPI_ISL_2494517, EPI_ISL_2494523, EPI_ISL_2494525, EPI_ISL_2494531, EPI_ISL_2494532, EPI_ISL_2494536, EPI_ISL_2494541, EPI_ISL_2494542, EPI_ISL_2494555, EPI_ISL_2494556, EPI_ISL_2494569, EPI_ISL_2494579, EPI_ISL_2494606, EPI_ISL_2494607, EPI_ISL_2494608, EPI_ISL_2494611, EPI_ISL_2494612, EPI_ISL_2494613, EPI_ISL_2494615, EPI_ISL_2494617, EPI_ISL_2494618, EPI_ISL_2494619, EPI_ISL_2494620, EPI_ISL_2494621, EPI_ISL_2494622, EPI_ISL_2494623, EPI_ISL_2494624, EPI_ISL_2494625, EPI_ISL_2494626, EPI_ISL_2727380, EPI_ISL_2727381, EPI_ISL_2727382, EPI_ISL_2727383, EPI_ISL_2727384, EPI_ISL_2727385, EPI_ISL_2727386, EPI_ISL_2727387, EPI_ISL_2727388, EPI_ISL_2727389, EPI_ISL_2727390, EPI_ISL_2727391, EPI_ISL_2727392, EPI_ISL_2727393, EPI_ISL_2727394, EPI_ISL_2727395, EPI_ISL_2727396, EPI_ISL_2727397, EPI_ISL_2727398, EPI_ISL_2727399, EPI_ISL_2727400, EPI_ISL_2727401, EPI_ISL_2727402, EPI_ISL_2727403 | | | |
| see above | NHLS_VIRO | KRISP, KZn Research Innovation and Sequencing Platform | Baillie Vicky; Giandhari Jennifer; Madhi Shabir; Naidoo Yeshnee; Pillay Sureshnee; San James; Tegally Houriiyah; Tshabuila Derek; Wilkinson Eduan; Yajna Ramphal; de Oliveira Tulio; du Plessis Jeanine |
| EPI_ISL_634978, EPI_ISL_634979, EPI_ISL_634980, EPI_ISL_634981, EPI_ISL_634982, EPI_ISL_634983, EPI_ISL_634984, EPI_ISL_634985, EPI_ISL_634986, EPI_ISL_634987, EPI_ISL_634988, EPI_ISL_634989, EPI_ISL_634990, EPI_ISL_634991, EPI_ISL_634992, EPI_ISL_634993, EPI_ISL_634994, EPI_ISL_634995, EPI_ISL_634996, EPI_ISL_634997, EPI_ISL_634998, EPI_ISL_634999, EPI_ISL_635000, EPI_ISL_635001, EPI_ISL_635002, EPI_ISL_635003, EPI_ISL_635004, EPI_ISL_635005, EPI_ISL_635006, EPI_ISL_635007, EPI_ISL_635008, EPI_ISL_635009, EPI_ISL_635010, EPI_ISL_635011, EPI_ISL_635012, EPI_ISL_635013, EPI_ISL_635014, EPI_ISL_635015, EPI_ISL_635016, EPI_ISL_635017, EPI_ISL_635018, EPI_ISL_635019, EPI_ISL_635020, EPI_ISL_635021, EPI_ISL_635022, EPI_ISL_635023, EPI_ISL_635024, EPI_ISL_635025, EPI_ISL_635026, EPI_ISL_635027, EPI_ISL_635028, EPI_ISL_635029, EPI_ISL_635030, EPI_ISL_635031, EPI_ISL_635032, EPI_ISL_635033, EPI_ISL_635034, EPI_ISL_635035, EPI_ISL_635036, EPI_ISL_635037, EPI_ISL_635038, EPI_ISL_635039, EPI_ISL_635040, EPI_ISL_635041, EPI_ISL_635042, EPI_ISL_635043, EPI_ISL_635044, EPI_ISL_635045, EPI_ISL_635046, EPI_ISL_635047, EPI_ISL_635048, EPI_ISL_635049, EPI_ISL_635050, EPI_ISL_635051, EPI_ISL_635052, EPI_ISL_635053, EPI_ISL_635054, EPI_ISL_635055, EPI_ISL_635056, EPI_ISL_635057, EPI_ISL_635058 | | | |
| see above | National Health Laboratory Service - Inkosi Albert Luthuli Central Hospital (NHLS-IALCH) | KRISP, KZN Research Innovation and Sequencing Platform | Giandhari J; Khan S; Lessells R; Mdlalose K; Pillay S; Tegally H; Wilkinson E; York D; de Oliveira T |
| EPI_ISL_4505703 | National Health Laboratory Service, Eastern Cape, South Africa | KRISP, KZN Research Innovation and Sequencing Platform | Giandhari Jennifer; Naidoo Yeshnee; Oluwakemi Laguda-Akingba; Pillay Sureshnee; San James; Tegally Houriiyah; Tshiabuila Derek; Wilkinson Eduan; Yajna Ramphal; de Oliveira Tulio |
| EPI_ISL_4505635, EPI_ISL_4505636, EPI_ISL_4505637, EPI_ISL_4505638, EPI_ISL_4505639, EPI_ISL_4505640, EPI_ISL_4505641, EPI_ISL_4505642, EPI_ISL_4505643, EPI_ISL_4505644, EPI_ISL_4505645, EPI_ISL_4505646, EPI_ISL_4505647, EPI_ISL_4505648, EPI_ISL_4505649, EPI_ISL_4505650, EPI_ISL_4505651, EPI_ISL_4505652, EPI_ISL_4505653, EPI_ISL_4505654, EPI_ISL_4505655, EPI_ISL_4505656, EPI_ISL_4505657, EPI_ISL_4505658, EPI_ISL_4505659, EPI_ISL_4505660, EPI_ISL_4505661, EPI_ISL_4505662, EPI_ISL_4505663, EPI_ISL_4505664, EPI_ISL_4505665, EPI_ISL_4505666, EPI_ISL_4505667, EPI_ISL_4505668, EPI_ISL_4505669, EPI_ISL_4505670, EPI_ISL_4505671 | | | |
| see above | National Health Laboratory Service, Kwazulu Natal, South Africa | KRISP, KZN Research Innovation and Sequencing Platform | Giandhari Jennifer; Naidoo Yeshnee; Nokukhanya Mdlalose; Pillay Sureshnee; San James; Tegally Houriiyah; Tshiabuila Derek; Wilkinson Eduan; Yajna Ramphal; de Oliveira Tulio |
| EPI_ISL_4876926, EPI_ISL_4876934, EPI_ISL_4876935, EPI_ISL_4876943, EPI_ISL_4876949, EPI_ISL_4876957, EPI_ISL_4876958, EPI_ISL_4876967, EPI_ISL_4876976, EPI_ISL_4876983, EPI_ISL_4876984, EPI_ISL_4876989, EPI_ISL_4876997, EPI_ISL_4876998, EPI_ISL_4877005, EPI_ISL_4877009, EPI_ISL_4877015, EPI_ISL_4877022, EPI_ISL_4877025, EPI_ISL_4877026, EPI_ISL_4877033, EPI_ISL_4877043, EPI_ISL_4877046, EPI_ISL_4877047, EPI_ISL_4877053, EPI_ISL_4877058, EPI_ISL_5099156, EPI_ISL_5099157, EPI_ISL_5099158, EPI_ISL_5099217, EPI_ISL_5942679, EPI_ISL_5942680, EPI_ISL_5942681, EPI_ISL_5942682, EPI_ISL_5942688, EPI_ISL_5942689, EPI_ISL_5942690, EPI_ISL_5942691, EPI_ISL_5942692, EPI_ISL_5942693, EPI_ISL_5942694, EPI_ISL_5942701, EPI_ISL_5942702, EPI_ISL_5942703, EPI_ISL_5942717, EPI_ISL_5942723, EPI_ISL_5942730, EPI_ISL_5942734, EPI_ISL_5942742, EPI_ISL_5942749, EPI_ISL_5942757, EPI_ISL_5942768, EPI_ISL_5942777, EPI_ISL_5942783, EPI_ISL_5942790, EPI_ISL_5942795, EPI_ISL_5942796, EPI_ISL_5942804, EPI_ISL_5942810, EPI_ISL_5942821, EPI_ISL_5942835, EPI_ISL_5942836, EPI_ISL_5942843, EPI_ISL_6422290, EPI_ISL_6422291, EPI_ISL_6422292, EPI_ISL_6422293, EPI_ISL_6422294, EPI_ISL_6422295, EPI_ISL_6422296, EPI_ISL_6422297, EPI_ISL_6422298, EPI_ISL_6422299, EPI_ISL_6422300, EPI_ISL_6422301, EPI_ISL_6422302, EPI_ISL_6422303, EPI_ISL_6422304, EPI_ISL_6585201, EPI_ISL_6585202, EPI_ISL_6585203, EPI_ISL_6585204, EPI_ISL_6585205, EPI_ISL_6585206, EPI_ISL_6585207, EPI_ISL_6585208, EPI_ISL_6585209, EPI_ISL_6585210, EPI_ISL_6585211, EPI_ISL_6585212, EPI_ISL_6585213, EPI_ISL_6585214, EPI_ISL_6585215, EPI_ISL_6585216, EPI_ISL_6585217, EPI_ISL_6585223, EPI_ISL_6585224, EPI_ISL_6585225, EPI_ISL_6585226, EPI_ISL_6585227, EPI_ISL_6585228, EPI_ISL_6585229, EPI_ISL_6699699, EPI_ISL_6699700, EPI_ISL_6699701, EPI_ISL_6699702, EPI_ISL_6699703, EPI_ISL_6699704, EPI_ISL_6699705, EPI_ISL_6699706, EPI_ISL_6699707, EPI_ISL_6699708, EPI_ISL_6699709, EPI_ISL_6699710, EPI_ISL_6699711, EPI_ISL_6699712, EPI_ISL_6699713, EPI_ISL_6699714, EPI_ISL_6699715, EPI_ISL_6699716, EPI_ISL_6699717, EPI_ISL_6699718, EPI_ISL_6699719, EPI_ISL_6699720, EPI_ISL_6699721, EPI_ISL_6699722, EPI_ISL_6699723, EPI_ISL_6699724, EPI_ISL_6699725, EPI_ISL_6699726, EPI_ISL_6699727, EPI_ISL_6795195, EPI_ISL_6795196, EPI_ISL_6795197, EPI_ISL_6795198, EPI_ISL_6795199, EPI_ISL_6795200, EPI_ISL_6795201, EPI_ISL_6795202, EPI_ISL_6795203, EPI_ISL_6795204, EPI_ISL_6795406, EPI_ISL_7015209, EPI_ISL_7015210, EPI_ISL_7015211, EPI_ISL_7015212, EPI_ISL_7015213, EPI_ISL_7015214, EPI_ISL_7015215, EPI_ISL_7015216, EPI_ISL_7015217, EPI_ISL_7015218, EPI_ISL_7015219, EPI_ISL_7015220, EPI_ISL_7015221, EPI_ISL_7015222, EPI_ISL_7015223, EPI_ISL_7015224, EPI_ISL_7015225, EPI_ISL_7015226, EPI_ISL_7015227, EPI_ISL_7015228, EPI_ISL_7015229, EPI_ISL_7015230, EPI_ISL_7015231, EPI_ISL_7015232, EPI_ISL_7015233, EPI_ISL_7310589, EPI_ISL_7310595, EPI_ISL_7310605, EPI_ISL_7310613, EPI_ISL_7310622, EPI_ISL_7310630, EPI_ISL_7310631, EPI_ISL_7310636, EPI_ISL_7310643, EPI_ISL_7310648, EPI_ISL_7310658, EPI_ISL_7310666, EPI_ISL_7310675, EPI_ISL_7310679, EPI_ISL_7310685, EPI_ISL_7310694, EPI_ISL_7310703, EPI_ISL_7310710, EPI_ISL_7310719, EPI_ISL_7310725, EPI_ISL_7310733, EPI_ISL_7310742, EPI_ISL_7310747, EPI_ISL_7358050, EPI_ISL_7358051, EPI_ISL_7358052, EPI_ISL_7358053, EPI_ISL_7358054, EPI_ISL_7358055, EPI_ISL_7358056, EPI_ISL_7358057, EPI_ISL_7358058, EPI_ISL_7358059, EPI_ISL_7358064, EPI_ISL_7358065, EPI_ISL_7358066, EPI_ISL_7358067, EPI_ISL_7358068, EPI_ISL_7358069, EPI_ISL_7358070, EPI_ISL_7358071, EPI_ISL_7358072, EPI_ISL_7358073, EPI_ISL_7358074, EPI_ISL_7358075, EPI_ISL_7358076, EPI_ISL_7358077, EPI_ISL_7358078, EPI_ISL_7358079, EPI_ISL_7358080, EPI_ISL_7358081, EPI_ISL_7358082, EPI_ISL_7358083, EPI_ISL_7358084, EPI_ISL_7358085, EPI_ISL_7358089, EPI_ISL_7358090, EPI_ISL_7358091, EPI_ISL_7358092, EPI_ISL_7358093, EPI_ISL_7381192, EPI_ISL_7381193, EPI_ISL_7381194, EPI_ISL_7381195, EPI_ISL_7381196, EPI_ISL_7381197, EPI_ISL_7381198, EPI_ISL_7381199, EPI_ISL_7381200, EPI_ISL_7381201, EPI_ISL_7381202, EPI_ISL_7381203, EPI_ISL_7381204, EPI_ISL_7381205, EPI_ISL_7381206, EPI_ISL_7381207, EPI_ISL_7417008, EPI_ISL_7417082, EPI_ISL_7417091, EPI_ISL_7417102, EPI_ISL_7417181, EPI_ISL_7417197, EPI_ISL_7417232, EPI_ISL_7417255, EPI_ISL_7417393, EPI_ISL_7545676, EPI_ISL_7545677, EPI_ISL_7545678, EPI_ISL_7545679, EPI_ISL_7545680, EPI_ISL_7545681, EPI_ISL_7545682, EPI_ISL_7545683, EPI_ISL_7545684, EPI_ISL_7545685, EPI_ISL_7545686, EPI_ISL_7545687, EPI_ISL_7545688, EPI_ISL_7545689, EPI_ISL_7545690, EPI_ISL_7545691, EPI_ISL_7545692, EPI_ISL_7545693, EPI_ISL_7545694, EPI_ISL_7545695, EPI_ISL_7545696, EPI_ISL_7545698, EPI_ISL_7545699, EPI_ISL_7545700, EPI_ISL_7545701, EPI_ISL_7545702, EPI_ISL_7545703, EPI_ISL_7545704, EPI_ISL_7545705, EPI_ISL_7545706, EPI_ISL_7545707, EPI_ISL_7545708, EPI_ISL_7545709, EPI_ISL_7545710, EPI_ISL_7545711, EPI_ISL_7545712, EPI_ISL_7545713, EPI_ISL_7545714, EPI_ISL_7545715, EPI_ISL_7545716, EPI_ISL_7545717, EPI_ISL_7545718, EPI_ISL_7545719, EPI_ISL_7545720, EPI_ISL_7545721, EPI_ISL_7545722, EPI_ISL_7545723, EPI_ISL_7545724, EPI_ISL_7545725, EPI_ISL_7545726, EPI_ISL_7545727, EPI_ISL_7545728, EPI_ISL_7545729, EPI_ISL_7545730, EPI_ISL_7545731, EPI_ISL_7545732, EPI_ISL_7545733, EPI_ISL_7545734, EPI_ISL_7545735, EPI_ISL_7545736, EPI_ISL_7545737, EPI_ISL_7545738, EPI_ISL_7545739, EPI_ISL_7545742, EPI_ISL_7545743, EPI_ISL_7545744, EPI_ISL_7545745, EPI_ISL_7545746, EPI_ISL_7545747, EPI_ISL_7545748, EPI_ISL_7545749, EPI_ISL_7545750, EPI_ISL_7545751, EPI_ISL_7545752, EPI_ISL_7545753, EPI_ISL_7545754, EPI_ISL_7545755, EPI_ISL_7545756, EPI_ISL_7545757, EPI_ISL_7545758, EPI_ISL_7545759, EPI_ISL_7545760, EPI_ISL_7545761, EPI_ISL_7545762, EPI_ISL_7545764, EPI_ISL_7545765, EPI_ISL_7545766, EPI_ISL_7545767, EPI_ISL_7545768, EPI_ISL_7545769, EPI_ISL_7545770, EPI_ISL_7545771, EPI_ISL_7545772, EPI_ISL_7545773, EPI_ISL_7545774, EPI_ISL_7545775, EPI_ISL_7545776, EPI_ISL_7545777, EPI_ISL_7545778, EPI_ISL_7545779, EPI_ISL_7545780, EPI_ISL_7545781, EPI_ISL_7545782, EPI_ISL_7545783, EPI_ISL_7545784, EPI_ISL_7545785, EPI_ISL_7545787, EPI_ISL_7545788, EPI_ISL_7545789, EPI_ISL_7545790, EPI_ISL_7545791, EPI_ISL_7545792, EPI_ISL_7545793, EPI_ISL_7545794, EPI_ISL_7747496, EPI_ISL_7747497, EPI_ISL_7747498, EPI_ISL_7747499, EPI_ISL_7747500, EPI_ISL_7747501, EPI_ISL_7747502, EPI_ISL_7747535, EPI_ISL_7747536, EPI_ISL_7747537, EPI_ISL_7747538, EPI_ISL_7747539, EPI_ISL_7747540, EPI_ISL_7747541, EPI_ISL_7747542, EPI_ISL_7747543, EPI_ISL_7747544, EPI_ISL_7747545, EPI_ISL_7747546, EPI_ISL_7747547, EPI_ISL_7747548, EPI_ISL_7747549, EPI_ISL_7747550, EPI_ISL_7747551, EPI_ISL_7747552, EPI_ISL_7747553, EPI_ISL_7747554, EPI_ISL_7747555, EPI_ISL_7747556, EPI_ISL_7747557, EPI_ISL_7747558, EPI_ISL_7747559, EPI_ISL_7747560, EPI_ISL_7747561, EPI_ISL_7747562, EPI_ISL_7747563, EPI_ISL_7747564, EPI_ISL_7747565, EPI_ISL_7747566, EPI_ISL_7747567, EPI_ISL_7747568, EPI_ISL_7747569, EPI_ISL_7747570, EPI_ISL_7747571, EPI_ISL_7747572, EPI_ISL_7747573, EPI_ISL_7747574, EPI_ISL_7747575, EPI_ISL_7747576, EPI_ISL_7747577, EPI_ISL_7747578, EPI_ISL_7747579, EPI_ISL_7747580, EPI_ISL_7747581, EPI_ISL_7747582, EPI_ISL_7747583, EPI_ISL_7747584, EPI_ISL_7747585, EPI_ISL_7747586, EPI_ISL_7747587, EPI_ISL_7747588, EPI_ISL_7747589, EPI_ISL_7747590, EPI_ISL_7747591, EPI_ISL_7747592, EPI_ISL_7747593, EPI_ISL_7747594, EPI_ISL_7747595, EPI_ISL_7747596, EPI_ISL_7747597, EPI_ISL_7747598, EPI_ISL_7747599, EPI_ISL_7747600, EPI_ISL_7747601, EPI_ISL_7747602, EPI_ISL_7747603, EPI_ISL_7747604, EPI_ISL_7747605, EPI_ISL_7747606, EPI_ISL_7747607, EPI_ISL_7747608 | | | |
| see above | National Health Laboratory Service, Kwazulu-Natal, South Africa | CERI, Centre for Epidemic Response and Innvoation, Stellenbosch University and KRISP, KZN Research Innovation and Sequencing Platform, UKZN. | Arisha Maharaj; Emmanuel SJ; Giandhari J; Giandhari Jennifer; Moir M; Naidoo Y; Naidoo Yeshnee; Nokukhanya Mdlalose; Pillay S; Pillay Sureshnee; Ramphal U; Ramphal Y; San JE; San James; Tegally H; Tegally Houriiyah; Tshiabuila D; Tshiabuila Derek; Upasana Ramphal; Wilkinson E; Wilkinson Eduan; Yajna Ramphal; de Oliveira T; de Oliveira Tulio; van Wyk S |
| EPI_ISL_4572309, EPI_ISL_4572310, EPI_ISL_4572311, EPI_ISL_4572312, EPI_ISL_4572313, EPI_ISL_4572314, EPI_ISL_4572315, EPI_ISL_4572316, EPI_ISL_4572317, EPI_ISL_4572318, EPI_ISL_4572326, EPI_ISL_4572332, EPI_ISL_4572354, EPI_ISL_4572362, EPI_ISL_4572377, EPI_ISL_4572380 | | | |
| see above | National Health Laboratory Service, Kwazulu-Natal, South Africa | KRISP, KZN Research Innovation and Sequencing Platform | Giandhari Jennifer; Naidoo Yeshnee; Nokukhanya Mdlalose; Pillay Sureshnee; San James; Tegally Houriiyah; Tshiabuila Derek; Wilkinson Eduan; Yajna Ramphal; de Oliveira Tulio |
| EPI_ISL_6829556, EPI_ISL_6829557, EPI_ISL_6829558 | National Health Laboratory Service, Kwazulu-Natal, South Africa | KRISP, KZN Research Innovation and Sequencing Platform | Arisha Maharaj; Giandhari J; Lessells R; Moir M; Naidoo Y; Nokukhanya M; Pillay S; Ramphal U; Ramphal Y; San JE; Tegally H; Tshiabuila D; Wilkinson E; de Oliveira T |
| EPI_ISL_5327838, EPI_ISL_5327839, EPI_ISL_5327840, EPI_ISL_5327841, EPI_ISL_5327842, EPI_ISL_5327843, EPI_ISL_5327844, EPI_ISL_5327845, EPI_ISL_5327846, EPI_ISL_5327847, EPI_ISL_5327848, EPI_ISL_5327849, EPI_ISL_5327850, EPI_ISL_5327851, EPI_ISL_5327852, EPI_ISL_5327853, EPI_ISL_5327854, EPI_ISL_5327855, EPI_ISL_5327856, EPI_ISL_5327857, EPI_ISL_5327858, EPI_ISL_5327859, EPI_ISL_5327860, EPI_ISL_5327861, EPI_ISL_5327862, EPI_ISL_5327863 | | | |
| see above | National Health Laboratory Service, South Africa | CERI, Centre for Epidemic Response and Innvoation, Stellenbosch University and KRISP, KZN Research Innovation and Sequencing Platform, UKZN. | Emmanuel S; Giandhari J; Naidoo Yeshnee; Nokukhanya Mdlalose; Pillay S; Tegally H; Tshabuila Derek; Wilkinson E; Yajna Ramphal; de Oliveira T |
| EPI_ISL_1250424, EPI_ISL_1250425, EPI_ISL_1250427, EPI_ISL_1250428, EPI_ISL_1250430, EPI_ISL_1250431, EPI_ISL_1250432, EPI_ISL_1250434, EPI_ISL_1250436, EPI_ISL_1250437, EPI_ISL_1250439, EPI_ISL_1250441, EPI_ISL_1250442, EPI_ISL_1250444, EPI_ISL_1250446, EPI_ISL_1250447, EPI_ISL_1250448, EPI_ISL_1250450, EPI_ISL_1250452, EPI_ISL_1250453, EPI_ISL_1250455, EPI_ISL_1250456, EPI_ISL_1250458, EPI_ISL_1250460, EPI_ISL_1250462, EPI_ISL_1250463, EPI_ISL_1250465, EPI_ISL_1250466, EPI_ISL_1250468, EPI_ISL_1250469, EPI_ISL_1250472, EPI_ISL_1250474, EPI_ISL_1250476, EPI_ISL_1250477, EPI_ISL_1250479, EPI_ISL_1250480, EPI_ISL_1250482, EPI_ISL_1250483, EPI_ISL_1250485, EPI_ISL_1250486, EPI_ISL_1250488, EPI_ISL_1250490, EPI_ISL_1250491, EPI_ISL_1250493, EPI_ISL_1250494, EPI_ISL_1250496, EPI_ISL_1250497, EPI_ISL_1250499, EPI_ISL_1250501, EPI_ISL_1250502, EPI_ISL_1250504, EPI_ISL_1250505, EPI_ISL_1550912, EPI_ISL_1550913, EPI_ISL_1550914, EPI_ISL_1550915, EPI_ISL_1550916, EPI_ISL_1550917, EPI_ISL_1550918, EPI_ISL_1550919, EPI_ISL_1550920, EPI_ISL_1550921, EPI_ISL_1550922, EPI_ISL_1550923, EPI_ISL_1550924, EPI_ISL_1550925, EPI_ISL_1550926, EPI_ISL_1550927, EPI_ISL_1550928, EPI_ISL_1550929, EPI_ISL_1550930, EPI_ISL_1550931, EPI_ISL_1550932, EPI_ISL_1550933, EPI_ISL_1550934, EPI_ISL_1550935, EPI_ISL_1550936, EPI_ISL_1550937, EPI_ISL_1550938, EPI_ISL_1550939, EPI_ISL_1550940, EPI_ISL_1550941, EPI_ISL_1550942, EPI_ISL_1550943, EPI_ISL_1550944, EPI_ISL_1550945, EPI_ISL_1550946, EPI_ISL_1550947, EPI_ISL_1550948, EPI_ISL_1550949, EPI_ISL_1550950, EPI_ISL_1550951, EPI_ISL_1550952, EPI_ISL_1550953, EPI_ISL_1550954, EPI_ISL_1550955, EPI_ISL_1550956, EPI_ISL_1550957, EPI_ISL_1550958, EPI_ISL_1550959, EPI_ISL_1550960, EPI_ISL_1550961, EPI_ISL_1550962, EPI_ISL_1817755, EPI_ISL_1817756, EPI_ISL_1817757, EPI_ISL_1817758, EPI_ISL_1817759, EPI_ISL_1817760, EPI_ISL_1817761, EPI_ISL_1817762, EPI_ISL_1817763, EPI_ISL_1817764, EPI_ISL_1817765, EPI_ISL_1817766, EPI_ISL_1817767, EPI_ISL_1817768, EPI_ISL_1817769, EPI_ISL_1817770, EPI_ISL_1817771, EPI_ISL_1817772, EPI_ISL_1817773, EPI_ISL_1817774, EPI_ISL_1817775, EPI_ISL_1817776, EPI_ISL_1817777, EPI_ISL_1817778, EPI_ISL_1817779, EPI_ISL_1817780, EPI_ISL_1817781, EPI_ISL_1817782, EPI_ISL_1817783, EPI_ISL_1817784, EPI_ISL_1817785, EPI_ISL_1817786, EPI_ISL_1817787, EPI_ISL_1817788, EPI_ISL_1817789, EPI_ISL_1817790, EPI_ISL_1817791, EPI_ISL_1817792, EPI_ISL_1817793, EPI_ISL_1817794, EPI_ISL_1817795, EPI_ISL_1817796, EPI_ISL_1817797, EPI_ISL_1817798, EPI_ISL_1817799, EPI_ISL_1817800, EPI_ISL_1817801, EPI_ISL_1817802, EPI_ISL_1817803, EPI_ISL_1817804, EPI_ISL_1817805, EPI_ISL_1817806, EPI_ISL_1817807, EPI_ISL_1817808, EPI_ISL_1817809, EPI_ISL_1817810, EPI_ISL_1817811, EPI_ISL_1817812, EPI_ISL_1817813, EPI_ISL_1817814, EPI_ISL_1817815, EPI_ISL_1817816, EPI_ISL_1817817, EPI_ISL_1817818, EPI_ISL_1817819, EPI_ISL_1817820, EPI_ISL_1817821, EPI_ISL_1817822, EPI_ISL_1817823, EPI_ISL_1817824, EPI_ISL_1817825, EPI_ISL_1817826, EPI_ISL_1817827, EPI_ISL_1817828, EPI_ISL_1817829, EPI_ISL_1817830, EPI_ISL_1817831, EPI_ISL_1817832, EPI_ISL_1817833, EPI_ISL_1817834, EPI_ISL_1817835, EPI_ISL_1817836, EPI_ISL_1817837, EPI_ISL_1817838, EPI_ISL_1817839, EPI_ISL_1817840, EPI_ISL_1817841, EPI_ISL_1817842, EPI_ISL_1817843, EPI_ISL_1817844, EPI_ISL_1817845, EPI_ISL_1817846, EPI_ISL_1817848, EPI_ISL_1817849, EPI_ISL_1817850, EPI_ISL_1817851, EPI_ISL_1817852, EPI_ISL_1817853, EPI_ISL_1817854, EPI_ISL_1817855, EPI_ISL_1817856, EPI_ISL_1817857, EPI_ISL_1817858, EPI_ISL_1817859, EPI_ISL_1817860, EPI_ISL_1817861, EPI_ISL_1817862, EPI_ISL_1817863, EPI_ISL_2086228, EPI_ISL_2086229, EPI_ISL_2086230, EPI_ISL_2086231, EPI_ISL_2086232, EPI_ISL_2086233, EPI_ISL_2086234, EPI_ISL_2086235, EPI_ISL_2086236, EPI_ISL_2086237, EPI_ISL_2086238, EPI_ISL_2086239, EPI_ISL_2086240, EPI_ISL_2086241, EPI_ISL_2086242, EPI_ISL_2086243, EPI_ISL_2086244, EPI_ISL_2086245, EPI_ISL_2086246, EPI_ISL_2086247, EPI_ISL_2397000, EPI_ISL_2397001, EPI_ISL_2397002, EPI_ISL_2397003, EPI_ISL_2397004, EPI_ISL_2397005, EPI_ISL_2397006, EPI_ISL_2397007, EPI_ISL_2397008, EPI_ISL_2397009, EPI_ISL_2397010, EPI_ISL_2397011, EPI_ISL_2397012, EPI_ISL_2397013, EPI_ISL_2397014, EPI_ISL_2397015, EPI_ISL_2397016, EPI_ISL_2397017, EPI_ISL_2397018, EPI_ISL_2397019, EPI_ISL_2397020, EPI_ISL_2397021, EPI_ISL_2397022, EPI_ISL_2397023, EPI_ISL_2397024, EPI_ISL_2397025, EPI_ISL_2397026, EPI_ISL_2397027, EPI_ISL_2397028, EPI_ISL_2397029, EPI_ISL_2397030, EPI_ISL_2397031, EPI_ISL_2397032, EPI_ISL_2397033, EPI_ISL_2397034, EPI_ISL_2397035, EPI_ISL_2397036, EPI_ISL_2397037, EPI_ISL_2397038, EPI_ISL_2397039, EPI_ISL_2397040, EPI_ISL_2397041, EPI_ISL_2397042, EPI_ISL_2397043, EPI_ISL_2397044, EPI_ISL_2397045, EPI_ISL_2397046, EPI_ISL_2397047, EPI_ISL_2397048, EPI_ISL_2397049, EPI_ISL_2397050, EPI_ISL_2397051, EPI_ISL_2397052, EPI_ISL_2397053, EPI_ISL_2397054, EPI_ISL_2397055, EPI_ISL_2397056, EPI_ISL_2397057, EPI_ISL_2397058, EPI_ISL_2397059, EPI_ISL_2397060, EPI_ISL_2397061, EPI_ISL_2397062, EPI_ISL_2397063, EPI_ISL_2397064, EPI_ISL_2397065, EPI_ISL_2397066, EPI_ISL_2397067, EPI_ISL_2397068, EPI_ISL_2397069, EPI_ISL_2397070, EPI_ISL_2397071, EPI_ISL_2397072, EPI_ISL_2397073, EPI_ISL_2397074, EPI_ISL_2397075, EPI_ISL_2397076, EPI_ISL_2397077, EPI_ISL_2397078, EPI_ISL_2397079, EPI_ISL_2397080, EPI_ISL_2397081, EPI_ISL_2397082, EPI_ISL_2397083, EPI_ISL_2397084, EPI_ISL_2397085, EPI_ISL_2397086, EPI_ISL_2397087, EPI_ISL_2397088, EPI_ISL_2397089, EPI_ISL_2397090, EPI_ISL_2397091, EPI_ISL_2397092, EPI_ISL_2397093, EPI_ISL_2397094, EPI_ISL_2397095, EPI_ISL_2397096, EPI_ISL_2397097, EPI_ISL_2397098, EPI_ISL_2397099, EPI_ISL_2397100, EPI_ISL_2397101, EPI_ISL_2397102, EPI_ISL_2397103, EPI_ISL_2397104, EPI_ISL_2397105, EPI_ISL_2397106, EPI_ISL_2397107, EPI_ISL_2397108, EPI_ISL_2397109, EPI_ISL_2397110, EPI_ISL_2397111, EPI_ISL_2397112, EPI_ISL_2397113, EPI_ISL_2397114, EPI_ISL_2397115, EPI_ISL_2397116, EPI_ISL_2397117, EPI_ISL_2397118, EPI_ISL_2397119, EPI_ISL_2397120, EPI_ISL_2397121, EPI_ISL_2397122, EPI_ISL_2397123, EPI_ISL_2397124, EPI_ISL_2397125, EPI_ISL_2397126, EPI_ISL_2397127, EPI_ISL_2397128, EPI_ISL_2397129, EPI_ISL_2397130, EPI_ISL_2397131, EPI_ISL_2397132, EPI_ISL_2397133, EPI_ISL_2397134, EPI_ISL_2397135, EPI_ISL_2397136, EPI_ISL_2397137, EPI_ISL_2397138, EPI_ISL_2397139, EPI_ISL_2397140, EPI_ISL_2397141, EPI_ISL_2397142, EPI_ISL_2397143, EPI_ISL_2397144, EPI_ISL_2397145, EPI_ISL_2397146, EPI_ISL_2397147, EPI_ISL_2397148, EPI_ISL_2397149, EPI_ISL_2397150, EPI_ISL_2397151, EPI_ISL_2397152, EPI_ISL_2397153, EPI_ISL_2397154, EPI_ISL_2397155, EPI_ISL_2397156, EPI_ISL_2397157, EPI_ISL_2397158, EPI_ISL_2494729, EPI_ISL_2494753, EPI_ISL_2494755, EPI_ISL_2494770, EPI_ISL_2494771, EPI_ISL_2494772, EPI_ISL_2494773, EPI_ISL_2494775, EPI_ISL_2494778, EPI_ISL_2494779, EPI_ISL_2494780, EPI_ISL_2494781, EPI_ISL_2494784, EPI_ISL_2494785, EPI_ISL_2494786, EPI_ISL_2494787, EPI_ISL_2494789, EPI_ISL_2494790, EPI_ISL_2494791, EPI_ISL_2494792, EPI_ISL_2494793, EPI_ISL_2494795, EPI_ISL_2494796, EPI_ISL_2494798, EPI_ISL_2494799, EPI_ISL_2494800, EPI_ISL_2494801, EPI_ISL_2494802, EPI_ISL_2494803, EPI_ISL_2494804, EPI_ISL_2494805, EPI_ISL_2494806, EPI_ISL_2494807, EPI_ISL_2494808, EPI_ISL_2494809, EPI_ISL_2494810, EPI_ISL_2494811, EPI_ISL_2494812, EPI_ISL_2494813, EPI_ISL_2494814, EPI_ISL_2494815, EPI_ISL_2494816, EPI_ISL_2494817, EPI_ISL_2494818, EPI_ISL_2494819, EPI_ISL_2494820, EPI_ISL_2494821, EPI_ISL_2494823, EPI_ISL_2494824, EPI_ISL_2494825, EPI_ISL_2494826, EPI_ISL_2494827, EPI_ISL_2494828, EPI_ISL_2494829, EPI_ISL_2494832, EPI_ISL_2494833, EPI_ISL_2494834, EPI_ISL_2494835, EPI_ISL_2494836, EPI_ISL_2494837, EPI_ISL_2494838, EPI_ISL_2494839, EPI_ISL_2494840, EPI_ISL_2494841, EPI_ISL_2494842, EPI_ISL_2494843, EPI_ISL_2494845, EPI_ISL_2494846, EPI_ISL_2494847, EPI_ISL_2494848, EPI_ISL_2494849, EPI_ISL_2494850, EPI_ISL_2494851, EPI_ISL_2494852, EPI_ISL_2494853, EPI_ISL_2494854, EPI_ISL_2494855, EPI_ISL_2494856, EPI_ISL_2494857, EPI_ISL_2494858, EPI_ISL_2494860, EPI_ISL_2494861, EPI_ISL_2494863, EPI_ISL_2494865, EPI_ISL_2494866, EPI_ISL_2494869, EPI_ISL_2494873, EPI_ISL_2494875, EPI_ISL_2494876, EPI_ISL_2494877, EPI_ISL_2494883, EPI_ISL_2494886, EPI_ISL_2494887, EPI_ISL_2494888, EPI_ISL_2494890, EPI_ISL_2494892, EPI_ISL_2494893, EPI_ISL_2494894, EPI_ISL_2494895, EPI_ISL_2494896, EPI_ISL_2494897, EPI_ISL_2494904, EPI_ISL_2494905, EPI_ISL_2494908, EPI_ISL_2494909, EPI_ISL_2494910, EPI_ISL_2494911, EPI_ISL_2494923, EPI_ISL_2494927, EPI_ISL_2494930, EPI_ISL_2494935, EPI_ISL_2494936, EPI_ISL_2494939, EPI_ISL_2494940, EPI_ISL_2494945, EPI_ISL_2494948, EPI_ISL_2494959, EPI_ISL_2494965, EPI_ISL_2494978, EPI_ISL_2494998, EPI_ISL_2495028, EPI_ISL_2495032, EPI_ISL_2495046, EPI_ISL_2495047, EPI_ISL_2495049, EPI_ISL_2495050, EPI_ISL_2495051, EPI_ISL_2495052, EPI_ISL_2495053, EPI_ISL_3663498, EPI_ISL_3663499, EPI_ISL_3663500, EPI_ISL_3663501, EPI_ISL_3663502, EPI_ISL_3663503, EPI_ISL_3663504, EPI_ISL_3663505, EPI_ISL_3663506, EPI_ISL_3663507, EPI_ISL_3663508, EPI_ISL_3663509, EPI_ISL_3663510, EPI_ISL_3663511, EPI_ISL_3663512, EPI_ISL_3663513, EPI_ISL_3663514, EPI_ISL_3663515, EPI_ISL_3663516, EPI_ISL_3663517, EPI_ISL_3663518, EPI_ISL_3663519, EPI_ISL_3663520, EPI_ISL_3663521, EPI_ISL_3663522, EPI_ISL_3663523, EPI_ISL_3663524, EPI_ISL_3663525, EPI_ISL_3663526, EPI_ISL_3663527, EPI_ISL_3663528, EPI_ISL_3663529, EPI_ISL_3663530, EPI_ISL_3663531, EPI_ISL_3663532, EPI_ISL_3663533, EPI_ISL_3663534, EPI_ISL_3663535, EPI_ISL_3663536, EPI_ISL_3663537, EPI_ISL_3663538, EPI_ISL_3663539, EPI_ISL_3663540, EPI_ISL_3663541, EPI_ISL_3663542, EPI_ISL_3663543, EPI_ISL_3663544 | | | |
| see above | National Health Laboratory Service, South Africa | KRISP, KZN Research Innovation and Sequencing Platform | Emmanuel SJ; Giandhari J; Giandhari Jennifer; Khan S; Lessells R; Maslo C; Mdlalose K; Naidoo Y; Naidoo Yeshnee; Pillay S; Pillay Sureshnee; Ramphal U; San James; Sitharam L; Tegally H; Tegally Houriiyah; Tshabuila Derek; Wilkinson E; Wilkinson Eduan; Yajna Ramphal; York D; de Oliveira T; de Oliveira Tulio |
| EPI_ISL_1048462, EPI_ISL_1048463, EPI_ISL_1048464, EPI_ISL_1048465, EPI_ISL_1048466, EPI_ISL_1048467, EPI_ISL_1048468, EPI_ISL_1048469, EPI_ISL_1048470, EPI_ISL_1048471, EPI_ISL_1048472, EPI_ISL_1048473, EPI_ISL_1048474, EPI_ISL_1048475, EPI_ISL_1048476, EPI_ISL_1048477, EPI_ISL_1048478, EPI_ISL_1048479, EPI_ISL_1048480, EPI_ISL_1048481, EPI_ISL_1048482, EPI_ISL_1048483, | | | |

| | | | |
|---|---|---|---|
| EPI_ISL_1048484, EPI_ISL_1048485, EPI_ISL_1048486, EPI_ISL_1048487, EPI_ISL_1048488, EPI_ISL_1048489, EPI_ISL_1048490, EPI_ISL_1048491, EPI_ISL_1048492, EPI_ISL_1048493, EPI_ISL_1048494, EPI_ISL_1048495, EPI_ISL_1048496, EPI_ISL_1048497, EPI_ISL_1048498, EPI_ISL_1048499, EPI_ISL_1048500, EPI_ISL_1048501, EPI_ISL_1048502, EPI_ISL_1048503, EPI_ISL_1048504, EPI_ISL_1048505, EPI_ISL_1048506, EPI_ISL_1048507, EPI_ISL_1048508, EPI_ISL_1048509, EPI_ISL_1048510, EPI_ISL_1048511, EPI_ISL_1048512, EPI_ISL_1048513, EPI_ISL_1048514, EPI_ISL_1048515, EPI_ISL_1048516, EPI_ISL_1048517, EPI_ISL_1048518, EPI_ISL_1048519, EPI_ISL_1048520, EPI_ISL_1048521, EPI_ISL_1048522, EPI_ISL_1048523, EPI_ISL_1048524, EPI_ISL_1048525, EPI_ISL_1048526, EPI_ISL_1048527, EPI_ISL_1048528, EPI_ISL_1048529, EPI_ISL_1048530, EPI_ISL_1048531, EPI_ISL_1048532, EPI_ISL_1048533, EPI_ISL_1048534, EPI_ISL_1048535, EPI_ISL_1048536, EPI_ISL_1048537, EPI_ISL_1048538, EPI_ISL_1048539, EPI_ISL_1048540, EPI_ISL_1048541, EPI_ISL_1048542, EPI_ISL_1048543, EPI_ISL_1048544, EPI_ISL_1048545, EPI_ISL_1048546, EPI_ISL_1048547, EPI_ISL_1048548, EPI_ISL_1048549, EPI_ISL_1048550, EPI_ISL_1048551, EPI_ISL_1048552, EPI_ISL_1048553, EPI_ISL_1048554, EPI_ISL_1048555, EPI_ISL_1048556, EPI_ISL_1048557, EPI_ISL_1048558, EPI_ISL_1048559, EPI_ISL_1048560, EPI_ISL_1048561, EPI_ISL_1048562, EPI_ISL_1048563, EPI_ISL_1048564, EPI_ISL_1048565, EPI_ISL_1048566, EPI_ISL_1048567, EPI_ISL_1048568, EPI_ISL_1371906, EPI_ISL_1371907, EPI_ISL_1371908, EPI_ISL_1371909, EPI_ISL_1371910, EPI_ISL_1371911, EPI_ISL_1371912, EPI_ISL_1371913, EPI_ISL_1371914, EPI_ISL_1371915, EPI_ISL_1371916, EPI_ISL_1371917, EPI_ISL_1371918, EPI_ISL_1371919, EPI_ISL_1371920, EPI_ISL_1371921, EPI_ISL_1371922, EPI_ISL_1371923, EPI_ISL_1371924, EPI_ISL_1371925, EPI_ISL_1371926, EPI_ISL_1371927, EPI_ISL_1371928, EPI_ISL_1371929, EPI_ISL_1371930, EPI_ISL_1371931, EPI_ISL_1371932, EPI_ISL_1371933, EPI_ISL_1371934, EPI_ISL_1371935, EPI_ISL_1371936, EPI_ISL_1371937, EPI_ISL_1371938, EPI_ISL_1371939, EPI_ISL_1371940, EPI_ISL_1371941, EPI_ISL_1371942, EPI_ISL_1371943, EPI_ISL_1371944, EPI_ISL_1371945, EPI_ISL_1371946, EPI_ISL_1371947, EPI_ISL_1371948, EPI_ISL_1371949, EPI_ISL_1371950, EPI_ISL_1371951, EPI_ISL_1371952, EPI_ISL_1371953, EPI_ISL_1371954, EPI_ISL_1371955, EPI_ISL_1371956, EPI_ISL_1371957, EPI_ISL_1371958, EPI_ISL_1371959, EPI_ISL_1371960, EPI_ISL_1371961, EPI_ISL_1371962, EPI_ISL_1371963, EPI_ISL_1371964, EPI_ISL_1371965, EPI_ISL_1371966, EPI_ISL_1371967, EPI_ISL_1371968, EPI_ISL_1371969, EPI_ISL_1371970, EPI_ISL_1371971, EPI_ISL_1371972, EPI_ISL_1371973, EPI_ISL_1371974, EPI_ISL_1371975, EPI_ISL_1371976, EPI_ISL_1371977, EPI_ISL_1371978, EPI_ISL_1371979, EPI_ISL_1371980, EPI_ISL_1371981, EPI_ISL_1371982, EPI_ISL_1371983, EPI_ISL_1371984, EPI_ISL_1371985, EPI_ISL_1371986, EPI_ISL_1371987, EPI_ISL_1371988, EPI_ISL_1371989, EPI_ISL_1371990, EPI_ISL_1371991, EPI_ISL_1371992, EPI_ISL_1371993, EPI_ISL_1371994, EPI_ISL_1371995, EPI_ISL_1371996, EPI_ISL_1371997, EPI_ISL_1371998, EPI_ISL_1371999, EPI_ISL_1372000, EPI_ISL_1372001, EPI_ISL_1372002, EPI_ISL_1372003, EPI_ISL_1372004, EPI_ISL_1372005, EPI_ISL_1372006, EPI_ISL_2617186, EPI_ISL_2617187, EPI_ISL_2617188, EPI_ISL_2617189, EPI_ISL_2617190, EPI_ISL_2617191, EPI_ISL_2617192, EPI_ISL_2617193, EPI_ISL_2617194, EPI_ISL_2617195, EPI_ISL_2617196, EPI_ISL_2617197, EPI_ISL_2617198, EPI_ISL_2617199, EPI_ISL_2617200, EPI_ISL_2617201, EPI_ISL_2617202, EPI_ISL_2617203, EPI_ISL_2617204, EPI_ISL_2617205, EPI_ISL_2617206, EPI_ISL_2617207, EPI_ISL_2617208, EPI_ISL_2617209, EPI_ISL_2617210, EPI_ISL_2617211, EPI_ISL_2617212, EPI_ISL_2617213, EPI_ISL_2617214, EPI_ISL_2617215, EPI_ISL_2617216, EPI_ISL_2688531, EPI_ISL_2688532, EPI_ISL_2688533, EPI_ISL_2688534, EPI_ISL_2688535, EPI_ISL_2688536, EPI_ISL_2688537, EPI_ISL_2688538, EPI_ISL_2688539, EPI_ISL_2688540, EPI_ISL_2688541, EPI_ISL_2688542, EPI_ISL_2688543, EPI_ISL_2688544, EPI_ISL_2688545, EPI_ISL_2688546, EPI_ISL_2688549, EPI_ISL_2688550, EPI_ISL_2688551, EPI_ISL_2688552, EPI_ISL_2688553, EPI_ISL_2688554, EPI_ISL_2688555, EPI_ISL_2688556, EPI_ISL_2688557, EPI_ISL_2688558, EPI_ISL_2688559, EPI_ISL_2688560, EPI_ISL_2688561, EPI_ISL_2688562, EPI_ISL_2688563, EPI_ISL_2688564, EPI_ISL_2688565, EPI_ISL_2688566, EPI_ISL_2688567, EPI_ISL_2688568, EPI_ISL_2688569, EPI_ISL_2688570, EPI_ISL_2688571, EPI_ISL_2688572, EPI_ISL_2688573, EPI_ISL_2688574, EPI_ISL_2688575, EPI_ISL_2899687, EPI_ISL_2899688, EPI_ISL_2899689, EPI_ISL_2899690, EPI_ISL_2899691, EPI_ISL_2899692, EPI_ISL_2899693, EPI_ISL_2899694, EPI_ISL_2899695, EPI_ISL_2899696, EPI_ISL_2899697, EPI_ISL_2899698, EPI_ISL_2899699, EPI_ISL_2899700, EPI_ISL_2899701, EPI_ISL_2899702, EPI_ISL_2899703, EPI_ISL_2899704, EPI_ISL_2899705, EPI_ISL_2899706, EPI_ISL_2899707, EPI_ISL_2899708, EPI_ISL_2899709, EPI_ISL_2899710, EPI_ISL_2899711, EPI_ISL_2899712, EPI_ISL_2899713, EPI_ISL_2899714, EPI_ISL_2899715, EPI_ISL_2899716, EPI_ISL_2899717, EPI_ISL_2899718, EPI_ISL_2899719, EPI_ISL_2899720, EPI_ISL_2899721, EPI_ISL_2899722, EPI_ISL_2899723, EPI_ISL_2899724, EPI_ISL_2899725, EPI_ISL_2899726, EPI_ISL_2899727, EPI_ISL_2899728, EPI_ISL_2899729, EPI_ISL_2899730, EPI_ISL_2899731, EPI_ISL_2899732, EPI_ISL_2899733, EPI_ISL_2899734, EPI_ISL_2899735, EPI_ISL_2899736, EPI_ISL_2899737, EPI_ISL_2899738, EPI_ISL_2899739, EPI_ISL_2899740, EPI_ISL_3132574, EPI_ISL_3132575, EPI_ISL_3132576, EPI_ISL_3132577, EPI_ISL_3132578, EPI_ISL_3132579, EPI_ISL_3132580, EPI_ISL_3132581, EPI_ISL_3132582, EPI_ISL_3132583, EPI_ISL_3132584, EPI_ISL_3132585, EPI_ISL_3132586, EPI_ISL_3132587, EPI_ISL_3132588, EPI_ISL_3132589, EPI_ISL_3132590, EPI_ISL_3132591, EPI_ISL_3132592, EPI_ISL_3132593, EPI_ISL_3132594, EPI_ISL_3132595, EPI_ISL_3132596, EPI_ISL_3132597, EPI_ISL_3132598, EPI_ISL_3132599, EPI_ISL_3132600, EPI_ISL_3132601, EPI_ISL_3132602, EPI_ISL_3132603, EPI_ISL_3132604, EPI_ISL_3132605, EPI_ISL_3132606, EPI_ISL_3132607, EPI_ISL_3132608, EPI_ISL_3132609, EPI_ISL_3132610, EPI_ISL_3132611, EPI_ISL_3132612, EPI_ISL_3132613, EPI_ISL_3132614, EPI_ISL_3132615, EPI_ISL_3132616, EPI_ISL_3132617, EPI_ISL_3132618, EPI_ISL_3132619, EPI_ISL_3132620, EPI_ISL_3132621, EPI_ISL_3132622, EPI_ISL_3730295, EPI_ISL_3730296, EPI_ISL_3730298, EPI_ISL_3730300, EPI_ISL_3730301, EPI_ISL_3730302, EPI_ISL_3730303, EPI_ISL_3730304, EPI_ISL_3730305, EPI_ISL_3730306, EPI_ISL_3730307, EPI_ISL_3730308, EPI_ISL_3730309, EPI_ISL_3730310, EPI_ISL_3730311, EPI_ISL_3730312, EPI_ISL_3730313, EPI_ISL_3730315, EPI_ISL_3730316, EPI_ISL_3730318, EPI_ISL_3730320, EPI_ISL_3730322, EPI_ISL_3730323, EPI_ISL_3730324, EPI_ISL_3730325, EPI_ISL_3730326, EPI_ISL_3730327, EPI_ISL_3730328, EPI_ISL_3730329, EPI_ISL_3730331, EPI_ISL_3730333, EPI_ISL_3730334, EPI_ISL_3730336, EPI_ISL_3730337, EPI_ISL_3730339, EPI_ISL_3730340, EPI_ISL_3730341, EPI_ISL_3730342, EPI_ISL_3730343, EPI_ISL_3730347, EPI_ISL_3730348, EPI_ISL_3730349, EPI_ISL_3730350, EPI_ISL_3730351, EPI_ISL_3730352, EPI_ISL_3730354, EPI_ISL_3730356, EPI_ISL_3730357, EPI_ISL_3730358, EPI_ISL_3730361, EPI_ISL_3730362, EPI_ISL_3730363, EPI_ISL_3730364, EPI_ISL_3730365, EPI_ISL_3730367, EPI_ISL_3730368, EPI_ISL_3730370, EPI_ISL_3730371, EPI_ISL_3799011, EPI_ISL_3799012, EPI_ISL_3799015, EPI_ISL_3799020, EPI_ISL_3799021, EPI_ISL_3799023, EPI_ISL_3799024, EPI_ISL_3799025, EPI_ISL_3799026, EPI_ISL_3799029, EPI_ISL_3799030, EPI_ISL_3799031, EPI_ISL_3799032, EPI_ISL_3799033, EPI_ISL_3799034, EPI_ISL_3799037, EPI_ISL_3799038, EPI_ISL_3799039, EPI_ISL_3799041, EPI_ISL_3799042, EPI_ISL_3799043, EPI_ISL_3799044, EPI_ISL_3799045, EPI_ISL_3799046, EPI_ISL_3799047, EPI_ISL_3799048, EPI_ISL_3799049, EPI_ISL_3799050, EPI_ISL_3799051, EPI_ISL_3799052, EPI_ISL_3799053, EPI_ISL_3799054, EPI_ISL_3799055, EPI_ISL_3799057, EPI_ISL_3799058, EPI_ISL_3799059, EPI_ISL_3799060, EPI_ISL_3799061, EPI_ISL_3799065, EPI_ISL_3799067, EPI_ISL_3799068, EPI_ISL_3799070, EPI_ISL_3799073, EPI_ISL_3799075, EPI_ISL_3799077, EPI_ISL_3799079, EPI_ISL_3799081, EPI_ISL_3799087, EPI_ISL_3799091, EPI_ISL_3799093, EPI_ISL_3799097, EPI_ISL_3799099, EPI_ISL_3799101, EPI_ISL_3799102, EPI_ISL_3799106, EPI_ISL_3799108, EPI_ISL_3799111, EPI_ISL_3799112, EPI_ISL_3799114, EPI_ISL_3799118, EPI_ISL_3799120, EPI_ISL_3799126, EPI_ISL_3799130, EPI_ISL_3799132, EPI_ISL_3799133, EPI_ISL_3799135, EPI_ISL_3799141, EPI_ISL_3799147, EPI_ISL_3799149, EPI_ISL_3799150, EPI_ISL_3799152, EPI_ISL_3799154, EPI_ISL_3799157, EPI_ISL_3799163, EPI_ISL_3799164, EPI_ISL_3799167, EPI_ISL_3799172, EPI_ISL_3799173, EPI_ISL_3799174, EPI_ISL_3799175, EPI_ISL_3799176, EPI_ISL_3799177, EPI_ISL_3799178, EPI_ISL_3799179, EPI_ISL_3799180, EPI_ISL_3799181, EPI_ISL_3799182, EPI_ISL_3799183, EPI_ISL_3799184, EPI_ISL_3799185, EPI_ISL_3939037, EPI_ISL_3939038, EPI_ISL_3939039, EPI_ISL_3939040, EPI_ISL_3939041, EPI_ISL_3939042, EPI_ISL_3939043, EPI_ISL_3939044, EPI_ISL_3939045, EPI_ISL_3939046, EPI_ISL_3939047, EPI_ISL_3939048, EPI_ISL_3939049, EPI_ISL_3939050, EPI_ISL_3939051, EPI_ISL_3939052, EPI_ISL_3939053, EPI_ISL_3939054, EPI_ISL_3939055, EPI_ISL_3939056, EPI_ISL_3939057, EPI_ISL_3939058, EPI_ISL_3939059, EPI_ISL_3939060, EPI_ISL_3939061, EPI_ISL_3939062, EPI_ISL_3939063, EPI_ISL_3939065, EPI_ISL_3939066, EPI_ISL_3939067, EPI_ISL_3939069, EPI_ISL_3939070, EPI_ISL_3939071, EPI_ISL_3939072, EPI_ISL_3939073, EPI_ISL_3939074, EPI_ISL_3939075, EPI_ISL_3939076, EPI_ISL_3939077, EPI_ISL_3939078, EPI_ISL_3939079, EPI_ISL_3939080, EPI_ISL_3939081, EPI_ISL_3939082, EPI_ISL_3939083, EPI_ISL_3939084, EPI_ISL_3939085, EPI_ISL_3939086, EPI_ISL_3939087, EPI_ISL_3939089, EPI_ISL_3939090, EPI_ISL_3939091, EPI_ISL_3939092, EPI_ISL_3939093, EPI_ISL_3939094, EPI_ISL_3939095, EPI_ISL_3939096, EPI_ISL_3939097, EPI_ISL_3939098, EPI_ISL_3939099, EPI_ISL_3939100, EPI_ISL_3939101, EPI_ISL_4121561, EPI_ISL_4121575, EPI_ISL_4121585, EPI_ISL_4121588, EPI_ISL_4121591, EPI_ISL_4121595, EPI_ISL_4121598, EPI_ISL_4121601, EPI_ISL_4121603, EPI_ISL_4121615, EPI_ISL_4121624, EPI_ISL_4121628, EPI_ISL_4121631, EPI_ISL_4121646, EPI_ISL_4121649, EPI_ISL_4121653, EPI_ISL_4121690, EPI_ISL_4121694, EPI_ISL_4121697, EPI_ISL_4121700, EPI_ISL_4121704, EPI_ISL_4121707, EPI_ISL_4121716, EPI_ISL_4121719, EPI_ISL_4121722, EPI_ISL_4121725, EPI_ISL_4121737, EPI_ISL_4121742, EPI_ISL_4121746, EPI_ISL_4121749, EPI_ISL_4121758, EPI_ISL_4121761, EPI_ISL_4121767, EPI_ISL_4121773, EPI_ISL_4121776, EPI_ISL_4121782, EPI_ISL_4121789, EPI_ISL_4121793, EPI_ISL_4336894, EPI_ISL_4336897, EPI_ISL_4336900, EPI_ISL_4336902, EPI_ISL_4336905, EPI_ISL_4336908, EPI_ISL_4336910, EPI_ISL_4336913, EPI_ISL_4336915, EPI_ISL_4336918, EPI_ISL_4336921, EPI_ISL_4336923, EPI_ISL_4336926, EPI_ISL_4336929, EPI_ISL_4336931, EPI_ISL_4336935, EPI_ISL_4336938, EPI_ISL_4336941, EPI_ISL_4336943, EPI_ISL_4336945, EPI_ISL_4336948, EPI_ISL_4336951, EPI_ISL_4336954, EPI_ISL_4336957, EPI_ISL_4336959 | | | |
| see above | National Health Laboratory Service, South Africa | KRISP, KZn Research Innovation and Sequencing Platform | Emmanuel S; Emmanuel SJ; Giandhari J; Giandhari Jennifer; Khan S; Lessells R; Maslo C; Mdlalose K; Naidoo Yeshnee; Pillay S; Pillay Sureshnee; San James; Sitharam L; Tegally H; Tegally Houriiyah; Tshabuila Derek; Tshiabuila Derek; Wilkinson E; Wilkinson Eduan; Yajna Ramphal; York D; de Oliveira T; de Oliveira Tulio |
| EPI_ISL_4891491, EPI_ISL_4891508, EPI_ISL_4891550, EPI_ISL_4891582, EPI_ISL_4891589, EPI_ISL_4891599, EPI_ISL_4891608, EPI_ISL_4891609, EPI_ISL_4891628, EPI_ISL_4891673, EPI_ISL_4891697, EPI_ISL_4891706, EPI_ISL_4891860, EPI_ISL_4891870, EPI_ISL_4891871, EPI_ISL_4891872, EPI_ISL_4891873, EPI_ISL_4891874, EPI_ISL_4891876, EPI_ISL_4891877, EPI_ISL_4891878, EPI_ISL_4891888, EPI_ISL_4891889, EPI_ISL_7456449 | | | |
| see above | National Institute For Communicable Diseases Of The National Health Laboratory Service | National Institute for Communicable Diseases of the National Health Laboratory Service | Amoako DG; Bhiman JN; Everatt J; Ismail A; Mahlangu B; Mnguni A; Mohale T; Ntuli N; Scheepers C; Wolter N |
| EPI_ISL_2433846, EPI_ISL_2447877, EPI_ISL_2447879, EPI_ISL_2447891, EPI_ISL_2447892, EPI_ISL_2447895, EPI_ISL_3451535, EPI_ISL_3451536, EPI_ISL_3451537, EPI_ISL_3451538, EPI_ISL_3451539, EPI_ISL_3451540, EPI_ISL_3451541, EPI_ISL_3451542, EPI_ISL_3451543, EPI_ISL_3451544, EPI_ISL_3451545, EPI_ISL_3451546, EPI_ISL_3451547, EPI_ISL_3451548, EPI_ISL_3451549, EPI_ISL_3451550, EPI_ISL_3451551, EPI_ISL_3451552, EPI_ISL_3451553, EPI_ISL_3451554, EPI_ISL_3451555, EPI_ISL_3451556, EPI_ISL_3451557, EPI_ISL_3451558, EPI_ISL_3451559, EPI_ISL_3451560, EPI_ISL_3451561, EPI_ISL_3451562, EPI_ISL_3451563, EPI_ISL_3451564, EPI_ISL_3451565, EPI_ISL_3451566, EPI_ISL_3451595, EPI_ISL_3451596, EPI_ISL_3451597, EPI_ISL_3451598, EPI_ISL_3451599, EPI_ISL_3451600, EPI_ISL_3451601, EPI_ISL_3451602, EPI_ISL_3451603, EPI_ISL_3451604, EPI_ISL_3451605, EPI_ISL_3451606, EPI_ISL_3451607, EPI_ISL_3451608, EPI_ISL_3451609, EPI_ISL_3451610, EPI_ISL_3451611, EPI_ISL_3451612, EPI_ISL_3451613, EPI_ISL_3451614, EPI_ISL_3451615, EPI_ISL_3451616, EPI_ISL_3643813, EPI_ISL_3643828, EPI_ISL_3643839, EPI_ISL_3643858, EPI_ISL_3643874, EPI_ISL_3643908, EPI_ISL_3643920, EPI_ISL_3644042, EPI_ISL_3644043, EPI_ISL_3644045, EPI_ISL_3644047, EPI_ISL_3644048, EPI_ISL_3717893, EPI_ISL_3717894, EPI_ISL_3717897, EPI_ISL_3717902, EPI_ISL_3717904, EPI_ISL_3717905, EPI_ISL_3717926, EPI_ISL_3717937, EPI_ISL_3717939, EPI_ISL_3717944, EPI_ISL_3717945, EPI_ISL_3717949, EPI_ISL_3717959, EPI_ISL_3717960, EPI_ISL_3717962, EPI_ISL_3717970, EPI_ISL_3717992, EPI_ISL_3717996, EPI_ISL_3718049, EPI_ISL_3718051, EPI_ISL_3718052, EPI_ISL_3718053, EPI_ISL_3718054, EPI_ISL_3718055, EPI_ISL_3718056, EPI_ISL_3718057, EPI_ISL_3718058, EPI_ISL_3718059, EPI_ISL_3718060, EPI_ISL_3718061, EPI_ISL_3718062, EPI_ISL_4029931, EPI_ISL_4029936, EPI_ISL_4029953, EPI_ISL_4029957, EPI_ISL_4029976, EPI_ISL_4029978, EPI_ISL_4030018, EPI_ISL_4030079, EPI_ISL_4030080, EPI_ISL_4030081, EPI_ISL_4030082, EPI_ISL_4030083, EPI_ISL_4030106, EPI_ISL_4030107, EPI_ISL_4030108, EPI_ISL_4030109, EPI_ISL_4030110, EPI_ISL_4030117, EPI_ISL_4253673, EPI_ISL_4253674, EPI_ISL_4253675, EPI_ISL_4253676, EPI_ISL_4253677, EPI_ISL_4253678, EPI_ISL_4253679, EPI_ISL_4253680, EPI_ISL_4253681, EPI_ISL_4253682, EPI_ISL_4253683, EPI_ISL_4253684, EPI_ISL_4253685, EPI_ISL_4253686, EPI_ISL_4253687, EPI_ISL_4253688, EPI_ISL_4253689, EPI_ISL_4651523, EPI_ISL_4651531, EPI_ISL_4651532, EPI_ISL_4651540, EPI_ISL_4651548, EPI_ISL_4651554, EPI_ISL_4651559, EPI_ISL_4651565, EPI_ISL_4651574, EPI_ISL_4651575, EPI_ISL_4651580, EPI_ISL_4651581, EPI_ISL_4651588, EPI_ISL_4651595, EPI_ISL_4651596, EPI_ISL_4651601, EPI_ISL_4651602, EPI_ISL_4651610, EPI_ISL_4651616, EPI_ISL_6204412, EPI_ISL_6886554, EPI_ISL_8002897, EPI_ISL_8002898, EPI_ISL_8051878 | | | |
| see above | National Institute for Communicable Diseases of the National Health Laboratory Service | National Institute for Communicable Diseases of the National Health Laboratory Service | Amoako DG; Bhiman JN; Everatt J; Ismail A; Kekana D; Mahlangu B; Mnguni A; Mohale T; Ntuli N; Scheepers C; Wolter N |
| EPI_ISL_1239851, EPI_ISL_1239853, EPI_ISL_1239854, EPI_ISL_1239856, EPI_ISL_1239857, EPI_ISL_1239858, EPI_ISL_1239859, EPI_ISL_1239860, EPI_ISL_1239862, EPI_ISL_1239863, EPI_ISL_1239865, EPI_ISL_1239868, EPI_ISL_1239869, EPI_ISL_1239871, EPI_ISL_1239872, EPI_ISL_1239873, EPI_ISL_1239877, EPI_ISL_1239878, EPI_ISL_1239880, EPI_ISL_1239881 | | | |
| see above | National Institute for Communicable Diseases,National Health Laboratory Services, Gauteng, South Africa | National Institute for Communicable Diseases of the National Health Laboratory Service | Amoako DG; Bhiman JN; Ismail A; Mahlangu B; Mohale T; Ntuli N; Scheepers C |
| EPI_ISL_2955500, EPI_ISL_3118697 | Netcare | KRISP, KZn Research Innovation and Sequencing Platform | Brown MJ; Emmanuel SJ; Giandhari J; Lessells R; Naidoo Y; Omar S; Pillay S; Ramphal U; Tegally H; Wadula J; Wilkinson E; Yajna R; de Oliveira T |
| EPI_ISL_2770489 | Netcare Kingsway | KRISP, KZN Research Innovation and Sequencing Platform | Giandhari Jennifer; Naidoo Yeshnee; Pillay Sureshnee; San James; Tegally Houriiyah; Tshabuila Derek; Wilkinson Eduan; Yajna Ramphal; de Oliveira Tulio |
| EPI_ISL_2693062 | Netcare Kingsway Hospital | KRISP, KZN Research Innovation and Sequencing Platform | Giandhari Jennifer; Naidoo Yeshnee; Pillay Sureshnee; San James; Tegally Houriiyah; Tshabuila Derek; Wilkinson Eduan; Yajna Ramphal; de Oliveira Tulio |
| EPI_ISL_2841651, EPI_ISL_3132628 | Netcare Kingsway Hospital | KRISP, KZn Research Innovation and Sequencing Platform | Giandhari Jennifer; Naidoo Yeshnee; Pillay Sureshnee; San James; Tegally Houriiyah; Tshabuila Derek; Wilkinson Eduan; Yajna Ramphal; de Oliveira Tulio |
| EPI_ISL_3132629 | Netcare Margate | KRISP, KZn Research Innovation and Sequencing Platform | Giandhari Jennifer; Naidoo Yeshnee; Pillay Sureshnee; San James; Tegally Houriiyah; Tshabuila Derek; Wilkinson Eduan; Yajna Ramphal; de Oliveira Tulio |
| EPI_ISL_3132630 | Netcare Parklands Hospital | KRISP, KZn Research Innovation and Sequencing Platform | Giandhari Jennifer; Naidoo Yeshnee; Pillay Sureshnee; San James; Tegally Houriiyah; Tshabuila Derek; Wilkinson Eduan; Yajna Ramphal; de Oliveira Tulio |
| EPI_ISL_3132632, EPI_ISL_3132633, EPI_ISL_3132634 | Netcare St Augustines | KRISP, KZn Research Innovation and Sequencing Platform | Giandhari Jennifer; Naidoo Yeshnee; Pillay Sureshnee; San James; Tegally Houriiyah; Tshabuila Derek; Wilkinson Eduan; Yajna Ramphal; de Oliveira Tulio |
| EPI_ISL_2494975, EPI_ISL_2501091, EPI_ISL_2501092, EPI_ISL_2501093, EPI_ISL_2501094, EPI_ISL_2501095 | Netcare St Augustines Hospital | KRISP, KZN Research Innovation and Sequencing Platform | Emmanuel SJ; Giandhari J; Khan S; Lessells R; Mdlalose K; Naidoo Y; Pillay S; Ramphal U; Sisonke Team; Tegally H; Wilkinson E; York D; de Oliveira T |
| EPI_ISL_2841650 | Netcare The Bay Hospital | KRISP, KZn Research Innovation and Sequencing Platform | Giandhari Jennifer; Naidoo Yeshnee; Pillay Sureshnee; San James; Tegally Houriiyah; Tshabuila Derek; Wilkinson Eduan; Yajna Ramphal; de Oliveira Tulio |
| EPI_ISL_3132631 | Netcare Umhlanga - Chronic patient | KRISP, KZn Research Innovation and Sequencing Platform | Giandhari Jennifer; Naidoo Yeshnee; Pillay Sureshnee; San James; Tegally Houriiyah; Tshabuila Derek; Wilkinson Eduan; Yajna Ramphal; de Oliveira Tulio |
| EPI_ISL_2727404, EPI_ISL_3730372 | Netcare Umhlanga Hospital | KRISP, KZn Research Innovation and Sequencing Platform | Giandhari Jennifer; Naidoo Yeshnee; Pillay Sureshnee; San James; Tegally Houriiyah; Tshabuila Derek; Tshiabuila Derek; Wilkinson Eduan; Yajna Ramphal; de Oliveira Tulio |
| EPI_ISL_3838638 | Private/Pathcare | National Institute for Communicable Diseases of the National Health Laboratory Service | Amoako DG; Bhiman JN; Everatt J; Ismail A; Mahlangu B; Mnguni A; Mohale T; Ntuli N; Scheepers C; Sisonke Team |
| EPI_ISL_7605682 | ROB FERREIRA LABORATORY | National Institute for Communicable Diseases of the National Health Laboratory Service | Amoako DG; Bhiman JN; Everatt J; Ismail A; Mahlangu B; Mnguni A; Mohale T; Ntuli N; Scheepers C; Wolter N |
| EPI_ISL_2086220, EPI_ISL_2086221, EPI_ISL_2086222, EPI_ISL_2086223, EPI_ISL_2086224, EPI_ISL_2086225, EPI_ISL_2086226, EPI_ISL_2086227 | | | |
| see above | ShipMed Inc | KRISP, KZN Research Innovation and Sequencing Platform | Emmanuel SJ; Giandhari J; Lessells R; Naidoo Y; Pillay S; Ramphal U; Tegally H; Wilkinson E; de Oliveira T |
| EPI_ISL_2271962 | Tshepong Hospital | National Institute for Communicable Diseases of the National Health Laboratory Service | Amoako DG; Bhiman JN; Ismail A; Mahlangu B; Mohale T; Ntuli N; Scheepers C |
| EPI_ISL_1324122, EPI_ISL_1324128, EPI_ISL_1324129 | UW Virology Lab | UW Virology Lab | Alexander Greninger; Hong Xie; Keith R Jerome; Lasata Shrestha; Margaret Mills; Meei-Li Huang; Michelle Lin; Noah Baker; Pavitra Roychoudhury; Saraswathi Sathees; Sean Ellis; Shah Mohamed Bakhash |
| EPI_ISL_2494712, EPI_ISL_2494713, EPI_ISL_2494741, EPI_ISL_2494744, EPI_ISL_2494752, EPI_ISL_2494759, EPI_ISL_2494764, EPI_ISL_2494769, EPI_ISL_2494831, EPI_ISL_2494864, EPI_ISL_2494901, EPI_ISL_2494903, EPI_ISL_2494913, EPI_ISL_2494916, EPI_ISL_2494917, EPI_ISL_2494918, EPI_ISL_2494920, EPI_ISL_2494921, EPI_ISL_2494922, EPI_ISL_2494925, EPI_ISL_2494929, EPI_ISL_2494932, EPI_ISL_2494934, EPI_ISL_2494938, EPI_ISL_2494950, EPI_ISL_2494953, EPI_ISL_2494956, EPI_ISL_2494957, EPI_ISL_2494958, EPI_ISL_2494963, EPI_ISL_2494964, EPI_ISL_2494969, EPI_ISL_2494971, EPI_ISL_2494977, EPI_ISL_2494979, EPI_ISL_2494982, EPI_ISL_2494985, EPI_ISL_2494989, EPI_ISL_2494993, EPI_ISL_2495003, EPI_ISL_2495054 | | | |
| see above | Vaccines and Infectious Diseases Analytics Research Unit (VIDA) | KRISP, KZN Research Innovation and Sequencing Platform | Baillie Vicky; Giandhari Jennifer; Madhi Shabir; Naidoo Yeshnee; Pillay Sureshnee; San James; Tegally Houriiyah; Wilkinson Eduan; de Oliveira Tulio; du Plessis Jeanine |
| EPI_ISL_2360343, EPI_ISL_2360344, EPI_ISL_2360345, EPI_ISL_2360346, EPI_ISL_2360347, EPI_ISL_2360349, EPI_ISL_2360351, EPI_ISL_2360352, EPI_ISL_2360354, EPI_ISL_2360356, EPI_ISL_2360357, EPI_ISL_2360358, EPI_ISL_2360359, EPI_ISL_2360361, EPI_ISL_2360364, EPI_ISL_2360365, EPI_ISL_2360367, EPI_ISL_2360368, EPI_ISL_2360369, EPI_ISL_2360372, EPI_ISL_2360373, EPI_ISL_2360374, EPI_ISL_2360375, EPI_ISL_2360377, EPI_ISL_2360378, EPI_ISL_2360379, EPI_ISL_2360380, EPI_ISL_2360381, EPI_ISL_2360384, EPI_ISL_2360385, EPI_ISL_2360387, EPI_ISL_2360388, EPI_ISL_2360389, EPI_ISL_2360391, EPI_ISL_2360392, EPI_ISL_2360395, EPI_ISL_2360396, EPI_ISL_2360397, EPI_ISL_2360412, EPI_ISL_2360413, EPI_ISL_2360418, EPI_ISL_2360419, EPI_ISL_2360420, EPI_ISL_2360421, EPI_ISL_2360422, EPI_ISL_2360423, EPI_ISL_2360424, EPI_ISL_2360425, EPI_ISL_2360449, EPI_ISL_2360450, EPI_ISL_2360451, EPI_ISL_2360452, EPI_ISL_2360453, EPI_ISL_2360454, EPI_ISL_2360455, EPI_ISL_2360456, EPI_ISL_2360457, EPI_ISL_2360458, EPI_ISL_2360459, EPI_ISL_2360469, EPI_ISL_2360470, EPI_ISL_2360471, EPI_ISL_2360472, EPI_ISL_2360473, EPI_ISL_2360474, EPI_ISL_2360475, EPI_ISL_2360476, EPI_ISL_2360477, EPI_ISL_2360478, EPI_ISL_2360479, EPI_ISL_2360480, EPI_ISL_2360481, EPI_ISL_2360482, EPI_ISL_2360483, EPI_ISL_2360484, EPI_ISL_2360485, EPI_ISL_2360486, EPI_ISL_2360487, EPI_ISL_2360488, EPI_ISL_2360489, EPI_ISL_2360490, EPI_ISL_2360491, EPI_ISL_2360492, EPI_ISL_2360493, EPI_ISL_2360494, EPI_ISL_2360495, EPI_ISL_2360496, EPI_ISL_2360497, EPI_ISL_2360498, EPI_ISL_2360499, EPI_ISL_2360500, EPI_ISL_2360501, EPI_ISL_2360502, EPI_ISL_2360503, EPI_ISL_2360504, EPI_ISL_2360505, EPI_ISL_2360506, EPI_ISL_2360507, EPI_ISL_2360508, EPI_ISL_2360509, EPI_ISL_2360510, EPI_ISL_2360511, EPI_ISL_2360512, EPI_ISL_2360513, EPI_ISL_2360514, EPI_ISL_2360515, EPI_ISL_2360516, EPI_ISL_2360517, EPI_ISL_2360518, EPI_ISL_2360524, EPI_ISL_2360525, EPI_ISL_2360526, EPI_ISL_2360527, EPI_ISL_2360528, EPI_ISL_2360529, EPI_ISL_2360550, EPI_ISL_2360551, EPI_ISL_2360552, EPI_ISL_2360554, EPI_ISL_2360557, EPI_ISL_2360570, EPI_ISL_2360571, EPI_ISL_2360572, EPI_ISL_2360573, EPI_ISL_2360574, EPI_ISL_2360575, EPI_ISL_2360576, EPI_ISL_2360597, EPI_ISL_2360598, EPI_ISL_2360601, EPI_ISL_2360608, EPI_ISL_2360612, EPI_ISL_2360613, EPI_ISL_2360614, EPI_ISL_2360617, EPI_ISL_2360634, EPI_ISL_2360638, EPI_ISL_2360642, EPI_ISL_2360643, EPI_ISL_2360644, EPI_ISL_2360645, EPI_ISL_2360646, EPI_ISL_2360647, EPI_ISL_2360649, EPI_ISL_2360660, EPI_ISL_2360661, EPI_ISL_2360666, EPI_ISL_2360667, EPI_ISL_2360669, EPI_ISL_2360670, EPI_ISL_2360671, EPI_ISL_2360675, EPI_ISL_2360677, EPI_ISL_2360678, EPI_ISL_2360679, EPI_ISL_2360680, EPI_ISL_2360692, EPI_ISL_2360693, EPI_ISL_2360694, EPI_ISL_2360695, EPI_ISL_2360701, EPI_ISL_2360703, EPI_ISL_2360704, EPI_ISL_2360707, EPI_ISL_2360708, EPI_ISL_2360709, EPI_ISL_2360712, EPI_ISL_2360721, EPI_ISL_2360722, EPI_ISL_2360723, EPI_ISL_2360730, EPI_ISL_2360731, EPI_ISL_2360732, EPI_ISL_2360733, EPI_ISL_2360735, EPI_ISL_2360736, EPI_ISL_2360737, EPI_ISL_2360738, EPI_ISL_2360739, EPI_ISL_2360740, EPI_ISL_2360741, EPI_ISL_2360743, EPI_ISL_2360744, EPI_ISL_2360745, EPI_ISL_2360746, EPI_ISL_2360758, EPI_ISL_2360763, EPI_ISL_2360764, EPI_ISL_2360765, EPI_ISL_2360766, EPI_ISL_2360768, EPI_ISL_2360771, EPI_ISL_2360772, EPI_ISL_2360775, EPI_ISL_2360776, EPI_ISL_2360782, EPI_ISL_2360785, EPI_ISL_2360787, EPI_ISL_2360792, EPI_ISL_2360794, EPI_ISL_2360828, EPI_ISL_2360829, EPI_ISL_2360839, EPI_ISL_2360849, EPI_ISL_2360851, EPI_ISL_2360852, EPI_ISL_2360854, EPI_ISL_2360862, EPI_ISL_2360863, EPI_ISL_2360864, EPI_ISL_2360865, EPI_ISL_2360871, EPI_ISL_2360872, EPI_ISL_2360876, EPI_ISL_2360881, EPI_ISL_2360882, EPI_ISL_2360883, EPI_ISL_2360889, EPI_ISL_2360892, EPI_ISL_2360893, EPI_ISL_2360902, EPI_ISL_2360903, EPI_ISL_2360904, EPI_ISL_2360905, EPI_ISL_2360907, EPI_ISL_2360908, EPI_ISL_2360909, EPI_ISL_2360910, EPI_ISL_2360916, EPI_ISL_2360917, EPI_ISL_2360918, EPI_ISL_2360919, EPI_ISL_2360920, EPI_ISL_2360921, EPI_ISL_2360925, EPI_ISL_2360926, EPI_ISL_2360934, EPI_ISL_2360935, EPI_ISL_2360940, EPI_ISL_2360941, EPI_ISL_2360944, EPI_ISL_2360954, EPI_ISL_2360955, EPI_ISL_2360962, EPI_ISL_2360964, EPI_ISL_2360966, EPI_ISL_2361002, EPI_ISL_2361013, EPI_ISL_2361014, EPI_ISL_2361018, EPI_ISL_2361026, EPI_ISL_2361027, EPI_ISL_2361047, EPI_ISL_2361048, EPI_ISL_2361049, EPI_ISL_2361050, EPI_ISL_2361051, EPI_ISL_2361052, EPI_ISL_2361053, EPI_ISL_2361054, EPI_ISL_2361055, EPI_ISL_2361056, EPI_ISL_2361057, EPI_ISL_2361058, EPI_ISL_2361067, EPI_ISL_2361071, EPI_ISL_2361072, EPI_ISL_2361073, EPI_ISL_2361074, EPI_ISL_2361077, EPI_ISL_2361078, EPI_ISL_2361079, EPI_ISL_2361080, EPI_ISL_2361081, EPI_ISL_2361082, EPI_ISL_2361083, EPI_ISL_2361084, EPI_ISL_2361085, EPI_ISL_2361086, EPI_ISL_2361087, EPI_ISL_2361088, EPI_ISL_2494358, EPI_ISL_2494361, EPI_ISL_2494362, EPI_ISL_2494370, EPI_ISL_2494375, EPI_ISL_2494377, EPI_ISL_2494383, EPI_ISL_2494384, EPI_ISL_2494385, EPI_ISL_2494401, EPI_ISL_2494410, EPI_ISL_2494415, EPI_ISL_2494419, EPI_ISL_2494467, EPI_ISL_2494473, EPI_ISL_2494477, EPI_ISL_2494483, EPI_ISL_2494496, EPI_ISL_2494497, EPI_ISL_2494498, EPI_ISL_2494518, EPI_ISL_2494521, EPI_ISL_2494524, EPI_ISL_2494533, EPI_ISL_2494535, EPI_ISL_2494544, EPI_ISL_2494545, EPI_ISL_2494546, EPI_ISL_2494548, EPI_ISL_2494550, EPI_ISL_2494554, EPI_ISL_2494561, EPI_ISL_2494565, EPI_ISL_2494571, EPI_ISL_2494572, EPI_ISL_2494573, EPI_ISL_2494597, EPI_ISL_2494599, EPI_ISL_2494614, EPI_ISL_2494629 | | | |
| see above | Vaccines and Infectious Diseases Analytics Research Unit (VIDA) | KRISP, KZn Research Innovation and Sequencing Platform | Baillie Vicky; Giandhari Jennifer; Madhi Shabir; Naidoo Yeshnee; Pillay Sureshnee; San James; Tegally Houriiyah; Tshabuila Derek; Wilkinson Eduan; de Oliveira Tulio; du Plessis Jeanine |
| EPI_ISL_6204413 | Vermaak | National Institute for Communicable Diseases of the National Health Laboratory Service | Amoako DG; Bhiman JN; Everatt J; Ismail A; Mahlangu B; Mnguni A; Mohale T; Ntuli N; Scheepers C |
